# Network-based integrative analysis of lithium response in bipolar disorder using transcriptomic and GWAS data

**DOI:** 10.1101/2022.01.10.21268493

**Authors:** Vipavee Niemsiri, Sarah Brin Rosenthal, Caroline M. Nievergelt, Adam X. Maihofer, Maria C. Marchetto, Renata Santos, Tatyana Shekhtman, Ney Alliey-Rodriguez, Amit Anand, Yokesh Balaraman, Wade H. Berrettini, Holli Bertram, Katherine E. Burdick, Joseph R. Calabrese, Cynthia V. Calkin, Carla Conroy, William H. Coryell, Anna DeModena, Scott Feeder, Carrie Fisher, Nicole Frazier, Mark A. Frye, Keming Gao, Julie Garnham, Elliot S. Gershon, Fernando Goes, Toyomi Goto, Gloria J. Harrington, Petter Jakobsen, Masoud Kamali, Marisa Kelly, Susan G. Leckband, Falk Lohoff, Michael J. McCarthy, Melvin G. McInnis, David Craig, Caitlin E. Millett, Francis Mondimore, Gunnar Morken, John I. Nurnberger, Claire O’ Donovan, Ketil J. Øedegaard, Kelly Ryan, Martha Schinagle, Paul D. Shilling, Claire Slaney, Emma K. Stapp, Andrea Stautland, Bruce Tarwater, Peter P. Zandi, Martin Alda, Kathleen M. Fisch, Fred H. Gage, John R. Kelsoe

## Abstract

Lithium (Li) is one of the most effective drugs for treating bipolar disorder (BD), however, there is presently no way to predict response to guide treatment. The aim of this study is to identify functional genes and pathways that distinguish BD Li responders (LR) from BD Li non-responders (NR). An initial Pharmacogenomics of Bipolar Disorder study (PGBD) GWAS of lithium response did not provide any significant results. As a result, we then employed network-based integrative analysis of transcriptomic and genomic data. In transcriptomic study of iPSC-derived neurons, 41 significantly differentially expressed (DE) genes were identified in LR vs NR regardless of lithium exposure. In the PGBD, post-GWAS gene prioritization using the GWA- boosting (GWAB) approach identified 1119 candidate genes. Following DE-derived network propagation, there was a highly significant overlap of genes between the top 500- and top 2000-proximal gene networks and the GWAB gene list (*P*_hypergeometric_=1.28E- 09 and 4.10E-18, respectively). Functional enrichment analyses of the top 500 proximal network genes identified focal adhesion and the extracellular matrix (ECM) as the most significant functions. Our findings suggest that the difference between LR and NR was a much greater effect than that of lithium. The direct impact of dysregulation of focal adhesion on axon guidance and neuronal circuits could underpin mechanisms of response to lithium, as well as underlying BD. It also highlights the power of integrative multi-omics analysis of transcriptomic and genomic profiling to gain molecular insights into lithium response in BD.

## Introduction

Bipolar disorder (BD) is a major psychiatric disorder characterized by recurrent episodes of mania and depression, and a high risk of suicide. Approximately 50% of BD patients suffer psychosis, and, if left untreated, up to about 17% will complete suicide^1^. Though effective treatments exist, little is understood regarding etiology to guide clinical drug selection or drug design.

Lithium (Li) is the first and remains the best mood stabilizing medication for BD^2, 3^. The mechanism of action of lithium has been studied for over six decades and multiple effects on cellular signaling processes have been identified such as: regulation of GSK3/Akt, G proteins and PKA signaling, inositol turnover, neuronal excitability (via Na+-K+ ATPase), or neurotransmitters^4^. Lithium is clinically effective in treating both mania and depression, but primarily used for prophylaxis. Approximately 30% of patients with BD enjoy a very robust response to lithium with almost complete elimination of symptoms^5, 6^. However, after onset, most patients go through multiple medication trials often over several years during which time they suffer and are at risk for suicide^1^. Many who would be excellent lithium responders never receive a trial of lithium. For these reasons, there is a great need for a predictor of lithium response to guide clinicians in prescribing lithium. Genetics may provide such a predictor as lithium responders have been shown to have a stronger family history of both BD and lithium responsiveness^5^.

One of the challenges of pharmacogenomics is the labor-intensive task of phenotyping. The gold standard for assessing drug response is the prospective clinical trial, but sample sizes for such studies are orders of magnitude smaller (500 vs 50 000) than those currently successful for genome-wide association studies (GWAS). Furthermore, there are few GWAS focusing on lithium response, most suffering from lack of power or failure in replication^7^. Though more data is being accumulated^8–14^, GWAS has so far not had the power to consistently detect reproducible genes.

Human induced pluripotent stem cells (iPSC) provide an alternative and complementary approach to identifying genes and mechanisms of lithium response. It is a revolutionary set of methods enabling access to living neurons from specific individuals and in part overcoming a major hurdle in neuropsychiatric research, the inability to readily access living brain tissue^15^. iPSC methods are now being developed to derive a variety of specific neurons which in turn can model diseases and drug response^16, 17^. For instance, we have previously demonstrated a differential response to lithium *in vitro* between iPSC neurons derived from lithium responders vs non-responders^18, 19^. These data are consistent with the notion that there are two different BD sub-populations with different pathophysiologies defined by lithium response^5^.

Network-based analysis is a powerful bioinformatic approach that employs the fundamental connectivity of gene networks and genetic data to derive models of disease or drug response. Such network models may implicate biological functions associated with complex traits and presumably serve specific cellular processes^20^.

Network-based methods require comprehensive information but have been shown to produce promising insights in studies of various diseases, including psychiatric disorders^21, 22^. Recently, network construction has advanced by integrating multiple sources of data, (e.g., genomic, transcriptomic, proteomic) that, in turn, improves performance^23, 24^. Therefore, a multi-omics network-based approach can be employed in psychiatric research for efficiently uncovering the mechanisms of complex traits and the goal of precision psychiatry^25, 26^.

The mechanisms underlying differential response to lithium in BD remain elusive. In this study, we aim to identify genes associated with lithium response in BD by combining genetic data from a GWAS of lithium response and data from a transcriptome study of iPSC-derived neurons challenged *in vitro* with lithium. The idea behind this integrative analysis is to improve our power to detect genes for lithium response by combining two different independent sources of data and examining the overlap in derived networks.

To our knowledge, this is the first integrative analysis of multi-omics data for lithium response. Here, we describe the results of combining data between a GWAS for lithium response and 41 differentially expressed genes that were identified in iPSC-derived BD neurons from responders (LR) and non-responders (NR). GWAS genes showed a highly significant overlap with the expression-derived network. The functional enrichment analyses identified focal adhesion and the extracellular matrix (ECM) as the most significant biological functions.

## Methods

The methods are summarized here and detailed in **Supplemental Methods**. The overall study design is illustrated in **Figure 1a**.

**Figure 1.**
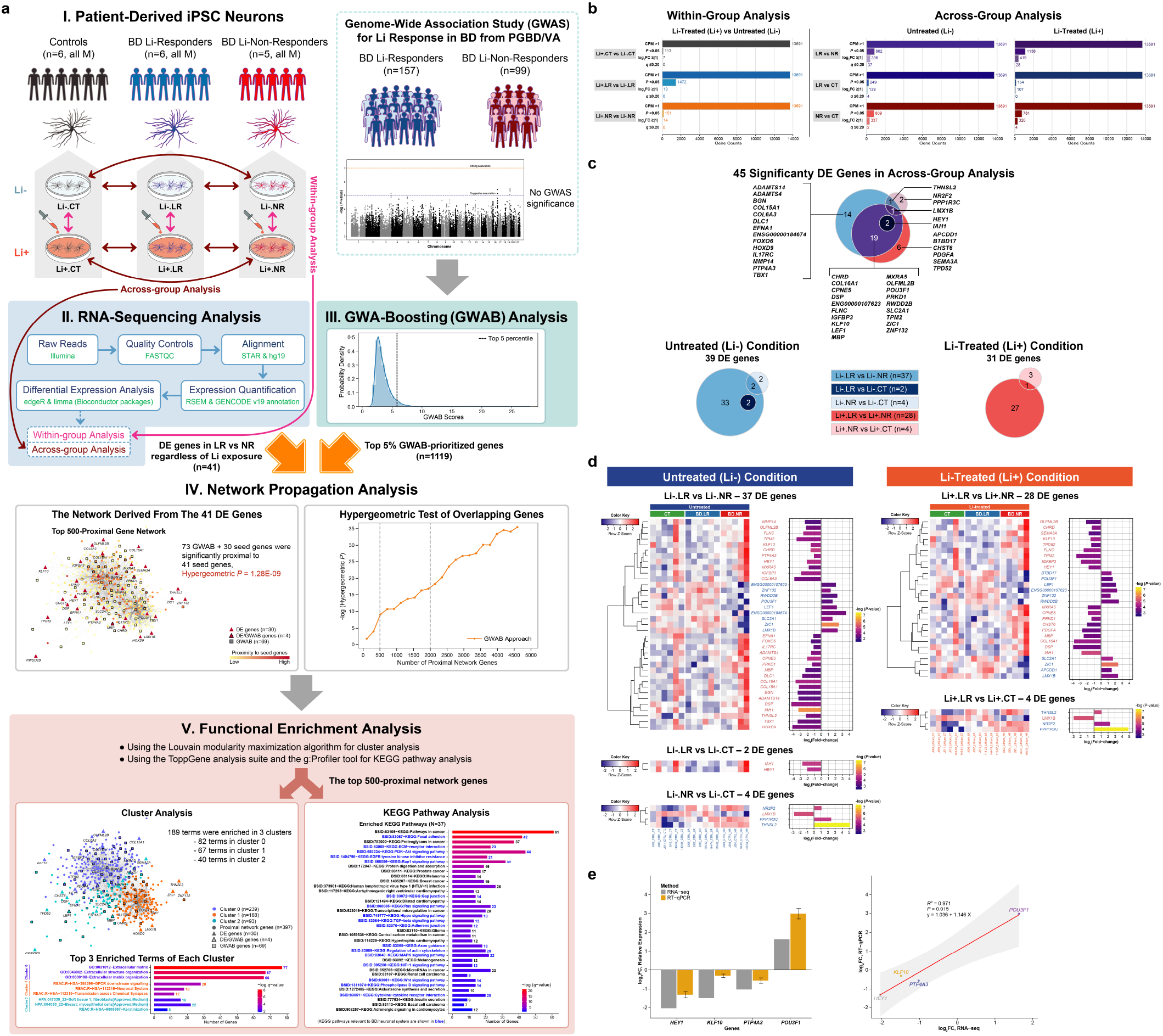
**a. Study design and analysis workflow.** **Figure 1a** shows a summary of our study workflow, including major findings. The study materials and methods are detailed in **Supplemental Methods**. Our comprehensive findings are described in the ‘**Results**’ section. **Step I:** iPSC-derived neurons of three sample groups (6 controls [CT], 6 BD Li responders [BD.LR], and 5 BD Li non-responders [BD.NR]) were tested with *in vitro* lithium. Part of step I is modified from Welham, et al (2015)^73^. **Step II:** RNA-seq pipeline was used for analyzing transcriptomic profiles of all three sample groups under Li- treated (Li+) and untreated (Li-) conditions. The RNA-seq analyses were classified into ‘within-group’ and ‘across-group’. Forty-one significantly DE genes were identified in LR vs NR regardless treatment conditions. **Step III:** The PGBD^12^/VA GWAS for lithium response in BD (*n*=256) was used in a GWA-boosting (GWAB) analysis^31^, which identified the top 5% of GWAB-prioritized genes (1119 genes). **Step IV:** Network propagation analysis^20^ of the top 500-proximal gene network derived from the 41 DE genes revealed a highly significant overlap with the 1119 GWAB-prioritized genes, containing 103 genes (73 GWAB and 30 DE) genes that were significantly proximal to the 41 seed genes with a hypergeometric *P* of 1.28E-09. **Step V:** Functional enrichment analysis of gene clusters and KEGG pathways in the top 500-proximal gene network identified 189 terms significantly enriched in three clusters and 37 significantly enriched KEGG pathways for lithium response in BD. **a. Distribution of RNA-seq genes identified in iPSC-derived neurons.** Bar plots of serial filtering RNA-seq genes for ‘within-group’ (*left* panel) and ‘across- group’ (*right* panel) analyses. Each analysis was categorized into subgroup comparisons (total *n*=9; see comparison details in **Supplemental Methods**). A total of 13 691 RNA-seq genes for each subgroup comparison were removed by serial filters. The RNA-seq genes that passed the serial filtering threshold of CPM >1, *P*-value ≤0.05, log_2_FC >|1|, and B-H *q*-value ≤0.20 were considered as ‘differentially expressed’ (DE) genes. Each subplot shows a plot of the serial filters (y-axis) against the number of genes (x-axis). RNA-seq genes identified in subgroup comparisons with detailed expression data are presented in **Supplementary Tables 4**, **5**, **6**, and **7**. **c-d. Distribution and expression profiles of 45 DE genes identified in across- group analysis of iPSC-derived neurons.** A total of 45 DE genes (*P* ≤0.05, log_2_FC of >|1|, and B-H *q*-value ≤0.20) were identified in ‘across-group’ analysis: 39 genes in untreated (Li-) condition and 31 genes in Li- treated (Li+) condition. Note that out of 45 DE genes, almost all (*n*=43) were found in LR vs NR comparisons: 37 genes in untreated condition and 28 genes in Li-treated condition, including 22 genes that overlapped between them. Lists of DE genes with detailed expression data under two treatment conditions are presented in **Supplementary Table 8**. **b. Venn diagrams of 45 DE genes comparisons among CT, BD LR, and BD NR under two treatment conditions.** **c. Expression of 45 DE genes classified into subgroup comparisons under untreated (Li-) and Li-treated (Li+) conditions.** Each subgroup comparison displays each graph comprising one heatmap and one bar plot. Heatmaps (*left* panel) display hierarchical clustering of gene expression levels for sets of DE genes. Gene symbols (rows) are listed and indicate direction of regulation (down- regulated, light red; up-regulated, light blue). The color scale (*top left*) represents the degree of differential expression (low, blue; high, red). The color boxes above the heatmaps represent sample groups (CT, green; BD.LR, blue; BD.NR, red). Texts (columns) below the heatmap represent samples and are colored by treatment conditions (Li-, dark blue; Li+, orange). Bar plots (*right* panel) present FC expression values (log_2_ transformed) and significance of gene expression (*P* ≤0.05). The color scale (*right*) represents the degree of significance in expression for each gene (low, dark purple; high, yellow), displayed as nominal *P*-values (-log transformed). **d. RNA-seq validation of selected DE genes using RT-qPCR.** Bar plot and scatter plot shows a FC expression comparison of RNA-seq and RT-qPCR results for the four selected DE genes (out of total 37, Figure 1d; **Supplementary Table 8a**) in Li-.LR vs Li-.NR. Bar plot (*left* panel) represents relative FC expression values (y-axis) of the selected four DE genes (x-axis), measured by RNA-seq (grey bars) and RT-qPCR (yellow bars; **Supplementary** Figure 4): *HEY1*, *KLF10*, *PTP4A3* were down-regulated; and *POU3F1* was up-regulated. FC values were presented as in log_2_ unit. RT-qPCR were calculated using the 2^−ΔΔCt^ method. Error bars represent the mean ± SEM of triplicate RT-qPCR data. Scatter plot (*right* panel) shows a high correlation (Pearson *R*^2^ = 0.971, *P* = 0.015) of log_2_FC expression between RNA-seq (x-axis) and RT-qPCR (y-axis) methods. B-H, Benjamini and Hochberg; CPM, counts-per-million; CT, controls; DE, differentially expressed; FC, fold-change; GWAS, genome-wide association study; LR, BD Li responders; NR, BD Li non-responders; RNA-seq, RNA-sequencing; RT-qPCR, real time quantitative PCR; SEM, the standard error of the mean.

All subjects provided written informed consent according to their institution’s approved procedures. Subjects for the GWAS were recruited as part of two studies of lithium response and BD: the multi-site Pharmacogenomics of Bipolar Disorder (PGBD) study, and an identical study of veterans recruited from Veterans Affairs San Diego Healthcare System (VA). The PGBD and veteran studies had a relapse prevention design where subjects were followed prospectively for up to 2.5 years^12^. All subjects had diagnoses confirmed using the Diagnostic Interview for Genetic Studies (DIGS)^27^ and all were of European American (EA) ancestry.

Subjects for the iPSC studies were selected from the PGBD/VA and Halifax samples. The Halifax sample from Dalhouise University was assessed retrospectively using the Alda scale^28^. Both responders and non-responders were selected from the ends of the distribution of response, either time in study (PGBD/VA sample) or Alda score (Halifax sample). Control (CT) subjects were recruited by advertising and screened for psychiatric diagnoses using the DIGS.

GWAS was conducted using the Illumina Human Psychchip on the Illumina Infinium platform (Illumina, San Diego, CA). Genotypes were called using Genome Studio (Illumina). The GWAS analysis employed the “entered_maintenance” phenotype, representing stabilization on lithium monotherapy after 4 months. Analysis began with quality control (QC), followed by alignment and imputation. Association testing used logistic regression in PLINK^29^ with age, sex and three population principal component covariates.

Following the initial analysis of all single nucleotide polymorphisms (SNPs), GWAS results were analyzed using Versatile Gene-Based Association (VEGAS) test^30^, which obtained a single empirical *P-*value for each gene. The Genome-Wide Association Boosting (GWAB) algorithm^31^ was employed in order to use network information to rank order genes.

Skin (PGBD/VA) or blood (Halifax) samples were obtained, and either fibroblasts or lymphoblasts, respectively were reprogrammed to iPSCs and differentiated to *prox1+* hippocampal dentate gyrus glutamatergic granule cells (DG) as described previously^18^ and in **Supplemental Methods**. The six PGBD/VA bipolar and four control cell lines used in this study were identical to the cells reprogrammed as reported previously^18^. The six bipolar subjects in the Halifax sample are also identical to those previously reported^19^. Evidence of pluripotency, normal karyotypes and neural induction has also been previously reported for these lines.

iPSC-derived DG-like neurons from both clinically validated lithium responders and non-responders were treated in culture both with and without lithium. Cells were treated for one week at 1 mM, a clinically effective blood concentration. RNA- sequencing (RNA-seq) was performed on all samples. Ribo-depleted libraries were constructed and cDNA was sequenced as paired ends on an Illumina HiSeq 2500. QC and RNA-seq analysis are detailed in **Supplemental Methods**. Validation of selected genes was performed using reverse transcription quantitative real-time PCR (RT- qPCR).

Initial functional analysis of gene expression was conducted using WebGestalt^32^ and g:Prolifer^33^. Network propagation was used to identify the network regions proximal to the RNA-seq differentially expressed genes^20^. A hypergeometric test was used to test for over-representation of the GWAB genes in the RNA-seq network. Clusters were derived using the Louvain graph-based clustering algorithm^34^ and functional analysis of the derived clusters was performed using ToppGene^35^ and g:Profiler^33^.

## Results

### Subjects

For the GWAS, out of total 728 enrolled subjects, 256 were selected based on completeness of data, Hardy-Weinberg equilibrium, and EA ancestry. Characteristics of the selected GWAS subjects are summarized in **Supplementary Table 1**. NR were significantly more likely to have rapid cycling illness. For the RNA-seq study, overall, there were no significant demographic or clinical differences between LR, NR, and CT groups selected for generation of iPSC-derived neurons (**Supplementary Table 2**).

### GWAS analysis yielded no significant results

As shown in **Supplementary Figure 1**, no SNP was genome-wide significant. The quantile-quantile plot is consistent with inadequate power. A gene-based VEGAS analysis showed similarly negative results (**Supplementary Table 3**).

### RNA-seq analysis

Overall, a total of 13 691 genes were expressed and included in downstream analyses. Filtering out low expression transcripts and using a trimmed mean of *M*- values transformation successfully normalized the expression levels (**Supplementary Figure 2**). We applied two analysis strategies: ‘within-group’ and ‘across-group’ in the RNA-seq analysis, described in detail in **Supplemental Methods**.

### The largest difference in gene expression was between LR and NR without lithium

For the within-group analysis, gene expression in neurons treated with lithium (Li+) was compared to those without lithium (Li-). Employing a significance threshold of *P*-value <0.05 and log_2_fold-change ≥|1|, 14 genes showed nominal significance, but none were significant (Benjamini and Hochberg (B-H) *q* ≤0.20) after multiple testing correction (**Figure 1b**; **Supplementary Figure 3a**; **Supplementary Table 4**). Therefore, lithium had a limited effect on gene expression.

For the across-group analysis, we examined the differential expression of genes between groups of neurons exposed to the same treatment condition. Six comparisons were made: LR vs NR, LR vs CT, and NR vs CT each with (Li+) and without lithium (Li-) (**Supplementary Figure 3b**; **Supplementary Tables 5**, **6**). Using the same significance criteria, a total of 45 genes (43 protein-coding) from all six comparisons were significantly differentially expressed (*P* <0.05, log_2_fold-change ≥|1|, and B-H *q* ≤0.20; ‘DE’ genes; **Figure 1b**). About half of the DE genes (25 of 45) were expressed in more than one comparison, suggesting genetic heterogeneity with small genetic effects across the comparisons. The majority of DE genes (43 of 45, 41 protein-coding) were found in LR vs NR comparisons (**Figures 1c**, **d**). Specifically, out of the 43 genes, the most was 37 DE genes in Li-.LR vs Li-.NR; next was 28 DE genes in Li+.LR vs Li+.NR. Moreover, 22 out of 43 genes (51.16%) were common to LR vs NR between the two treatments. Note that the direction of changes in expression of all 45 DE genes were similar among each comparison regardless of the treatment conditions. No interaction effect was significant (**Supplementary Figure 3c**; **Supplementary Table 7**). The entire list and detailed differential expression of 45 DE genes are shown in **Supplementary Table 8**.

### RT-qPCR validation of selected differential expressed genes

RT-qPCR was performed for technical validation of four selected genes (*HEY1*, *KLF10*, *POU3F1*, and *PTP4A3*) from the 41 protein-coding DE genes in LR vs NR comparisons. These four genes were selected because they were identified in our previous gene expression study of lithium response^36^. RT-qPCR quantification correlated well with RNA-seq for the four genes examined (**Figure 1e**; **Supplementary Figure 4**).

### There was a highly significant overlap of genes between GWAB and DE-derived networks

Using standard GWAS analytic methods, we failed to identify genes significantly associated with lithium response in BD not only in SNP-based but also in gene-based analysis.

In order to prioritize potential candidate genes in the GWAS dataset, we first used a gene-based VEGAS to conduct analysis of the GWAS data. A total of 1180 genes were identified in the top 5% of the VEGAS-prioritized genes, including three genes (*APCDD1*, *DSP*, and *PTP4A3*) shared with the 41 protein-coding DE gene list. We then employed GWAB to boost the GWAS results and obtain prioritized genes utilizing network information^31^. GWAB reprioritizes GWAS genes by boosting “not quite significant” genes that are near other more significant genes in the network. This method reprioritizes genes but does not provide an association statistic for each gene, nor does it provide weights for edges indicating the functional interaction of genes. We identified a total of 1119 genes in the top 5% of the GWAB-prioritized gene list, including four genes (*FLNC*, *LEF1*, *MBP*, and *PRKD1*) that were shared with the DE gene list (*n*=41). The top 5% prioritized genes obtained by VEGAS and GWAB are listed in **Supplementary Table 9**.

We performed network propagation of 41 DE genes using the GIANT brain interactome database^37^ to construct a 500-proximal gene network with 25 020 edges (**Figures 2a-c**). We further boosted the network to a 2000-proximal gene network with 157 688 edges (**Figure 2d**). Out of 41 DE genes, 34 were present in both networks. For the GWAB-prioritized genes, 73 and 241 were detected in the top 500- and top 2000- gene networks, respectively. For the VEGAS-prioritized genes, 36 and 118 were detected in the top 500- and top 2000-gene networks, respectively. The details and genes included in the two networks are presented in **Supplementary Table 10**.

**Figure 2.**
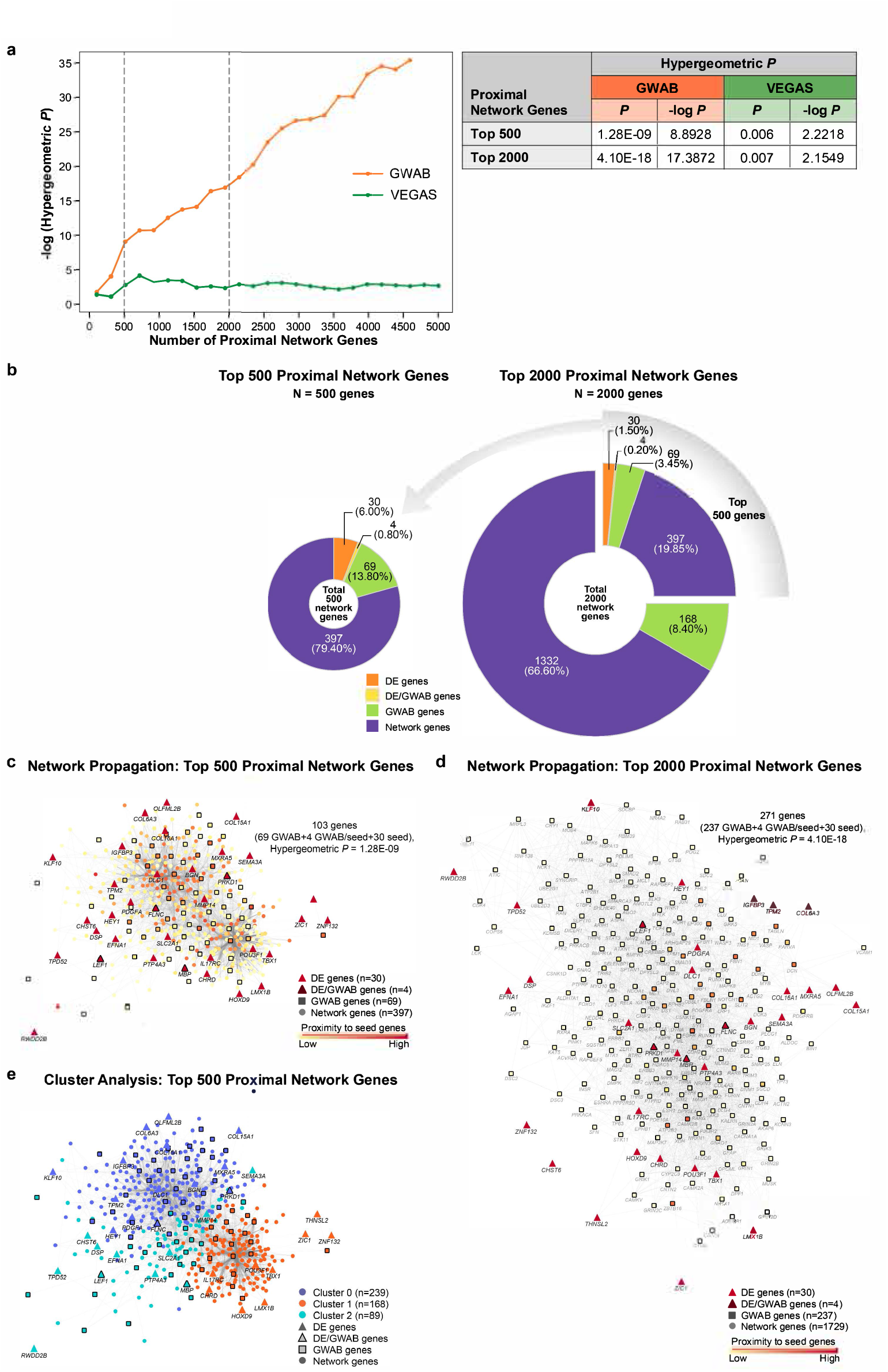
**a. Results of hypergeometric test between the top 500 or top 2000 proximal network genes and the top 5% prioritized genes.** A line graph (*left* panel) plots the -log (hypergeometric *P*-value) against the number of proximal network genes. Each line corresponds to the top 5% prioritized gene sets for GWAB^31^ (orange) and VEGAS^30^ (green). Vertical dashed grey lines at 500 and 2000 (on the x-axis) represents the top 500 and top 2000 proximal network genes, respectively. A table (*right* panel) shows summary of hypergeometric test results. A hypergeometric *P*-value of <0.05 shows the significance of overlap in genes between the top 500 or top 2000 proximal network genes and the top 5% prioritized genes obtained by either GWAB (1119 genes) or VEGAS (1180 genes). See the lists of the top 5% prioritized genes and the top 500 and top 2000 proximal network genes in **Supplementary Tables 9** and **10**, respectively. See the detailed results of the hypergeometric test in **Supplementary Table 11**. **b. Distribution of genes in the top 500- and top 2000-proximal gene networks derived from the 41 protein-coding DE genes.** Pie charts show the number and types of genes in the proximal gene networks: the top 500-proximal gene network (*left*) and the top 2000-proximal gene network (*right*). Each pie chart represents the proportion of gene types as both numbers and percentages. Colors represent gene types (DE, orange; DE/GWAB, yellow; GWAB, green; network, purple). The list of 41 protein-coding DE genes is presented in Figures 1c, **d**, and **Supplementary Table 8**. **c-d. Network propagation of the top 500 and top 2000 proximal network genes.** The network used 41 protein-coding DE genes as the seeds and GIANT interactome^37^ (Tissue-specific gene networks from HumanBase; https://hb.flatironinstitute.org/about) as the background. Color scale on genes (nodes) represents the degree of proximity from the seeds (low/further, yellow; high/nearer, red). Shapes represent gene types. **c. Network propagation of the top 500 proximal network genes.** The network resulted in 500 genes (nodes) connected with 25 025 edges, and contained 30 DE, 4 DE/GWAB, 69 GWAB, and 397 network genes. A subset of 103 (69 GWAB, 4 GWAB/seed, and 30 seed) genes showed statistical significance of the overlap (*P*_hypergeometric_=1.28E-09) in genes between the top 500-proximal gene network and top 5% GWAB-prioritized data, two independent sources of data. The interactive graph of the top 500-proximal gene network can be accessed at https://ndexbio.org/viewer/networks/8ddb8cc6-aea3-11eb-9e72-0ac135e8bacf. **d. Network propagation of the top 2000 proximal network genes.** The network resulted in 2000 genes (nodes) connected with 157 688 edges, and contained 30 DE, 4 DE/GWAB, 237 GWAB, and 1729 network genes. Note that a subset of 271 (30 DE, 4 DE/GWAB, and 241 GWAB) genes with 3135 edges are shown here. These relevant 271 genes showed statistically significant overlap (*P*_hypergeometric_=4.10E-18) between the top 2000-proximal gene network and top 5% GWAB-prioritized data. The interactive graph of the top 2000-proximal gene network can be accessed at https://ndexbio.org/viewer/networks/8ddb8cc6-aea3-11eb-9e72-0ac135e8bacf. **e. Cluster enrichment analysis of the top 500-proximal gene network derived from the 41 protein-coding DE genes.** Three clusters were identified based on biological functions. Colors on genes (nodes) indicate for each cluster (0, blue; 1, orange; 2, green). Shapes represent gene types. The basic network prior cluster enrichment analysis is displayed in Figure 2c. See the list and distribution of genes for each cluster in **Supplementary Table 10**, and Figure 3, respectively. The interactive graph of cluster enrichment for the top 500-proximal gene network can be publicly accessed at https://ndexbio.org/viewer/networks/43073550-aea3-11eb-9e72-0ac135e8bacf. DE, differentially expressed; GWAB, genome-wide association boosting; VEGAS, versatile gene-based association study.

Next, we examined the significance of the gene overlap between the DE-derived top 500- or top 2000-proximal gene networks and the 1119 GWAB-prioritized genes. This overlap was striking and highly significant (hypergeometric *P*=1.28E-09 for the top 500 network genes; 4.10E-18 for the top 2000 network genes). In contrast, the overlap between the DE-derived gene networks and 1180 VEGAS-prioritized genes showed nominal significance (hypergeometric *P*=0.006 for the top 500 network genes; 0.007 for the top 2000 network genes) (**Figure 2a**; **Supplementary Table 11**). The highly significant overlap from independent data sources suggests convergence on valid common biological functions.

### Focal adhesion and the extracellular matrix were the most enriched biological functions

To gain the biological insights into the 41 DE genes, we initially explored the functional enrichment of the 41 gene set using the WebGestalt^32^ and g:Profiler^33^ analysis tools. The results in **Supplementary Figure 5** show that the highest ranked functions were related to ‘focal adhesion’ (KEGG; WebGestalt) and the ‘extracellular matrix’ (REACTOME; g:Profiler). However, no significant enrichment (*P* <0.05 with B-H *FDR* <1.0E-05) was observed in either test.

We further evaluated the functional enrichment of the genes in the top 500-gene network proximal to the 41 DE gene seed set. Cluster enrichment analysis of the top 500 proximal network genes identified three clusters (**Figure 2e**) with 189 enriched terms (B-H *q* <0.05, range=0.0498-1.01E08), comprising clusters 0, 1, and 2, with 82, 67, and 40 functional terms, respectively. The enrichment for each of the three clusters, including the top 10 terms and gene contribution are summarized in **Figure 3**. The details of each functional term for each cluster are described in **Supplementary Figure 6** and **Supplementary Tables 12-14**.

**Figure 3.**
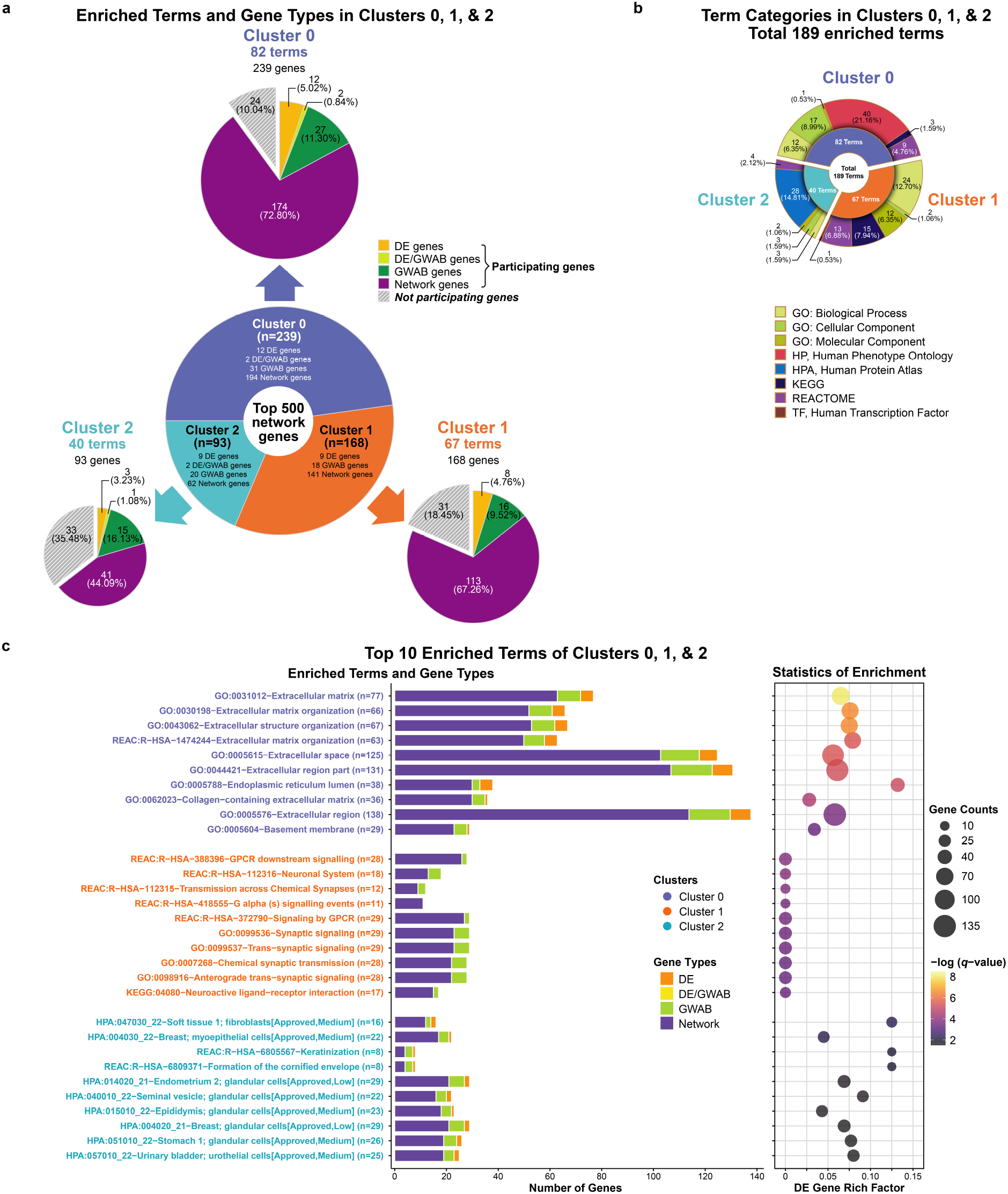
**a-c. Significantly enriched terms (B-H *q*-value ≤0.05) identified in cluster enrichment analysis of the top 500-proximal gene network derived from the 41 protein-coding DE genes.** The cluster analysis identified three clusters: 0 (blue), 1 (orange), and 2 (green), containing 239, 168, and 93 genes, respectively. A total 189 functional terms were significantly enriched (B-H *q*-value ≤0.05) among the three clusters. Overall, cluster 0 had the most enriched terms, of these, the term with the greatest enrichment was ‘extracellular matrix’ (GO:0031012; B-H *q*=1.01E-08). Details of corresponding terms for clusters 0, 1, and 2 are listed in **Supplementary Tables 12**, **13**, and **14**, respectively. The distribution of genes for each term is summarized in **Supplementary** Figure 6. **a. Distribution of significantly enriched terms and gene sets in three clusters of the top 500-proximal gene network.** The sunburst diagram (*central*) shows the proportion of genes involved in each of the three clusters. Each subordinate pie chart (*marginal*) shows the proportion of genes involved in significantly enriched terms for each cluster. Colors on the subordinate pie charts represent gene types. Solid and stripe patterns indicate functional (participating) and non-functional (non-participating) of the genes in each cluster, respectively. The size of each pie slice corresponds to the number of genes. The proportion is presented as both numbers and percentages. **b. Categories of significantly enriched terms in three clusters of the top 500- proximal gene network.** A sunburst diagram corresponds to a total of 189 significantly enriched terms, comprising of 82, 67, and 40 terms enriched in clusters 0, 1, and 2, respectively. Among six various categories, the Gene Ontology (GO) and Human Phenotype Ontology (HP) were the majority of enrichment in cluster 0; while GO and Human Proteome Atlas (HPA) were the most terms enriched in clusters 1 and 2, respectively. The size of each slice of the pie chart corresponds to the number of enriched terms. Each proportion is displayed as both numbers and percentages. The *inner* ring shows the proportion of enriched terms for each of the three clusters, colored by clusters. The *outer* ring shows the proportion of biological term categories for each cluster, colored by categories of enriched terms. **c. Identification and summarizing features of the top 10 significantly enriched terms of clusters 0, 1, and 2.** In brief, cluster 0 had the terms with the greatest enrichment, and almost all the top 10 terms mainly involved in the extracellular matrix (ECM). The top 10 significantly enriched terms in cluster 1 tended to involve in neuronal systems and synapses. Whereas those in cluster 2 showed various tissue-specific association other than neuronal tissues. Bar plot (*left* panel) shows the number and types of genes in each of the top 10 enriched terms for each cluster. The x-axis represents the number of genes (gene counts) and gene types distributing to each corresponding term. The y-axis corresponds to significantly enriched terms grouped by clusters, shown in text colors. Colors on bars indicate gene types. Bubble plot (*right* panel) shows the statistics of enrichment for each of the enriched terms. The size of bubbles represents the number of genes (gene counts) in each corresponding term. The bubble coordinate on the x-axis represents the degree of enrichment for DE genes known as ‘DE gene rich factor’. The DE gene rich factor is the ratio of DE genes in each term to total genes in each term. The larger rich factor represents the greater enrichment. The color scale indicates the degree of significance (B-H *q*-value ≤0.05) in enrichment for each corresponding term (low, dark purple; high, yellow). The significance of enrichment is presented as the -log transformed B-H *q*- value. Gene types are indicated by colors: DE, orange; DE/GWAB, light green (**3a**) or yellow (**3c**); GWAB, green; network, purple. B-H, Benjamini and Hochberg; DE, differentially expressed; GWAB, genome-wide association boosting.

Overall, cluster 0 had the most enriched terms. Strikingly, almost all of the top 10 enriched terms in cluster 0 involved in the ECM, including the term with the greatest enrichment—‘extracellular matrix’ (GO:0031012; B-H *q*=1.01E-08), suggesting that cluster 0 mainly represented an ECM-related subnetwork. Whereas, the top ranked terms in cluster 1, such as G-protein-coupled receptors (GPCR) signaling and neuronal transmission, yielded subnetworks relevant to neuronal systems. The greatest enrichment in cluster 1 was ‘GPCR downstream signaling’ (REAC:R-HSA-388396; B-H *q*=1.57E-04). In contrast, most of the enriched terms in cluster 2 were related to tissue- specific functions, but not strongly significant. Only one term (HPA:007010_22, cerebellum; Purkinje cells; B-H *q*=0.04) was related to neurons. The greatest enrichment in cluster 2 (B-H *q*=0.0054) included ‘soft tissue 1; fibroblasts’ (HPA:047030_22) and ‘breast; myoepithelial cells’ (HPA:004030_22). Thus, the major findings of cluster analysis implicate the ECM in lithium response.

The KEGG pathway enrichment analysis of the top 500 proximal network genes revealed a total of 37 KEGG pathways that were significantly enriched (B-H *q* <0.05, range*=*0.02409-1.05E-21) among 196 genes (**Figures 4a**, **b**; **Supplementary Figure 7**; **Supplementary Table 15**). The 37 significant KEGG pathways involved four main KEGG categories: cellular processes, environmental information processing, human diseases, and organismal systems, of which the one with the greatest enrichment was ‘pathways in cancers’ (hsa05200; B-H *q*=1.05E-21), followed by ‘focal adhesion’ (hsa04510; B-H *q*=8.04-E20) as the second highest enrichment.

**Figure 4.**
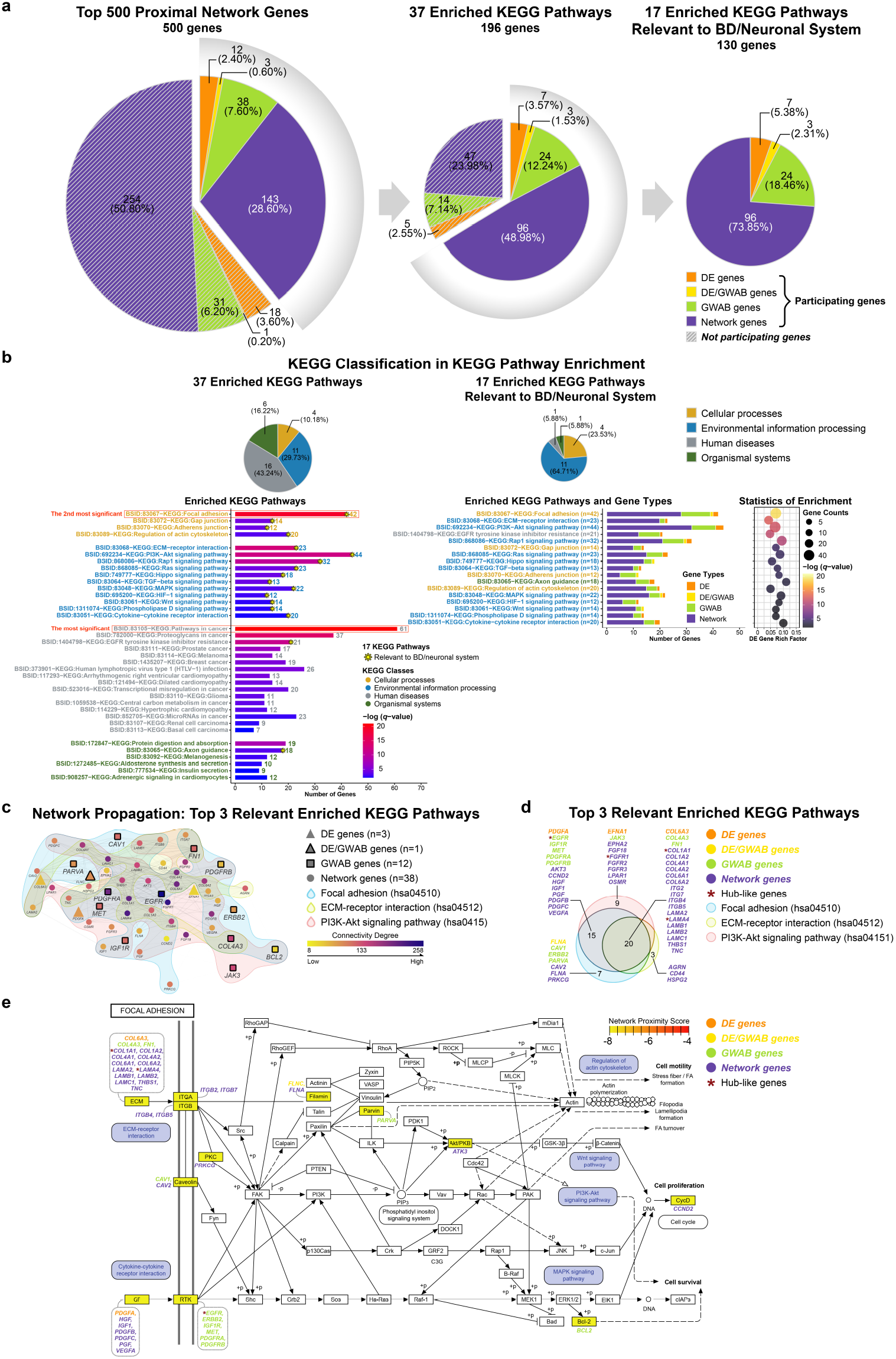
**a-b. Significantly enriched KEGG pathways (B-H *q*-value ≤0.05) identified in KEGG pathway enrichment analysis of the top 500-proximal gene network derived from the 41 protein-coding DE genes.** The KEGG pathway analysis (containing at least one seed gene and one GWAB gene) identified a total of 37 KEGG pathways, including a subset of 17 KEGG pathways relevant to BD/neuronal system, which were significantly enriched (B-H *q*-value ≤0.05) in the top 500-proximal gene network. Details for each pathway are listed in **Supplementary Table 15**. The distribution of genes for each pathway is summarized in **Supplementary** Figure 7. **a. Summary of significantly enriched KEGG pathways and gene sets in the top 500-proximal gene network.** Pie charts display the number of genes from each source that comprise the top 500- proximal gene network (*left*), a set of 196 genes of the 37 enriched KEGG pathways (*middle*), and a set of 130 genes of the 17 relevant enriched KEGG pathways (*right*). Each pie represents the proportion of gene types as both numbers and percentages. Colors represent gene types. Solid and stripe patterns indicate function (participating) and non-function (non-participating) of genes, respectively, in each gene set of either 37 or 17 enriched KEGG pathways. **b. KEGG pathway classification and enrichment statistics of significantly enriched KEGG pathways for the top 500-proximal gene network.** A total of 37 enriched KEGG pathways is on the *left* panel, and a subset of 17 relevant enriched KEGG pathways is on the *right* panel. These pathways were classified into four classes: cellular processes (yellow), environmental information processing (blue), human diseases (grey), and organismal systems (green). Of the 37 enriched KEGG pathways, the most significant enrichment was ‘pathways in cancer’ (hsa05200; B-H *q*=1.05E-21; see red box). The second most significant enrichment was ‘focal adhesion’ (hsa04510; B-H *q*=8.04-E20; see red box), which was also the most significant one among the 17 relevant enriched KEGG pathways. Of the 17 relevant enriched KEGG pathways, the top 3 significant enrichment were ‘focal adhesion’ (as mentioned), ‘ECM-receptor interaction’ (hsa04512; B-H *q*=1.57E-13), and ‘PI3K-Akt signaling pathway’ (hsa04151; B-H *q*=9.62E-13), respectively. Pie charts (*top* panel) show the distribution of KEGG pathway classification. Each pie displays the proportion of KEGG classes as both numbers and percentages, colored by KEGG classes. Bar and bubble plots (*bottom* panel) show the distribution of genes and/or the statistical enrichment for each of the enriched KEGG pathways, respectively. Bar plots show the distribution of genes for each KEGG pathway. The x-axis represents the number of genes contributing to each corresponding pathway. The y-axis corresponds to significantly enriched KEGG pathways categorized into KEGG classes and shown in text colors. For the 37 enriched KEGG pathways (*bottom left* panel), the color scale on bars indicates the degree of significance (B-H *q*-value ≤0.05) of enrichment (low, blue; high, red). An asterisk (*) on bars specifies the KEGG pathways that are relevant to BD/neuronal system. Whereas, for a subset of the 17 relevant enriched KEGG pathways (*bottom middle* panel), colors on bars indicate gene types. Bubble plot (*bottom right* panel) shows the statistical enrichment of a subset 17 relevant enriched KEGG pathways. The size of bubbles represents the number of genes (gene counts) in each corresponding KEGG pathway. The bubble coordinate on the x-axis represents the degree of enrichment for DE genes known as ‘DE gene rich factor’. The DE gene rich factor is the ratio of DE genes in each pathway to total genes in each pathway. The larger rich factor represents the greater enrichment. The color scale indicates the degree of significance (B-H *q*-value ≤0.05) in enrichment for each corresponding pathway (low, dark purple; high, yellow). The significance of enrichment is presented as the -log transformed B-H *q*-value. **c-e. Identification and summarizing features of the top 3 significantly enriched KEGG pathways relevant to BD/neuronal system (B-H *q*-value ≤0.05).** The top 3 of the 17 relevant significantly enriched KEGG pathways are ‘top 1’ - focal adhesion (hsa04510; light blue), ‘top 2’ - ECM-receptor interaction (hsa04512; light yellow), and ‘top 3’ - PI3K-Akt signaling pathway (hsa04151; light pink), respectively. A subset of 54 genes (out of the top 500 proximal network genes) are present in the top 3 relevant enriched KEGG pathways. **c. Sub-networks among the 54 genes in the top 3 significantly enriched KEGG pathways relevant to BD/neuronal system.** A propagation-based network illustrates sub-networks among a set of 54 genes (out of the top 500 proximal network genes) present in the top 3 relevant enriched KEGG pathways. This network included 54 genes (nodes) connected with 506 edges, containing 3 DE, 1 DE/GWAB, 12 GWAB, and 38 network genes. Shapes represent gene types. Color scale indicates the degree of connectivity calculated from the top 500 proximal network genes (low degree/poorly connected, yellow; high degree/highly connected, dark purple). Background colors are specified for the top 3 relevant enriched KEGG pathways. The connectivity of the 54 proximal network genes of the top 3 relevant enriched KEGG pathways is presented in **Supplementary 16**. **d. Venn diagram of the 54 genes in the top 3 significantly enriched KEGG pathways relevant to BD/neuronal system.** A total of 54 gene names are listed and categorized into gene types. There are 42, 23, and 44 genes involved in the top 1, top 2, and top 3 relevant enriched KEGG pathways, respectively, including 20 overlapping genes and four ‘hub-like’ genes. Venn diagram colors indicate the top 3 relevant enriched KEGG pathways. **e. Pathview of focal adhesion (hsa04510), the top significantly enriched KEGG pathway relevant to BD/neuronal system.** The top relevant significantly enriched KEGG pathway,’ focal adhesion’ (B-H *q*- value=8.04E-20), contained a total of 42 genes (including three ‘hub-like’ genes), which are listed and categorized into gene types. This figure also illustrates several KEGG pathways connected with focal adhesion. Those relevant significantly enriched pathways (B-H *q*-value ≤0.05) identified in our study are highlighted in blue. The color scale indicates the degree of network proximity to seed genes (more proximal/farther, yellow; less proximal/nearer, red). Pathview map created by the R-based Pathview^74^ software from Bioconductor. Gene types are indicated by colors: DE, orange; DE/GWAB, yellow; GWAB, green; network, purple. An asterisk (*) in dark red represents the ‘hub-like’ genes. B-H, Benjamini and Hochberg; DE, differentially expressed; ECM, the extracellular matrix; GWAB, genome-wide association boosting.

Of these 37 KEGG pathways, 17 were selected a priori based on their involvement in brain function and relevant involvement in pathways modulated by lithium, such as PI3K-Akt signaling, Ras signaling, MAPK signaling, and Wnt signaling (**Figures 4a**, **b**). These 17 relevant pathways with 130 network genes represented 26.0% of top 500 network genes, including 7 DE, 3 DE/GWAB, and 24 GWAB genes. Of the 17 relevant KEGG pathways, the top 3 were ‘focal adhesion’ (hsa04510; B-H *q*=8.04-E20), ‘ECM-receptor interaction’ (hsa04512; B-H *q*=1.57E-13), and ‘PI3K-Akt signaling pathway’ (hsa04151; B-H *q*=9.62E-13), respectively (**Figure 4b**). These top 3 pathways were significantly overrepresented in a subset of 54 network genes (10.8% of top 500 network genes). Note that 20 out of 54 genes overlapped among the top 3 relevant KEGG pathways (**Figures 4c**, **d**). The top relevant KEGG pathway — ‘focal adhesion’ with 42 network genes, is illustrated in **Figure 4e**, which also shows the connection between focal adhesion and other enriched pathways.

Notably, the enrichment of the top 500-proximal gene network was consistent with the preliminary enrichment of the 41 DE gene set obtained via WebGestalt^32^ and g:Profiler^33^ (**Supplementary Figure 5**). Likewise, **Figure 5a** summarizes the UniProtKB functions^38^ of the 41 DE genes, which were associated with cell/focal adhesion, the ECM, neurogenesis, including development of axon and synapse. Similarly, the UniProtKB function^38^ of 22 genes (**Figure 5b**) shared by gene sets of the top 3 terms of cluster 0 (91 genes) and the top 3 relevant KEGG pathways (54 genes) were involved in the same functions.

**Figure 5.**
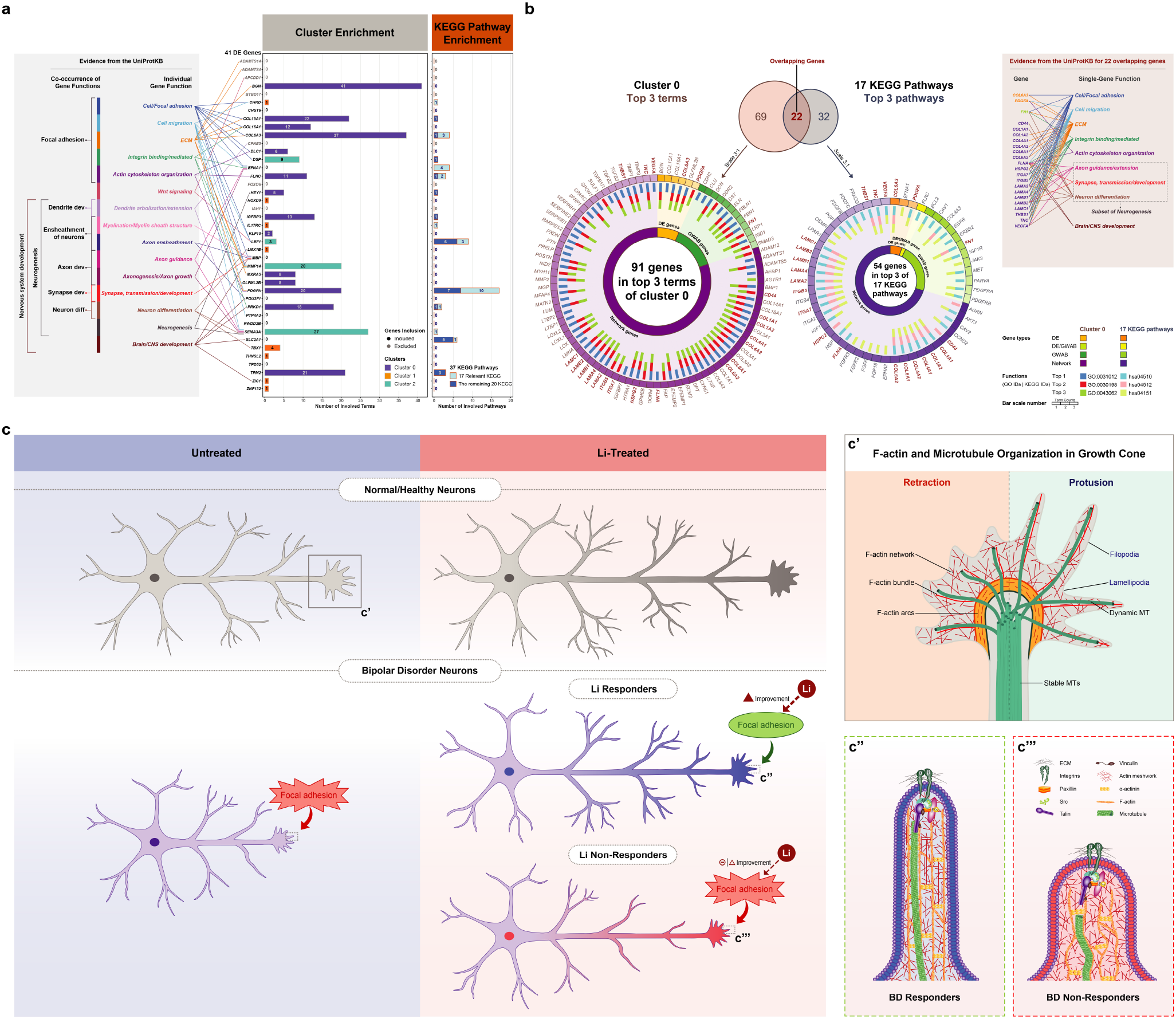
**a. Summary of functional annotation for 41 protein-coding DE genes: individual- and multi-gene functions.** The bar plots show the functions of each protein-coding DE gene (*n*=41; **Supplementary Table 8**) based on the functional enrichment analysis of the 500- proximal gene network from this study. The 41 DE genes are annotated in the three clusters (clusters: 0, blue; 1, orange; 2, green) (**Supplementary Tables 12**, **13**, and **14**, respectively), shown on the *left* bar plot, and annotated in the 37 significantly enriched KEGG pathways (17 relevant enriched KEGG, light blue; the remaining 20 KEGG, dark blue) (**Supplementary Table 15**), shown on the *right* bar plot. Note that, out of 41, seven genes (*ADAMTS14*, *ADMATS4*, *APCDD1*, *BTBD17*, *CPEN5*, *FOXO6*, and *IAH1*; texts in grey) were excluded from the top 500-proximal gene network prior the functional enrichment analysis. Another six (*CHST6*, *MBP*, *POU3F1*, *PTP4A3*, *RWDD2B*, and *TPD52*) out of 34 remaining genes were not in either cluster or KEGG functions. The x-axis indicates the number of participating terms in clusters or participating pathways in KEGG functions. The y-axis indicates a list of 41 protein-coding DE gene names in alphabetical order. The tubular texts (*left* box) show the single and co-occurrence gene functional modules based on the UniProt Knowledgebase evidence^38^ (UniProtKB, https://www.uniprot.org/). The functions for each DE gene and co-occurrence genes are shown in *right* and *left* tableau, respectively. Among the single-gene functions, the majority were ‘cell/focal adhesion’, ‘cell migration’, and ‘extracellular matrix’ (ECM). While the co-occurrence functions in multi-genes also tended to cluster for ‘focal adhesion’, ‘nervous system development’, including ‘neurogenesis’ and ‘axon development’. **a. Distribution and summary of gene sets in the top 3 cluster 0 significantly enriched terms (91 genes) and the top 3 17 relevant significantly enriched KEGG pathways (54 genes) (B-H *q*-value ≤0.05).** Venn diagrams (*top* panel) show the numbers of genes participating in the top 3 cluster 0 significantly enriched terms (91 genes, tan; **Supplementary** Figure 6d; **Supplementary Table 12**) and the top 3 17 relevant significantly enriched KEGG pathways (54 genes, grey; Figures 4c, **d**; **Supplementary** Figure 7; **Supplementary Table 15**), including 22 genes overlapping (dark red bold) between them. The size of the Venn diagrams is shown as proportion to a size of 500 genes. Doughnut charts (*bottom* panel) exhibit the distribution of gene types and functions for the top 3 cluster 0 significantly enriched terms (*left*) and the top 3 17 relevant significantly enriched KEGG pathways (*right*). (**i**) *Inner* ring represents gene types: DE (orange), DE/GWAB (yellow), GWAB (green), and network (purple) genes. (**ii**) *Middle* ring represents numbers of terms/pathways in which each gene was participating, shown as bar scale (range from 1 to 3). (**iii**) *Outer* ring represents gene names that are listed in alphabetical order and colored by gene types. Overlapping genes are highlighted in dark red bold. A total of 22 overlapping genes including their single-gene functions based on evidence in the UniProt Knowledgebase^38^ (UniProtKB, https://www.uniprot.org/) are listed (*top right* box). The functions among 22 genes are mainly involved in ‘cell/focal adhesion’, ‘cell migration’, ‘ECM’, ‘neurogenesis’, including ‘axon guidance/extension’, which are similar to the functions of 41 protein-coding DE genes (Figure 5a, *left* box). **b. Schematic of a proposed model of focal adhesion and its role in lithium response in BD. Boxes are magnified for physiological cytoskeleton dynamics (5c’) and a proposed model (5c’’- 5c’’’) at the neuronal growth cone.** As shown in Figure 5c, based on our KEGG pathway enrichment result, we hypothesize that a defect in focal adhesion and the ECM, including the integrin-ECM interactions, accounts for an underlying mechanism of lithium response in BD. However, to date, the assessment of focal adhesion function in BD, including the cytoskeleton dynamics and morphology of growth cones in BD neurons, has not yet been well studied. In addition, lithium has been reported to have positive effects on axonal and growth cone morphology of normal neurons^64–68^ (*top right* panel). Moreover, without drug treatments, collective evidence favors the pathological morphology^69–71^ in BD neurons (in purple) compared with healthy neurons (in grey) (*bottom left* panel). Altogether, we propose that, lithium treatment (*bottom right* panel) results in substantial improvement of axon guidance and synaptic connectivity in BD responder neurons (with blue axon), thereby correcting an underlying causative defect and resulting in successful treatment in BD responders. Whereas, BD non-responder neurons (with red axon) may have BD not only due to focal adhesion but also due to other different molecular etiologies not involving focal adhesion, and therefore not responsive to lithium. **(5c’). Physiological cytoskeleton dynamics.** The neuronal growth cone is a motile structure at the peripheral tip of the axon, which is enriched in two cytoskeletal filaments—filamentous actin (F-actin) and microtubules (MTs). The growth cone is essential in axon outgrowth, guidance and pathfinding in order to form proper synaptic connections. The interaction between actin filaments and MTs is a dynamic process resulting in protrusion or retraction along the edge of growth cones^50–54^. In brief, the protrusion of filopodia (F-actin parallel bundles) and lamellipodia (F-actin meshwork) are responsive to extrinsic attractive cues in the ECM. This induces actin polymerization and subsequent coupling of F-actin and MTs for MT polymerization and stabilization, resulting in growth cone turning and axon outgrowth. Whereas, retraction occurs in response to repulsive guidance cues in the ECM by F-actin severing, disassembly of F-actin and MTs, and MT depolymerization/destabilization. To develop proper axon outgrowth, focal adhesion is required as an adhesive linkage between the cytoskeleton and the ECM that regulates actin cytoskeletal organization (see **5c’’- c’’’**), thereby enabling growth cone behavior and axon guidance^58, 59^. This figure is adapted from Vitriol and Zheng (2012)^53^ and Cammarata, Bearce, and Lowery (2016)^54^ with permission. **(5c’’- 5c’’’). Focal adhesion at growth cones of BD neurons in response to lithium in the proposed model.** Focal adhesion plays a pivotal role in linking the cytoskeleton to ECM via an interaction between transmembrane integrin receptors and ECM proteins (i.e., laminins, collagens, and fibronectin)^46–49^. Focal adhesions and integrin-ECM interactions occur in neuronal cells, mainly at the tip of growth cones, named point contact (PC). At PC, several signaling and adhesion molecules such as FAK-Src, paxillin, vinculin, talin, and Cas-csk are recruited and form the focal adhesion complex^46–49^, which regulates actin cytoskeleton organization downstream of the focal adhesion complex^50–54^. Thus, focal adhesion has complex, dynamic functions and is essential for regulating axon outgrowth and guidance in response to axon guidance cues (see **c’**), which leads to growth cone turning and synapse formation in neuronal cells^58, 59^. Our hypothesized model is described in detail here. In BD responder neurons (**5c’’**) treated with lithium, focal adhesion dysfunction is corrected resulting in strong and well-organized actin filaments and MTs. Vigorous filopodia and lamellipodia are restored at the growth cone, and axon elongation and growth cone dynamics (protrusion and retraction, see **5c’**) are normalized. This suggests that focal adhesions can be rescued and preserved by lithium effects in BD responder neurons, leading to response to lithium treatment in BD responders. In contrast, for BD non-responder neurons (**5c’’’**), when treated with lithium, focal adhesion function remains defective perhaps due to inefficient effects of lithium on those neurons, resulting in persistently weak and poorly-organized actin and microtubule structures. Unwell filopodia and lamellipodia at the growth cone cause poor neuronal axon guidance and disrupted synaptic formation. Therefore, lithium has no major effects on axonal outgrowth and growth cone morphology in BD non-responder neurons, leading to no response to lithium in BD non-responders. More details and references are provided in the ‘**Discussion**’ section. B-H, Benjamini and Hochberg; Cas, Crk-associated substrate; csk, c-terminal Src kinase; DE, differentially expressed; ECM, the extracellular matrix; FAK, focal adhesion kinase; GWAB, genome-wide association boosting.

Hub-like genes are defined as genes that have a high degree of connectivity among pathways. Among a total of 90 genes from the top 3 of 37 KEGG pathways, seven genes (*EGFR*, *TGFB2*, *CAMK2B*, *FGFR1*, *TIMP3, LAMA4*, and *COL1A1*) showed the highest degree of connectivity (degree >200, range=202-258; **Supplementary Table 16**), which were considered ‘hub-like’ genes. Out of these seven, four genes were also found in a 54-gene subset of the top 3 of 17 KEGG pathways (**Figures 4c**, **d**). The top scoring gene (*EGFR)* was involved in all three pathways. The remaining three genes (*FGFR1*, *LAMA4*, and *COL1A1*) were involved in one or two pathways. Noticeably, all seven hub-like genes were not DE; but rather GWAB or network genes, since only 34 DE genes (6.80%) were part of the top 500- proximal gene network.

## Discussion

In this study, we have combined GWAS and transcriptomic data to improve our overall power to detect genes and biological functions associated with lithium response. Analysis of the RNA-seq data indicated that the largest overall difference was between LR and NR, particularly in the absence of lithium, implying that inter-individual differences are a larger effect than the effect of lithium. In addition, the effect of lithium was similar between LR and NR subjects. We further demonstrated a highly significant overlap of the DE gene-propagated network and GWAB-prioritized genes (*P*_hypergeometric_=1.28E-09 for the top 500-proximal gene network; 4.10E-18 for the top 2000-proximal gene network), indicating that GWAB genes are in the same gene neighborhoods as the 41 DE genes. Functional enrichment analyses of the 500-gene network identified more than 200 functions and revealed ‘focal adhesion’ (KEGG), ‘ECM-related functions’ (KEGG and cluster 0) and ‘PI3K-Akt signaling’ (KEGG) as the functions influencing lithium response. PI3K-Akt signaling has long been implicated in lithium’s action, though the role of focal adhesion and the ECM in lithium response in BD are relatively novel results.

Several limitations temper the interpretation of our results. Most prominent is the small sample size of each iPSC-derived neuron group (6 LR; 5 NR; 6 controls) and the GWAS (*n*=256). Not surprisingly, no significant SNPs in the GWAS were detected due to low power. Only EA subjects were included, limiting generalization to other racial/ethnic groups. Lastly, our iPSC model using very young neurons may not reflect the behavior of mature neurons interacting with other surrounding cells *in vivo* from BD patients.

There are limited reports with which to compare our results. Recently, studies of lithium response in BD have examined gene expression but utilized different tissues or different comparisons (BD vs controls; BD LR vs BD NR; Li-exposure vs non-exposure). The majority of studies utilized lymphocytes^36, 39–41^, whose expression pattern is of limited relevance to brain; while few studies utilized either iPSC derived neurons^18, 42^ or post-mortem brains^43^. In the current study, six of our 41 DE genes from LR vs NR comparisons were also reported in previous transcriptomic studies of lithium response in BD. For five genes (*HEY1*, *KLF10*, *PDGFA*, *POU3F1*, and *PTP4A3*), expression in lymphoblasts was shown to be modulated by lithium^36^. Another gene (*LEF1*) was reported to be responsible for resistance to lithium in NR^42^. Surprisingly, only *PDGFA* appeared to be involved in both focal adhesion and the ECM. Our study failed to detect genes previously reported for lithium response (i.e., *GADL1*^10^*, GRIA2*^8^, *SESTD1*^13^, lncRNAs^11^, and HLA antigen genes^11^). Nonetheless, we performed a *post-hoc* comparison of our 37 DE genes (Li-.LR vs Li-.NR) with genes identified in our PGBD/VA and two other GWA studies: the ConLiGen consortium GWAS of lithium response^9^ (*n*=2563) and the Psychiatric Genomics Consortium Bipolar Disorder Working Group (PGC-BD) GWAS of BD^44^ (*n*=51 710). Of the 35 DE genes for which GWAS results were available, 14 genes showed significant SNP association after Bonferroni correction (12 in our PGBD/VA study; *DSP* and *LMX1B* in the ConLiGen study; *ADAMTS14* and *FOXO6* in the PGC-BD study; **Supplementary Table 17**). *LMX1B* and *ADAMTS14* were significant in two studies. Of these 14 significantly associated genes, 12 were part of the top 500-proximal gene network and achieved statistically significant functional enrichment. Taken together, our *post-hoc* findings indicate a striking level of correspondence between the RNA-seq and three independent GWAS results.

Focal adhesion and the ECM were novel and unexpected findings. In neurons, focal adhesions are a complex of proteins that bind multiple ECM proteins^45–47^. Integrins on the cell surface are activated by mechanical or chemical signals from the ECM, and in turn form the focal adhesion complex, which promotes actin-microtubule polymerization^46, 48, 49^. Axon guidance occurs as filopodia and lamellipodia of the growth cone detect and respond to axon guidance signals from the ECM proteins (known as guidance cues), resulting in growth-cone motility and turning^50–54^. The ECM, comprising various proteins, e.g., collagens and non-collagenous glycoproteins^55^, has been shown to be a dynamic structure that provides not just structural support for neurons and glia cells but has an important role in axon guidance and regulation of axonal growth^56, 57^. Thus, in neurons, focal adhesions and the ECM together form a “motor” that propels growth-cone movement and steering via downstream regulation of actin cytoskeleton organization^47, 49, 52–54, 58, 59^.

**Figure 4e** is the KEGG pathway diagram for focal adhesion (hsa04510), the most enriched among the 17 pathways relevant to BD/neuronal system, annotated with gene involvement from our analyses. Among these multiple genes (*n*=42), of particular interest is *EGFR*, our top hub-like gene, that appears to be essential for neuronal development, including neurite outgrowth and axonal regeneration^60^. A recent study also demonstrates that *EGFR* can modulate integrin tension and focal adhesion formation^61^. Altogether, it supports the role of *EGFR* in the focal adhesion process and the mechanism of lithium response in BD. It also shows an example of biological interconnections contributing to lithium response in BD, which were successfully identified by integrative multi-omics approaches.

A variety of studies indicate a strong effect of lithium on neurons in culture and in animal models^62, 63^. Lithium has been shown at therapeutic concentrations to regulate axon morphology, i.e., promote axon growth, enlarge the growth cone, and increase neurite branching^64, 65^. Inhibition of GSK3β results in a similar phenotype of elongated axons and increased branching consistent with lithium’s action being mediated by its inhibition of GSK3β^66, 67^. Lithium and two other mood stabilizing drugs, carbamazepine and valproate, all prevent growth-cone collapse, increase growth-cone area and axonal branching^68^.

BD neurons are shown to have morphopathological changes in a variety of measures, e.g., reductions in number, size, density, and/or dendrite lengths^69–71^, which suggests that BD neurons tend to be smaller with short dendrites and axons, as compared with normal/healthy neurons. Together with our findings, we hypothesize that BD LR inherit genetic defects in ‘focal adhesion’ and the ECM including the integrin- ECM interactions that cause disruption of focal adhesion function. This results in poorly branched and shortened axons with malformed growth cones that convey susceptibility to BD (**Figure 5c**). Lithium, through its actions (likely upon inhibition of GSK3β) of facilitating branching, re-arborization, and supporting the growth cone, is effective for BD LR by correcting these morphological and functional defects (**Figure 5c’’**). BD NR, on the other hand, have BD for reasons other than dysfunctional focal adhesion, and therefore, lithium does not rescue the relevant mechanisms and they fail to respond (**Figure 5c’’’**). In this model, BD LR and BD NR result from different disease mechanisms as has been previously proposed^5, 18, 19, 36^. Consistent with this, another line of evidence shows a significant difference in cellular adhesion in directly induced neuron-like cells between BD LR than BD NR^72^. This highlights the role of the biological cell adhesion-ECM process in the underlying mechanism of lithium response in BD.

In conclusion, to our knowledge, our study is the first to report the significant role of ‘focal adhesion’ and the ‘ECM’ influencing lithium response in BD. This study also demonstrates the power of applying network methods to multi-omics data. Our results suggest that both genetic and functional studies of lithium response and/or BD should focus efforts on the pathways of focal adhesion and the ECM as well as regulation of axonal growth/extension and synaptic connectivity. Distinguishing two distinct forms of BD would advance our understanding of disease mechanism and facilitate the development of novel therapeutics or a clinical test for lithium response.

## Data Availability

All data produced in the present study are available upon reasonable request to the authors.

## Acknowledgements

We thank the patients who participated. The study was primarily supported by a grant to JRK from the NIMH (U01 MH92758) as part of the Pharmacogenomics Research Network (PGRN) and a grant from the Department of Veterans Affairs. KMF is supported by a grant from the National Center for Advancing Translational Sciences, part of the NIH, as part of support for the UCSD CTSA (UL1TR001442). FHG and MCM are supported by U19MH106434, part of the National Cooperative Reprogrammed Cell Research Groups (NCRCRG) to Study Mental Illness. FHG is also supported by grants from AHA-Allen Initiative in Brain Health and Cognitive Impairment Award made jointly through the American Heart Association and The Paul G. Allen Frontiers Group (19PABH134610000), The JPB Foundation, Bob and Mary Jane Engman, Annette C. Merle-Smith, R01 MH095741, and Lynn and Edward Streim. The Halifax group (MA, CVC, JG, CO, and CS) is supported by grants from Canadian Institutes of Health Research (#166098), ERA PerMed project PLOT-BD, Research Nova Scotia, Genome Atlantic, Nova Scotia Health Authority and Dalhousie Medical Research Foundation (Lindsay Family Fund).

## Author Contributions

**Designed the study:** KEB, JRC, WHC, ESG, SGL, MJM, MGM, DC, JIN, KJØ, PPZ, MA, and JRK.

**Collected the data:** VN, MCM, RS, NA, AA, YB, WHB, HB, KEB, JRC, CVC, CC, WHC, AD, SF, CF, NF, MF, KG, JG, ESG, FG, TG, GH, PJ, M Kamali, M Kelly, SGL, FL, MJM, MGM, DC, CEM, FM, GM, JIN, CO, KJØ, KR, MS, PDS, CS, EKS, AS, BT, PPZ, MA, FHG, and JRK.

**Analysis and interpretation of the data:** VN, SBR, CMN, AXM, DC, PDS, KMF, and JRK.

**Wrote the manuscript:** VN, SBR, CMN, JRC, WHC, ESG, MJM, MGM, DC, JIN, KJØ, PDS, PPZ, MA, KMF, FHG, and JRK.

**Securing funding:** KEB, JRC, WHC, ESG, SGL, MJM, MGM, DC, JIN, KJØ, MA, and JRK.

**Overall principal investigator:** JRK.

All authors have read and approved the final version of the manuscript.

## Ethical Declarations

The Human Research Protection Program of UCSD gave ethical approval for this work.

