## Supplemental Methods for "Network-based integrative analysis of lithium response in bipolar disorder using transcriptomic and GWAS data"

#### Subjects

All subjects in each study provided written informed consent according to procedures approved by their local human subjects committee. The subjects for this study came from three sources: the Pharmacogenomics of Bipolar Disorder study (PGBD), an identical study of Veterans Affairs San Diego Health Care System (VA), and a collection from Dalhouise University (Halifax).

The goal of the Pharmacogenomics of Bipolar Disorder (PGBD) study was to identify genetic variants associated with lithium response in bipolar disorder (BD). The PGBD study included 11 clinical sites in the US, Canada, and Norway. Subjects were required to have a Bipolar I diagnosis and to have had an episode within the last year. The study design has been previously described in detail[1]. Briefly, a longitudinal relapse prevention design was used. Subjects were initially screened and over a 4-month period lithium was started and titrated to 1.0 mEq/L or as close as could be tolerated. Screening included an interview using the Diagnostic Interview for Genetic Studies (DIGS)[2] for standardized diagnosis. Those that were stabilized on lithium defined by a Clinical Global Impression score (CGI) <4 entered the observation phase and were observed for one month on monotherapy before they entered the maintenance phase where they were monitored for relapse over a 2-year period. The VA study was essentially identical to the PGBD save enrolling only veterans, and data were combined for a PGBD/VA sample. The Halifax sample was drawn from a larger set of bipolar I subjects who have been followed clinically for many years and assessed retrospectively using the Alda scale[3, 4].

Subjects for the induced pluripotent stem cells (iPSC) portion of the study were selected from the PGBD/VA, and Halifax samples, subjects were selected for reprogramming from the extremes of response. For the PGBD/VA study, responders advanced to maintenance and were stable for up to 2 years. Non-responders were those who received a good trial and tolerated the medication but did not respond. For the Halifax sample, subjects were selected from the extremes of the distribution of Alda scores. Subjects from the PGBD/VA study, as well as the control group underwent skin biopsies and iPSC lines from fibroblasts were established[5], while lymphoblasts were used to establish iPSC lines for the Halifax subjects[4]. A total of BD patients (*n*=11) and controls (*n*=6) were recruited from the study settings: the PGDB/VA study (BD, *n*=5; controls, *n*=2) and Halifax study (BD, *n*=6; controls, *n*=4). Control subjects were healthy volunteers who were also screened for any history of psychiatric illness using the DIGS interview as with the BD subjects.

### Genome-Wide Association Study (GWAS)

A total of 1106 DNA samples were genotyped with the Human PsychChip when including additional quality control (QC) samples. The PsychChip is a custom version of the Infinium CoreExome-24 v1.1 BeadChip (#WG-331-1111, Illumina, San Diego, CA) that contains additional content from sequencing studies of multiple psychiatric disorders by the Psychiatric Genomics Consortium (PGC). Genotypes were called using GenomeStudio software (Illumina) using the standard self-clustering, and custom clustering files from the Broad Institute developed from a more extensive set of samples to ensure the accuracy of rarer variants (https://sites.google.com/a/broadinstitute.org/psych-chip-resources/home). Self-clustering genotypes were used for initial QC and the Broad provided clusters were used for downstream analysis. Each plate was designed to contain duplicates, a HapMap control sample, and known sex information for all samples to ensure accuracy and quality control. Samples were verified for concentration using Picogreen in triplicate and compared to a standard reference curve, and then normalized to 60 ng/μl with 4 μL used for the assay. Microplates were prepared in batches of two with 8 beadchips per batch, and a total of 588 454 single nucleotide polymorphism (SNP) loci were genotyped. Initial calls identified 7 gender mismatches across all samples, and 14 samples with call rate lower than 0.99.

#### GWAS Analysis

*Genotype Quality Control*

Using SNPs with call rates >95%, subjects were excluded for <98% genotyping rate, excessive heterozygosity ($f_{het}$ <−0.2 or >0.2), or discordance between self-reported sex and sex estimated from inbreeding coefficients calculated from X chromosome SNPs. SNPs were excluded for call rates <98%, Hardy-Weinberg *P*-value <1.0E-06 (among European ancestry subjects), being monomorphic, or significant differences in call rate between plates (*P* <1.0E-08 on one plate or *P* <1.0E-04 on two plates).

*Imputation*

Genotype data was aligned to the same strand as 1000 Genomes Phase 3 data. For ambiguous markers, European ancestry subjects were used to calculate allele frequencies and SNPs with minor allele frequency (MAF) <40% and ≤15% difference in MAF to 1000G Europeans were retained. Phasing was performed without reference population subjects in 3 megabase (MB) blocks with a 1 MB buffer at each block-end using SHAPEIT[6] v2.r837 (https://mathgen.stats.ox.ac.uk/genetics_software/shapeit/shapeit.html). Haplotypes were imputed using IMPUTE2 v2.2.2[7] with 1000 Genomes Phase 3 reference data and genetic map, a 1 MB buffer, and effective population size set to 20,000.

*Phenotype Definition*

The present work employed the "entered_maintenance" phenotype, which describes whether the subject was stabilized on lithium alone in the first 4-month stabilization and observation phases of the study. It is a measure of the short-term response to lithium.

*Analysis*

Analyses were conducted using logistic regression as implemented in PLINK[8]. Age, sex and the first three population principal components were included as covariates. Only subjects of European American ancestry were included in the analysis.

**VEGAS Analysis**

Versatile gene-based association study (VEGAS) genes were identified from GWAS summary statistics, using the VEGAS2 algorithm[9], using the webtool (https://vegas2.qimrberghofer.edu.au/), with default parameters. After mapping SNPs to genes and pruning of SNPs in linkage disequilibrium, VEGAS calculates one empirical *P*-value for each gene.

### GWAS Boosting Analysis

We used a network-based method to boost and reprioritize the marginal hits from the lithium-response GWAS results. The network boosting method we used was GWAB, Genome-Wide Association Boosting[10]. This method employs a Bayesian model to reprioritize marginally significant GWAS genes, using information from a tissue non-specific interactome (HumanNet brain interactome database; http://www.functionalnet.org/humannet/about.html)[11]. GWAB uses a simple method of selecting the SNP with the best *P*-value within 10 kilobases (kb) to map to a gene.

### iPSC Studies

#### Generation of iPSC-Derived Neurons

*Induction of Pluripotency and Differentiation to Hippocampal Neurons*

Reprogramming and differentiation of cells to *prox1*+ hippocampal dentate gyrus granule cells (DG) have been previously described[4, 5]. Briefly, BD and control iPSCs from the PGBD/VA study were derived from fibroblasts and using a Cyto-Tune Sendai reprogramming kit (Invitrogen, Thermo Fisher Scientific, Waltham, MA) according to the manufacturer's instructions. BD and control iPSCs from the PGBD/VA study were derived from skin biopsies and fibroblast cultures, while those from Halifax were derived from Epstein–Barr virus (EBV)-immortalized B-lymphocytes using an episomal vector set described by Okita et al.[12], consisting of three sets of episomal plasmids expressing reprograming factors: pCXLE-hOCT3/4-shp53, pCXLE-hSK (SOX2, KLF4), and pCXLE-hUL (L-MYC, LIN28).

All iPSCs were characterized for pluripotency by pan-differentiation, and karyotyping. iPSC colonies were cultured on Matrigel-coated dishes (BD Biosciences, Franklin Lakes, NJ) using mTeSR1 medium (StemCell Technologies Inc., Cambridge, MA). Mechanical dissociation of iPSC colonies using collagenase and plating onto low-adherence dishes in DMEM/F12 (Invitrogen) supplemented with N2 and B27 was used for formation of embryoid bodies. Floating embryoid bodies were treated with DKK1 (0.5 μg $ml^{-1}$), SB431542 (10 μm), noggin (0.5 μg $ml^{-1}$), and cyclopamine (1 μm) for 20 days. Embryoid bodies were plated onto polyornithine/laminin (Sigma-Aldrich, St. Louis, MO)-coated dishes in DMEM/F12 plus N2 and

B27 to obtain neural progenitor cells. Rosettes were manually collected and dissociated with accutase (Chemicon International, Fisher Scientific, Waltham, MA). After 1 week they were plated onto laminin-coated dishes in neural progenitor cell media (DMEM/F12, 1×N2, 1×B27 (Invitrogen), 1 μg $ml^{-1}$ laminin, and 20 ng $ml^{-1}$ FGF2 (Invitrogen). Neural progenitor cells were then differentiated in DMEM/F12 supplemented with 1×N2, 1×B27, 20 ng $ml^{-1}$ BDNF (PeproTech, Cranbury, NJ), 1 mM dibutyrl-cyclicAMP (Sigma-Aldrich), 200 nM ascorbic acid (Sigma-Aldrich), 1 μg $ml^{-1}$ laminin, and 620 ng $ml^{-1}$ Wnt3a (R&D Systems, Minneapolis, MN) for 3 weeks. Wnt3a was removed after 3 weeks. All cells used in the present study were verified as free from mycoplasma contamination.

*Lithium Treatment of iPSC-Derived Neurons*

iPSC-derived neurons from both clinically validated BD lithium responders (LR) and BD non-responders (NR) were treated in culture both with (lithium treated) and without lithium (untreated). Cells were treated for one week at 1 mM, a clinically effective blood concentration.

**RNA-Sequencing of iPSC-Derived Neurons**

*RNA-Sequencing Process*

Cells were transfected with a lentivirus reporter construct *prox1*::GFP, in order to identify the DG neurons. The reporter was used to distinguish *prox1*+ DG neurons for fluorescence activated cell sorting (FACS) in order to sort the cells into a pure population of DG neurons. After one week of rest, RNA-sequencing (RNA-seq) libraries were prepared using an Illumina TruSeq Stranded Total RNA Sample Prep Kit (Illumina). SuperScript™ II reverse transcriptase (Invitrogen) was used to reverse transcribe the depleted RNA into cDNA. Stranded cDNA sequencing libraries were generated and sequenced paired-end 2×100 base pairs (bp) using the Illumina HiSeq 2500 platform according to the manufacturer's specifications.

*RNA-Seq Analysis*

Quality control of the raw fastq files was performed using the software tool FastQC[13] (https://www.bioinformatics.babraham.ac.uk/projects/fastqc/). Sequencing reads were aligned to the human genome (hg19) using the STAR v2.5.1a aligner[14] (https://github.com/alexdobin/STAR). Read quantification was performed with RSEM[15] v1.3.0 (https://github.com/deweylab/RSEM) and Ensembl annotation (GENCODE Human v19; https://www.gencodegenes.org/). The R BioConductor packages edgeR[16] and limma[17] were used to implement the limma-voom[18] method for differential expression analysis. Lowly expressed genes were filtered out (counts-per-million [CPM] >1 in at least one sample). Trimmed mean of M-values (TMM)[19] normalization was applied. The experimental design was modeled upon time point and treatment (~0 +condition_treatment + tissue). The lmFit function in limma with consensus correlation to account for repeated measures of patient followed by the eBayes function was used to fit the design on voom normalized counts per gene. Significance was defined by using an adjusted *P*-value cut-off of 0.05 after multiple testing correction using a moderated t-statistic in limma.

*Comparisons of RNA-Seq Analysis of iPSC-Derived Neurons*

We applied two strategies in the comparisons of RNA-seq analysis of iPSC-derived neurons as follows and illustrated in **Supplemental Methods Figure 1**:

1. ***Within-group analysis***. We compared the gene expression in neurons from the same group of subjects between under lithium treated (Li+) and untreated (Li-) conditions to see the effects of lithium on each of sample groups (LR, NR, and controls). A total of three comparisons were made.

2. ***Across-group analysis***. We compared the gene expression in neurons from the different group of subjects under the same treatment conditions (either lithium treated [Li+] or untreated [Li-]) to see the effects of lithium on the three different sample groups (among LR, NR, and controls). A total of six comparisons were made.

3. In addition, the interaction between (LR vs NR) and (Li+ vs Li-) was also evaluated.

**Reverse Transcription Quantitative Real-Time PCR (RT-qPCR) Validation**

Briefly, one µg of total RNA from seven patient-derived iPSC neurons (LR, *n*=2; NR, *n*=3; controls, *n*=2) was quantified by a NanoDrop™ 2000 (Thermo Fisher Scientific, Waltham, MA), and then reversed-transcribed to generate cDNAs using Invitrogen™ SuperScript™ III

Reverse Transcriptase (Invitrogen). Each of a 20 µl-RT-qPCR reaction mix was prepared following to the manufacturer's instructions, consisting of a cDNA, a TaqMan® Gene Master Mix (Thermo Fisher Scientific), and a TaqMan® Gene Expression Assay (Thermo Fisher Scientific). TaqMan® Gene Expression Assays were a set of probes and primers targeting four genes of interest: *HEY1* (Hs01114113_m1), *KLF10* (Hs00921811_m1), *POU3F1* (Hs00538614_s1), and *PTP4A3* (Hs02341135_m1). *HPRT1* gene was used as an endogenous control assay (Hs99999909_m1, Thermo Fisher Scientific). RT-qPCR reactions of each sample for each gene were amplified and performed in triplicate in the CFX Connect™ Real-Time PCR Detection System (Life Science, Hercules, CA). Cycling conditions were conducted following the manufacturer’s protocol. RT-qPCR data was acquired by the CFX Manager™ Software (Life Science). Relative quantification was analyzed using the delta delta Ct method.

**Preliminary Functional Enrichment Analysis**

In this study, the RNA-seq genes that reached the *P*-value <0.05, $\log_2$fold-change ≥|1|, and Benjamini-Hochberg (B-H) false discovery rate (FDR) (B-H *q*-value) ≤0.20 based on the results of RNA-seq analysis are considered as significantly differentially expressed genes, called ‘DE genes’ (total *n*=45; **Figures 2c**, **d**; **Supplementary Table 8**).

We performed the preliminary functional enrichment analysis of the 41 protein-coding DE genes, using the WebGestalt[20] and the g:Profiler[21] web tools. WebGestalt version 2017 (http://www.webgestalt.org/2017/option.php#) was used to carry out the overrepresentation enrichment analysis (OSA) of the 41 DE genes in KEGG pathways, using human genome protein coding as background. While g:Profiler version r1760_e93_eg40 (https://biit.cs.ut.ee/gprofiler_archive2/r1760_e93_eg40/web/) was used to perform the enrichment analysis of the 41 DE genes in several functional domains (i.e., gene ontology, biological pathways, regulatory motifs, proteins, and human phenotype ontology), using all genes annotated in the Ensembl database as a background. The B-H adjusted FDR was used in these analyses for a multiple testing correction in both tools using a different threshold for significance. The enrichment threshold of the nominal *P*-value <0.05 and B-H *FDR* <0.05 was considered statistically significant. Those with B-H *FDR* <1.0E-05 were considered strongly significant.

### Network Propagation and Functional Enrichment Analyses

#### Network Propagation Analysis and Hypergeometric Testing

We used network propagation[22] to find associations between the 41 protein-coding DE genes from RNA-seq with network-boosted GWAS results. Network propagation provides information about the local neighborhood of a set of 'seed' genes in network space, and genes that have high network propagation scores are likely to be related to these 'seed' genes. We used the GIANT brain interactome[23] (Tissue-specific gene networks from HumanBase; https://hb.flatironinstitute.org/about) as the basis for propagation (filtering on edges with weight >0.2). Network propagation and visualization were conducted using visJS2jupyter[24] and Cytoscape[25]. Both the top 500- and top 2000-proximal gene networks were generated.

A hypergeometric test was used to compare the overlap in gene sets between the top 500 or 2000 network genes derived from DE genes, and the top 5% of prioritized genes identified by each of GWAB ($n$=1119) or VEGAS ($n$=1180). All genes in the interactome were used as the background set for this calculation.

#### Functional Enrichment Analysis

*Cluster Analysis*

Clusters in the top 500-gene proximal subgraph were identified using the Louvain modularity maximization algorithm[26]. The algorithm identifies groups of genes which have more interconnections between cluster genes than between non-cluster genes. Three clusters were identified in this study **Figures 2e**, **3**; **Supplementary Table 10**).

Functional enrichment analysis was performed on the 500-gene proximal subgraph using the ToppGene analysis suite[27], and the g:Profiler tool[21], using all protein-coding genes as the background set.

Functional enrichment analysis was performed on each of the three clusters identified in this study, with the 500 genes in the proximal network as the background set.

*KEGG Pathway Analysis*

Significant results from this enrichment analysis were filtered to focus on KEGG (Kyoto Encyclopedia of Genes and Genomes) pathways (https://www.genome.jp/kegg/pathway.html), which contained at least one seed gene and one GWAB gene, with the full set of protein coding genes as the background set. This resulted in 37 significantly enriched KEGG pathways (**Figures 4a**, **b**; **Supplementary Figure 7**; **Supplementary Table 15**). The connectivity degree of the 500 proximal network genes was assessed by measuring the number of interacting neighbors in the network. Genes with a high degree of connectivity were considered as 'hub-like' genes.

*Pathview Creation with Degree of Proximity*

The Pathview tool[28] in R was used to create KEGG pathway maps. Genes in the 500-proximal gene network were color-coded by the network proximity value.

### ***Post-Hoc* Comparison Analysis**

To see whether significant DE genes presented in GWAS, we searched for our 37 DE genes (Li-.LR vs Li-.NR; **Figures 1c**, **d**; **Supplementary Table 8a**) from the iPSC-derived neuron RNA-seq in our PGBD/VA study (*n*=256) and additional GWAS data available from two studies: the ConLiGen study of lithium response[29] (*n*=2563) and the Psychiatric Genomics Consortium Bipolar Disorder Working Group study of genes for BD[30] (PGC-BD2 dataset; *n*=51 710). Out of 37 DE genes, 35 were interrogated among three GWAS data sets (**Supplementary Table 17**) and used in a *post-hoc* comparison analysis for significance of the 35 DE genes in the PGBD and other two GWAS. Though no individual SNP was genome-wide significant in each GWAS, in this comparison analysis only 105 tests were conducted for a Bonferroni corrected significance threshold of 4.76E-04, which was met by 14 genes in the three GWAS.

**Supplemental Methods Figure 1: The comparison scheme of RNA-sequencing analysis of iPSC-derived neurons.**

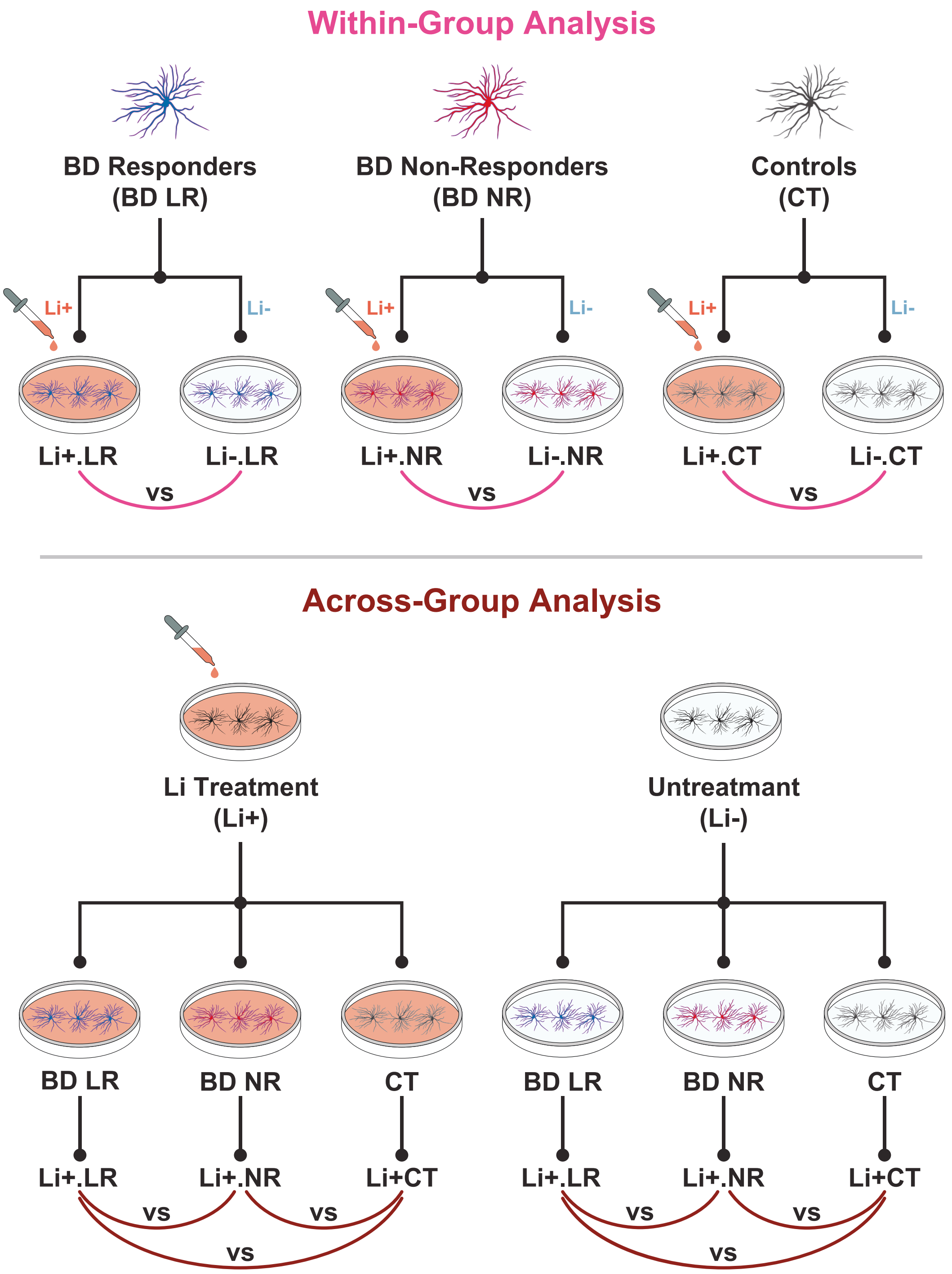

The illustration shows the pairwise comparisons (total *n*=9) obtained in the RNA-seq analysis of iPSC neurons by applying the two strategies as follows: (**i**) 'Within-group' analysis (*top* panel) by comparing in each group of subjects (LR, NR, or CT) under different treatments (between Li+ and Li-), resulting in three group comparisons. (**ii**) 'Across-group' analysis (*bottom* panel) by comparing among the different group of subjects (LR, NR, and CT) under the same treatments (either Li+ or Li-), resulting in six group comparisons. The interaction between clinical response and *in vitro* treatment was also examined (*not* shown in illustration).

BD, Bipolar disorder; CT, controls; iPSC, induced pluripotent stem cells; Li+, Li-treated; Li-, untreated; LR, BD responders; NR, BD non-responders; RNA-seq, RNA-sequencing.
