## Supplementary Figures for "Network-based integrative analysis of lithium response in bipolar disorder using transcriptomic and GWAS data"

### SUPPLEMENTARY FIGURES 1-7 FOR:

### Network-Based Integrative Analysis of Lithium Response in Bipolar Disorder Using Transcriptomic and GWAS Data

Vipavee Niemsiri[1], Sarah Brin Rosenthal[2], Caroline M. Nievergelt[1], Adam X. Maihofer[1], Maria C. Marchetto[3,4], Renata Santos[4,5], Tatyana Shekhtman[1], Ney Alliey-Rodriguez[6,7], Amit Anand[8,9], Yokesh Balaraman[10], Wade H. Berrettini[11], Holli Bertram[12], Katherine E. Burdick[9], Joseph R. Calabrese[13,14], Cynthia V. Calkin[15], Carla Conroy[13,14], William H. Coryell[16], Anna DeModena[17], Scott Feeder[18], Carrie Fisher[10], Nicole Frazier[12], Mark A. Frye[18], Keming Gao[13,14], Julie Garnham[19], Elliot S. Gershon[6], Fernando Goes[20], Toyomi Goto[13], Gloria J. Harrington[12], Petter Jakobsen[21], Masoud Kamali[8,12], Marisa Kelly[12], Susan G. Leckband[17], Falk Lohoff[11], Michael J. McCarthy[1], Melvin G. McInnis[12], David Craig[22], Caitlin E. Millett[9], Francis Mondimore[20], Gunnar Morken[23], John I. Nurnberger[10,24], Claire O' Donovan[19], Ketil J. Øedegaard[21], Kelly Ryan[12], Martha Schinagle[13], Paul D. Shilling[1], Claire Slaney[19], Emma K. Stapp[25], Andrea Stautland[26], Bruce Tarwater[16], Peter P. Zandi[20], Martin Alda[19,27], Kathleen M. Fisch[2,28], Fred H. Gage[4], John R. Kelsoe[1,29]

[1] Department of Psychiatry, University of California, San Diego, La Jolla, CA, USA
[2] Center for Computational Biology and Bioinformatics, University of California, San Diego, La Jolla, CA, USA
[3] Department of Anthropology, University of California, San Diego, La Jolla, CA, USA
[4] Laboratory of Genetics, Salk Institute for Biological Studies, La Jolla, CA, USA
[5] University of Paris, Institute of Psychiatry and Neuroscience of Paris (IPNP), INSERM U1261266, Laboratory of Dynamics of Neuronal Structure in Health and Disease, Paris, France
[6] Department of Psychiatry and Behavioral Neuroscience, University of Chicago, Chicago, IL, USA
[7] Department of Psychiatry and Behavioral Neuroscience, Northwestern University, Chicago, IL, USA
[8] Department of Psychiatry, Massachusetts General Hospital, Harvard Medical School, Boston, MA, USA
[9] Department of Psychiatry, Brigham and Women's Hospital, Harvard Medical School, Boston, MA, USA
[10] Department of Psychiatry, Indiana University School of Medicine, Indianapolis, IN, USA
[11] Department of Psychiatry, University of Pennsylvania, Philadelphia, PA, USA
[12] Department of Psychiatry, University of Michigan, Ann Arbor, MI, USA
[13] Mood Disorders Program, Case Western Reserve University School of Medicine, Cleveland, OH, USA
[14] Mood Disorders Program, University Hospitals Cleveland Medical Center, Cleveland, OH, USA
[15] Department of Psychiatry and Medical Neuroscience, Dalhousie University, Halifax, Nova Scotia, Canada
[16] Department of Psychiatry, University of Iowa, Iowa City, IA, USA
[17] Psychiatry Service, VA San Diego Healthcare System, San Diego, CA, USA
[18] Department of Psychiatry, The Mayo Clinic, Rochester, MN, USA
[19] Department of Psychiatry, Dalhousie University, Halifax, Nova Scotia, Canada
[20] Department of Psychiatry and Behavioral Sciences, Johns Hopkins University, Baltimore, MD, USA
[21] Norment, Division of Psychiatry, Haukeland University Hospital and Department of Clinical medicine, University of Bergen, Bergen, Norway
[22] Department of Translational Genomics, University of Southern California, Los Angeles, CA, USA
[23] Division of Mental Health Care, St Olavs University Hospital, and Department of Mental Health, Norwegian University of Science and Technology – NTNU, Trondheim, Norway
[24] Medical and Molecular Genetics, Stark Neurosciences Research Institute, Indiana University School of Medicine, Indianapolis, IN, USA
[25] Division of Psychiatry, Faculty of Medicine and Dentistry, Stavanger University Hospital, University of Bergen, Stavanger, Norway
[25] Department of Mental Health, Johns Hopkins Bloomberg School of Public Health, Johns Hopkins University, Baltimore, MD, USA
[26] Department of Clinical Medicine, University of Bergen, Bergen, Norway
[27] National Institute of Mental Health, Klecany, Czech Republic
[28] Department of Obstetrics, Gynecology & Reproductive Sciences, University of California, San Diego, La Jolla, CA, USA
[29] Institute for Genomic Medicine, University of California, San Diego, La Jolla, CA, USA

**Supplementary Figure 1: Result of genome-wide association analysis for lithium response.**

**a**

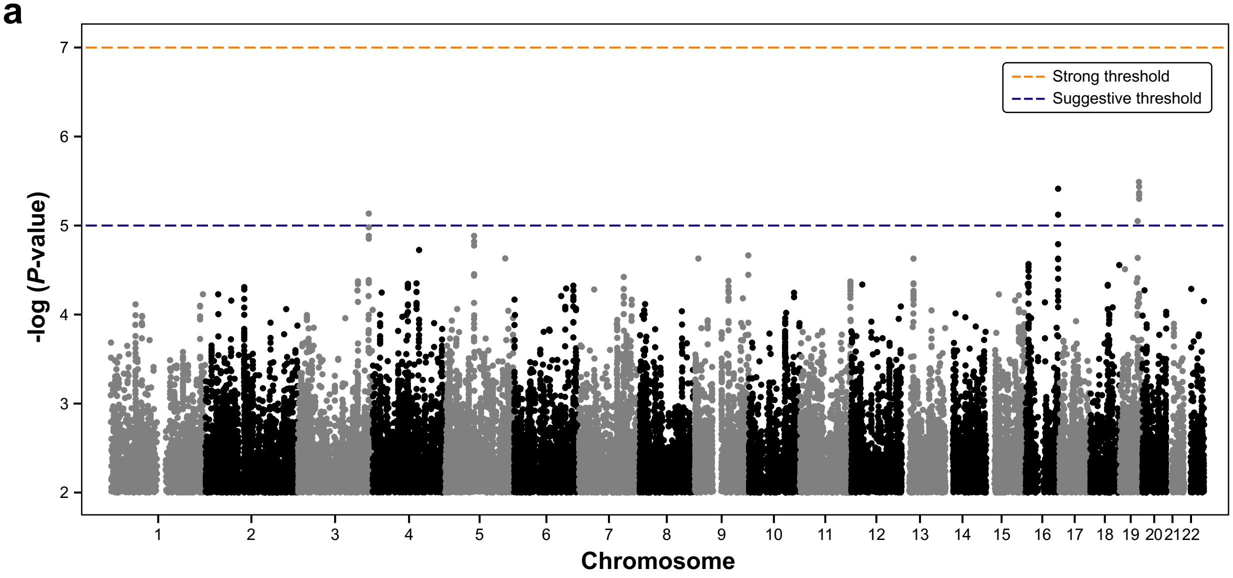

**b**

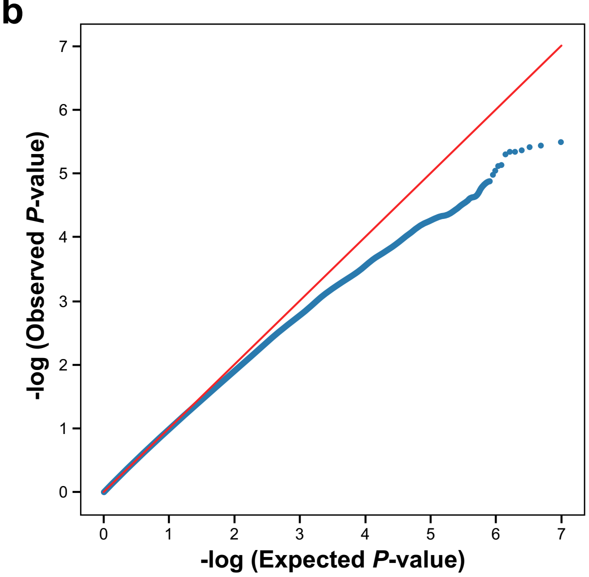

In our genome-wide association analysis, a total of 256 BD patients were obtained from the PGBD[1]/VA study. All were European American. Characteristics of 256 BD patients are described in **Supplementary Table 1**. See additional details of the PGBD/VA study in **Supplemental Methods**.

**1a, Manhattan plot.**
The Manhattan plot shows the distribution of -log *P*-values against chromosomes. No single-nucleotide polymorphism reached genome-wide significance (*P*=5.0E-08). The orange dashed line indicates a genome-wide strong threshold (*P*=1.0E-07). The blue dashed line indicates the genome-wide suggestive threshold (*P*=1.0E-05).

**1b, Quantile-quantile (Q-Q) plot.**
The Q-Q plot represents the -log of *P*-values for the observed distribution (x-axis) against the expected distribution (y-axis). It shows the observed *P*-values did significantly deviate from the expected distribution, indicating an underpowered study.

**Supplementary Figure 2: Metrics for RNA-sequencing validation of iPSC-derived neuron samples.**

a

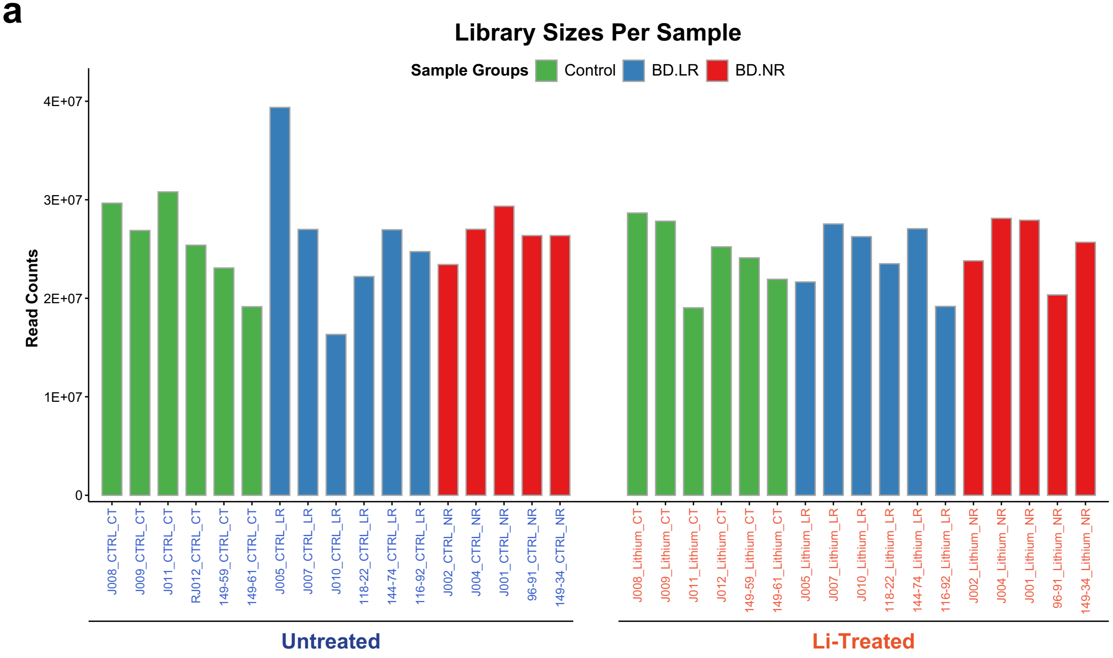

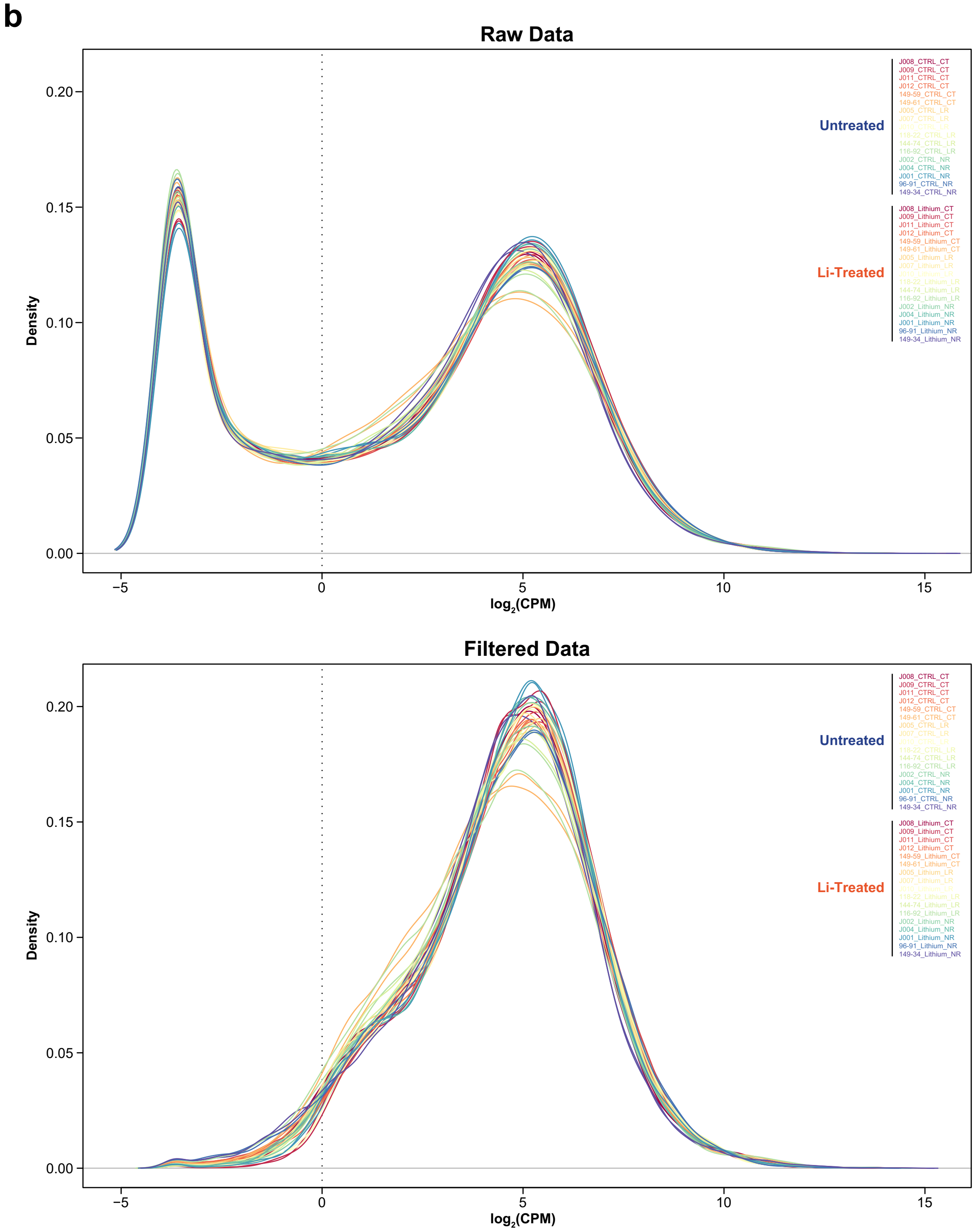
b
Raw Data
Density
log2(CPM)
0.00
0.05
0.10
0.15
0.20
−5
0
5
10
15
Untreated
J008_CTRL_CT
J009_CTRL_CT
J011_CTRL_CT
J012_CTRL_CT
149-59_CTRL_CT
149-61_CTRL_CT
J005_CTRL_LR
J007_CTRL_LR
J010_CTRL_LR
118-22_CTRL_LR
144-74_CTRL_LR
116-92_CTRL_LR
J002_CTRL_NR
J004_CTRL_NR
J001_CTRL_NR
96-91_CTRL_NR
149-34_CTRL_NR
Li-Treated
J008_Lithium_CT
J009_Lithium_CT
J011_Lithium_CT
J012_Lithium_CT
149-59_Lithium_CT
149-61_Lithium_CT
J005_Lithium_LR
J007_Lithium_LR
J010_Lithium_LR
118-22_Lithium_LR
144-74_Lithium_LR
116-92_Lithium_LR
J002_Lithium_NR
J004_Lithium_NR
J001_Lithium_NR
96-91_Lithium_NR
149-34_Lithium_NR
Filtered Data
Density
log2(CPM)
0.00
0.05
0.10
0.15
0.20
−5
0
5
10
15
Untreated
J008_CTRL_CT
J009_CTRL_CT
J011_CTRL_CT
J012_CTRL_CT
149-59_CTRL_CT
149-61_CTRL_CT
J005_CTRL_LR
J007_CTRL_LR
J010_CTRL_LR
118-22_CTRL_LR
144-74_CTRL_LR
116-92_CTRL_LR
J002_CTRL_NR
J004_CTRL_NR
J001_CTRL_NR
96-91_CTRL_NR
149-34_CTRL_NR
Li-Treated
J008_Lithium_CT
J009_Lithium_CT
J011_Lithium_CT
J012_Lithium_CT
149-59_Lithium_CT
149-61_Lithium_CT
J005_Lithium_LR
J007_Lithium_LR
J010_Lithium_LR
118-22_Lithium_LR
144-74_Lithium_LR
116-92_Lithium_LR
J002_Lithium_NR
J004_Lithium_NR
J001_Lithium_NR
96-91_Lithium_NR
149-34_Lithium_NR

**c**

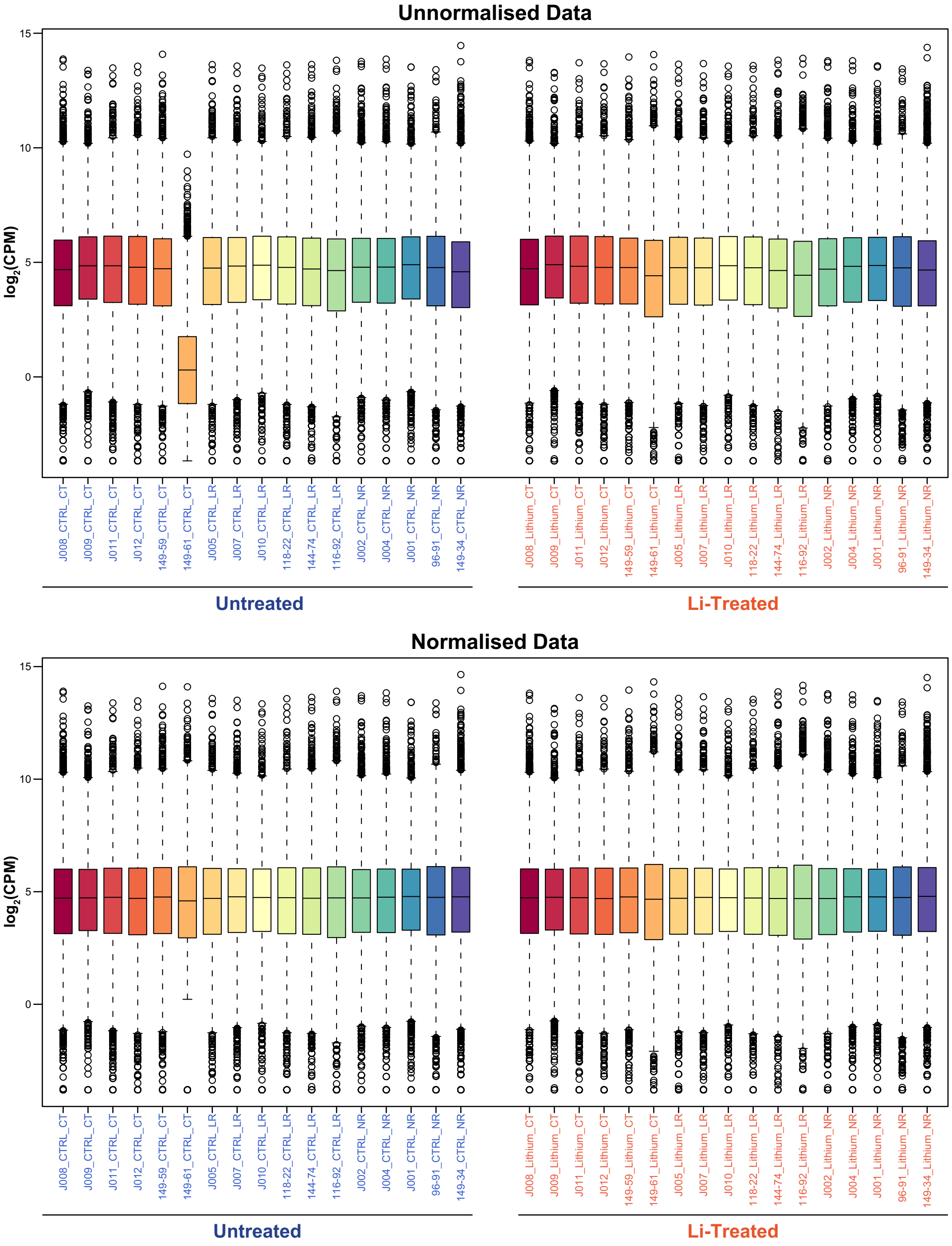
Unnormalised Data
Normalised Data
log2(CPM)
15
10
5
0
J008_CTRL_CT
J009_CTRL_CT
J011_CTRL_CT
J012_CTRL_CT
149-59_CTRL_CT
149-61_CTRL_CT
J005_CTRL_LR
J007_CTRL_LR
J010_CTRL_LR
118-22_CTRL_LR
144-74_CTRL_LR
116-92_CTRL_LR
J002_CTRL_NR
J004_CTRL_NR
J001_CTRL_NR
96-91_CTRL_NR
149-34_CTRL_NR
J008_Lithium_CT
J009_Lithium_CT
J011_Lithium_CT
J012_Lithium_CT
149-59_Lithium_CT
149-61_Lithium_CT
J005_Lithium_LR
J007_Lithium_LR
J010_Lithium_LR
118-22_Lithium_LR
144-74_Lithium_LR
116-92_Lithium_LR
J002_Lithium_NR
J004_Lithium_NR
J001_Lithium_NR
96-91_Lithium_NR
149-34_Lithium_NR
Untreated
Li-Treated

**d**

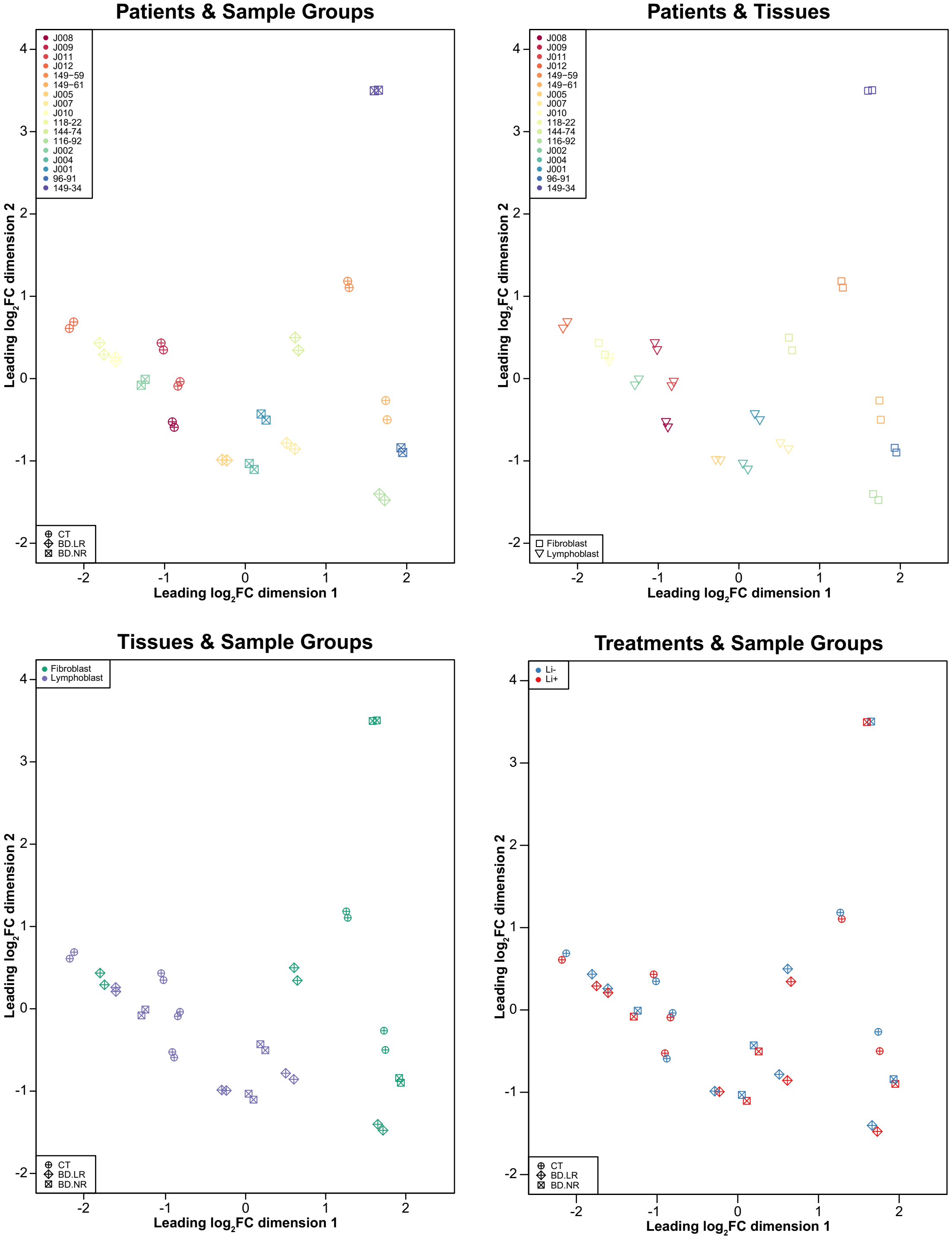

Patients & Sample Groups
J008
J009
J011
J012
149−59
149−61
J005
J007
J010
118-22
144-74
116-92
J002
J004
J001
96-91
149-34
CT
BD.LR
BD.NR
Leading log2FC dimension 1
Leading log2FC dimension 2
Patients & Tissues
Fibroblast
Lymphoblast
Tissues & Sample Groups
Treatments & Sample Groups
Li-
Li+

e

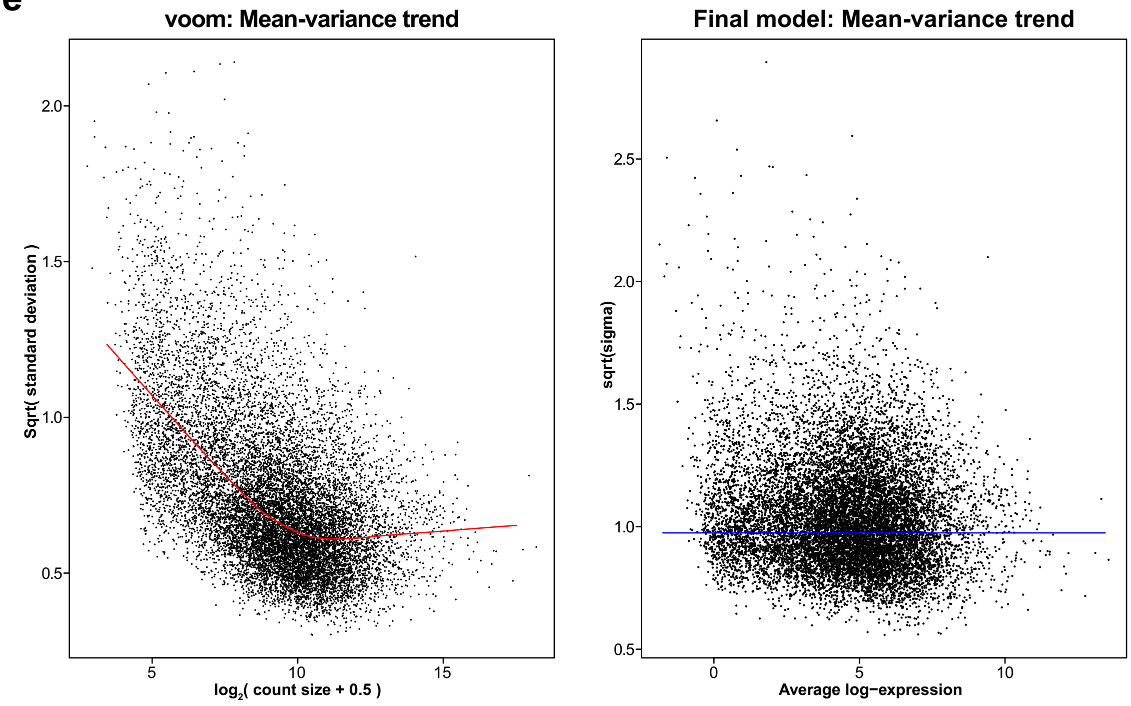

The iPSC-derived neurons were generated from a total 17 patients categorized into 3 subgroups: controls (*n*=6), BD responders (*n*=6), and BD non-responders (*n*=5) under two *in vitro* treatment conditions: untreated (Li-) and lithium-treated (Li+). Characteristics of patients (total *n*=17) used for the iPSC-derived neurons are described in **Supplementary Table 2**. Additional information on subjects-derived iPSC neurons and RNA-sequencing analysis are also provided in **Supplemental Methods**.

**2a, Bar plot of library sizes for each sample.**
Bar plot of the library sizes shows the numbers of sequence mapped reads for each sample. Library sizes represent the sequence read counts after normalization (set to 1), displayed in numbers on the y-axis. Each bar represents the read counts for each iPSC neuron sample, which are present on the x-axis. Bars are grouped by treatments (untreated, *left* panel; Li-treated, *right* panel) and colored by sample groups (controls, green; BD responders, blue; BD non-responders, red). These sequence reads were mapped to the human genome (hg19) assembly with STAR v2.5.1a aligner[2] (https://github.com/alexdobin/STAR) and quantified with RSEM[3] v1.3.0 (https://github.com/deweylab/RSEM) mapped to GENCODE Human v19 (https://www.gencodegenes.org/).

**2b, Density plots of the per sample gene expression distribution.**
Density plots show the distribution of read counts for iPSC-derived neurons from each subject under two *in vitro* treatment conditions. Each sample set was produced from raw pre-filtered data (*top* panel) and post-filtered data (*bottom* panel). The x-axis represents the number of mapped read counts in $log_2$CPM unit. The y-axis represents the density

of mapped read counts. A filtering threshold for removing unexpressed or lowly-expressed genes is set at a CPM value of 1 shown as the dotted vertical line on the x-axis at the $log_2$CPM of 0.

**2c, Box plots of the per sample gene expression distribution.**
Box plots show the read count distribution of each iPSC-derived neuron sample for the unnormalized data (*top* panel) and the Trimmed Mean of *M*-values (TMM)[4] normalized data (*bottom* panel). The x-axis represents each iPSC neuron under two *in vitro* treatment conditions. The y-axis represents the number of mapped reads in $log_2$CPM unit.

For **Figures 2b** and **c**, each sample is specified by colors and classified by treatment conditions.

**2d, Multi-dimensional scaling (MDS) plots of gene expression data.**
MDS plots show the fold-change (in $log_2$FC unit) values over dimension 1 (on the x-axis) and dimension 2 (on the y-axis) for the iPSC-derived neurons. The MDS plots demonstrate a relationship among samples showing three clustering groups of patients (controls; BD responder; BD non-responder). Each point (total *n*=34). represents an individual data of each patient (*n*=17) who was obtained tissues (fibroblasts or lymphoblasts) for generating iPSC-derived neurons, which then were tested under two *in vitro* treatments (untreated, Li-; Li-treated, Li+).

MDS subplots exhibit expression data for different variability settings as follows:
(**i**) *top left*, patients (in colors) and sample groups (in shapes).
(**ii**) *top right*, patients (in colors) and obtained tissues (in shapes).
(**iii**) *bottom left*, obtained tissues (in colors) and sample groups (in shapes).
(**iv**) *bottom right*, *in vitro* treatments (in colors) and sample groups (in shapes).
Distances on the plot correspond to the leading $log_2$FC, which is the average (root-mean-square) $log_2$FC for the 500 genes most divergent between each pair of samples.

**2e, Mean-variance relationships of gene expression data.**
Mean-variance relationships represent the relationship between means and variances of genes in expression data. Each point represents the mean-variance relationship of each gene before (**i**) and after (**ii**) voom precision weights[5] are applied to the data.

(**i**) **Voom model** (*left*).
Voom model is the voom trend that plots variances in square-root standard deviations on the y-axis against mean expression as average $log_2$(count size + 0.5) on the x-axis. Count size corresponds to CPM. The estimated expression trend displays as a red curve line.

(**ii**) **Final model** (*right*).
The final model is the SA plot that plots residual variances in square-root residual standard deviations (or sigma) on the y-axis against mean expression in average log-

CPM values on the x-axis. Sigma is the estimated residual standard deviation. The average $log_2$residual standard deviation displays as a horizontal blue line.

BD.LR, BD Li responders; BD.NR, BD Li non-responders; CPM, counts-per-million; CT, controls; FC, fold-change.

**Supplementary Figure 3: Heatmaps of gene expression of the top 50 RNA-sequencing genes (*P* <0.05).**

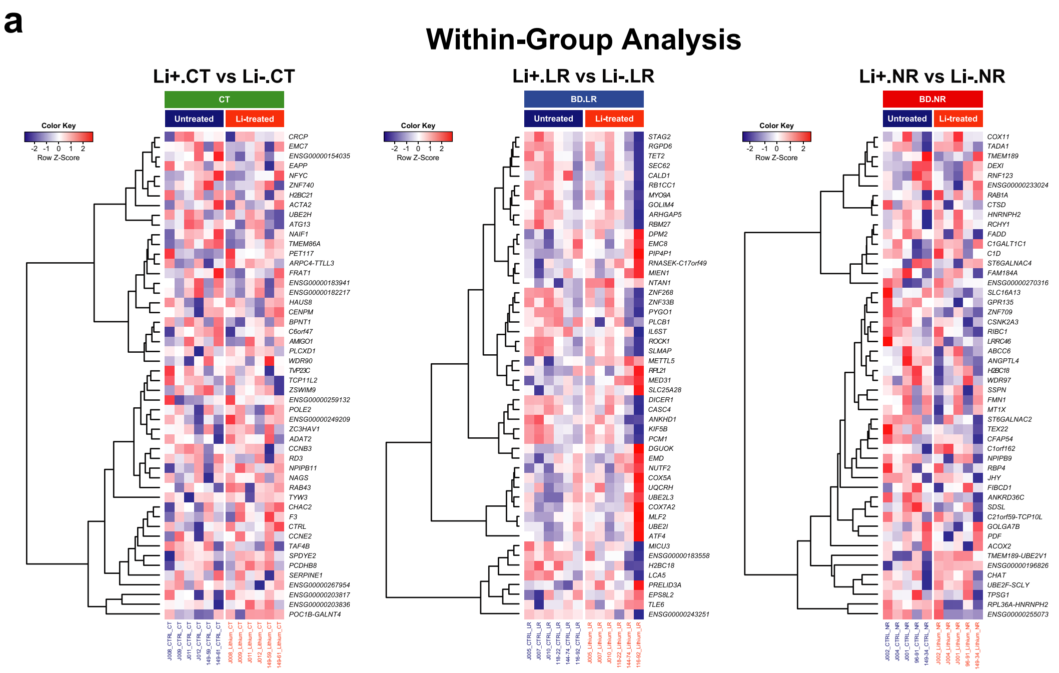

b-1

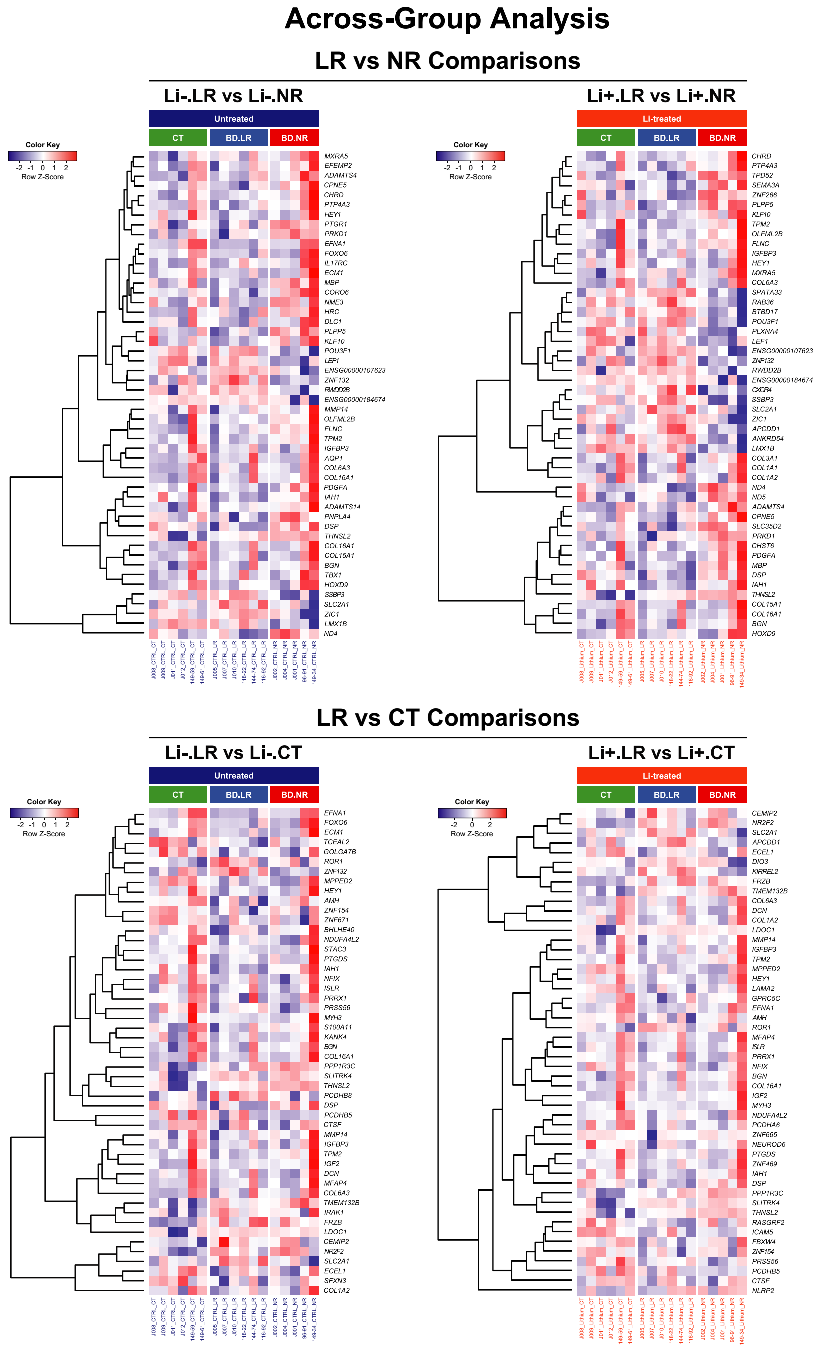
Across-Group Analysis
LR vs NR Comparisons
Li-.LR vs Li-.NR
Li+.LR vs Li+.NR
LR vs CT Comparisons
Li-.LR vs Li-.CT
Li+.LR vs Li+.CT
Untreated
Li-treated
CT
BD.LR
BD.NR
Color Key
Row Z-Score

**b-2**

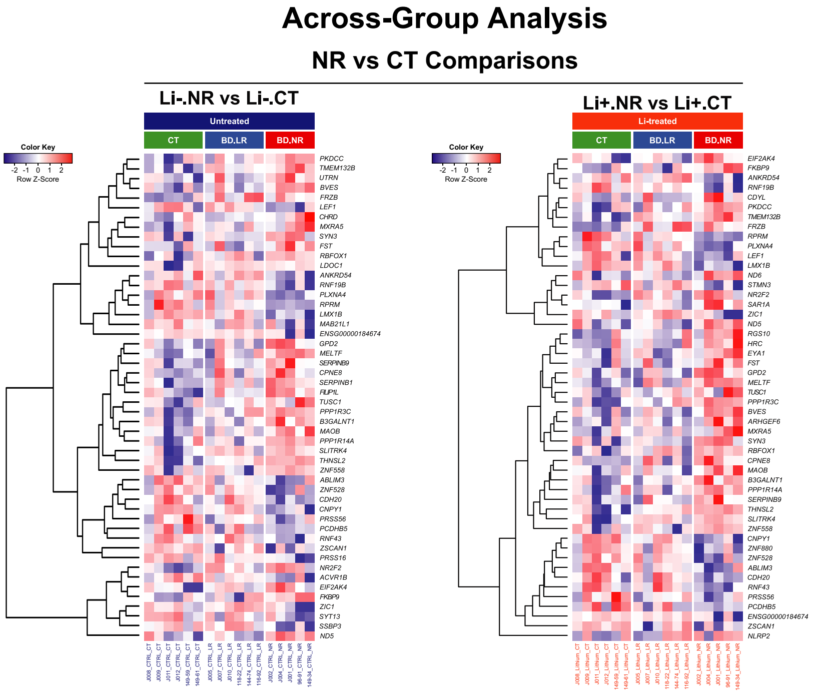

**c**

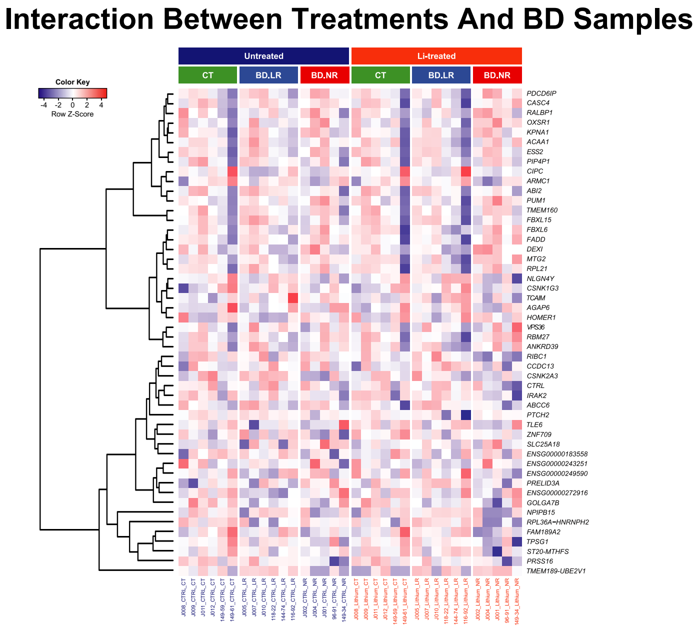

Heatmaps display hierarchical clustering of gene expression levels for the top 50 RNA-seq genes from each comparison in the within-group analysis (**a**), across-group analysis (**b-1** and **2**), and interaction between treatments (untreated [Li-] and Li-treated [Li+]) and

BD samples (BD.LR and BD.NR) (**c**). The color scale (*top left*) represents the degree (Z scores) of differential expression (low, blue; high, red). Gene symbols (rows) are listed. The color boxes above the heatmaps represent sample groups (CT, green; BD.LR, blue; BD.NR, red) and are colored by treatments (Li-, dark blue; Li+, orange). Texts (columns) below the heatmap represent samples and are colored by treatment conditions. Details in group comparisons are described in **Supplemental Methods**. Results of RNA-seq analysis are summarized in **Figures 1b-d** and **Supplementary Tables 4-8**.

**3a,** The top 50 RNA-seq genes ($P$ <0.05) in the within-group analysis:
(**i**) Li+.CT vs Li-.CT (*left*).
(**ii**) Li+.LR vs Li-.LR (*middle*).
(**iii**) Li+.NR vs Li-.NR (*right*).

**3b,** The top 50 RNA-seq genes ($P$ <0.05) in the across-group analysis:
(**i**) LR vs NR comparisons (**b-1**, *top*): Li-.LR vs Li-.NR and Li+. LR vs Li+.NR.
(**ii**) LR vs CT comparisons (**b-1**, *bottom*): Li-.LR vs Li-.CT and Li+.LR vs Li+.CT.
(**iii**) NR vs CT comparisons (**b-2**): Li-.NR vs Li-.CT and Li+.NR vs Li+.CT.

**3c,** The top 50 RNA-seq genes ($P$ <0.05) in the interaction between treatments (Li+ vs Li-) and BD samples (BD.LR vs BD.NR).

CT, controls; LR, BD Li responders; NR, BD Li non-responders; RNA-seq, RNA-sequencing.

**Supplementary Figure 4: Relative gene expression of four selected genes by RT-qPCR.**

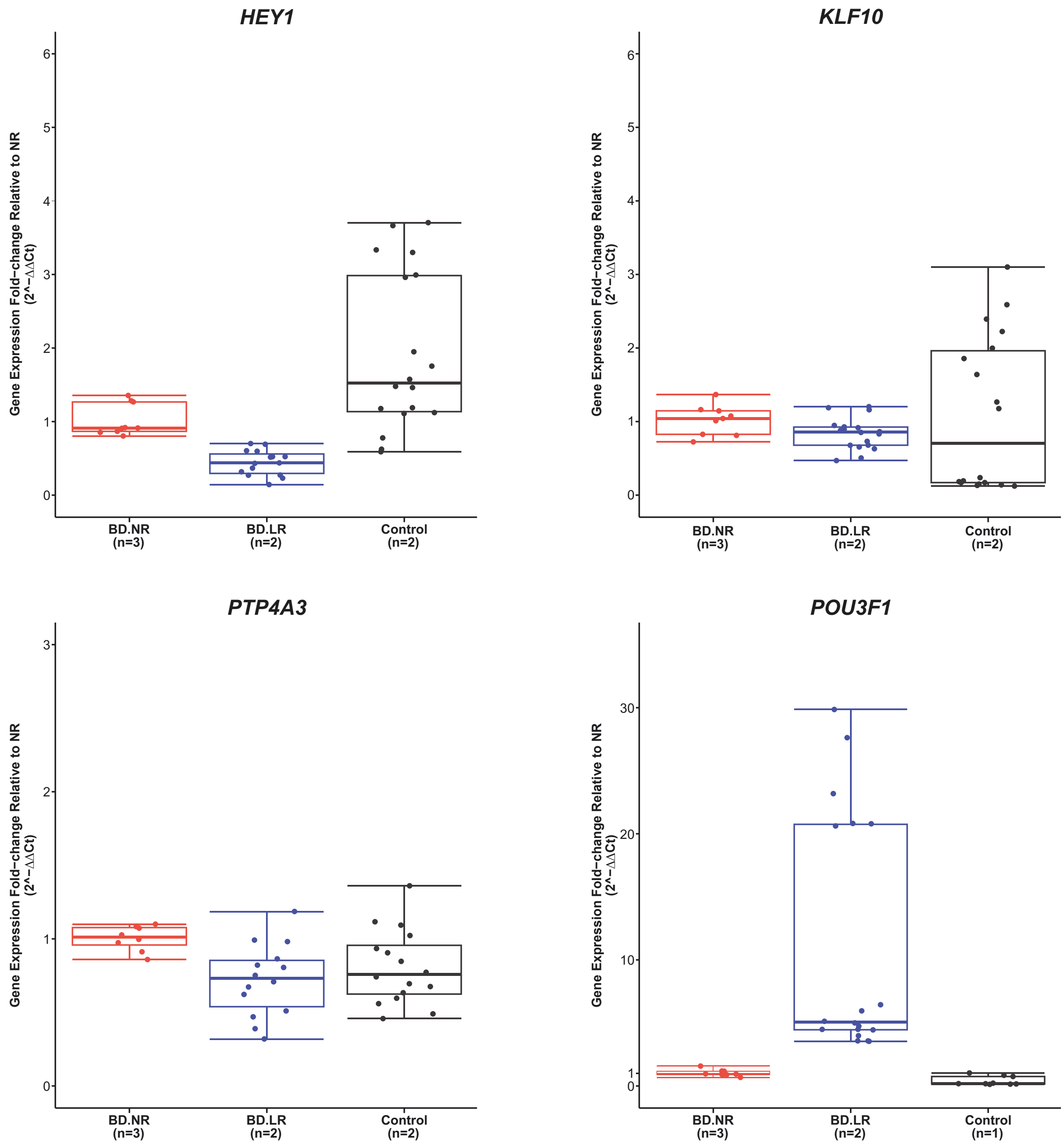

Relative gene expression of four selected differentially expressed (DE) genes *in vitro* untreated (Li-) iPSC-derived neurons of BD LR and CT compared to BD NR, validated by RT-qPCR. These four genes were selected because its expression levels were altered by lithium as the result of our previous study[6].

The box plots show relative gene expression of *HEY1*, *KLF1*, *PTP4A3*, and *POU3F1* plotted against sample groups. Down-regulated expression of *HEY1*, *KLF1*, and *PTP4A3*, while up-regulated expression of *POU3F1* were found in BD LR relatively compared to BD NR.

Each gene was conducted in the three sample groups: controls (*n*=2; black), BD LR (*n*=2; blue), and BD NR (*n*=3; red). Each dot represents an independent value of each triplet sample. The number in parenthesis on the x-axis are those with a completeness of data used to produce the plots. Relative gene expression was obtained by the $2^{-\Delta\Delta Ct}$ method and presented as fold-change (FC) in a $\log_2$ scale on the y-axis. *HPRT1* was used as a reference gene. The expression values of each gene are in comparison with BD NR (equal to 1). The box indicates the 25% and 75% percentiles. The median value is indicated by a horizontal line in each box. Error bars represent the standard deviation of the mean (SEM).

CT, controls; LR, BD Li responders; NR, BD Li non-responders; RNA-seq, RNA-sequencing; RT-qPCR, real time quantitative PCR.

#### Supplementary Figure 5: Preliminary functional enrichment results for 41 protein-coding DE genes in LR vs NR comparisons using WebGestalt[7] (a) and g:Profiler[8] (b) tools.

### a-1

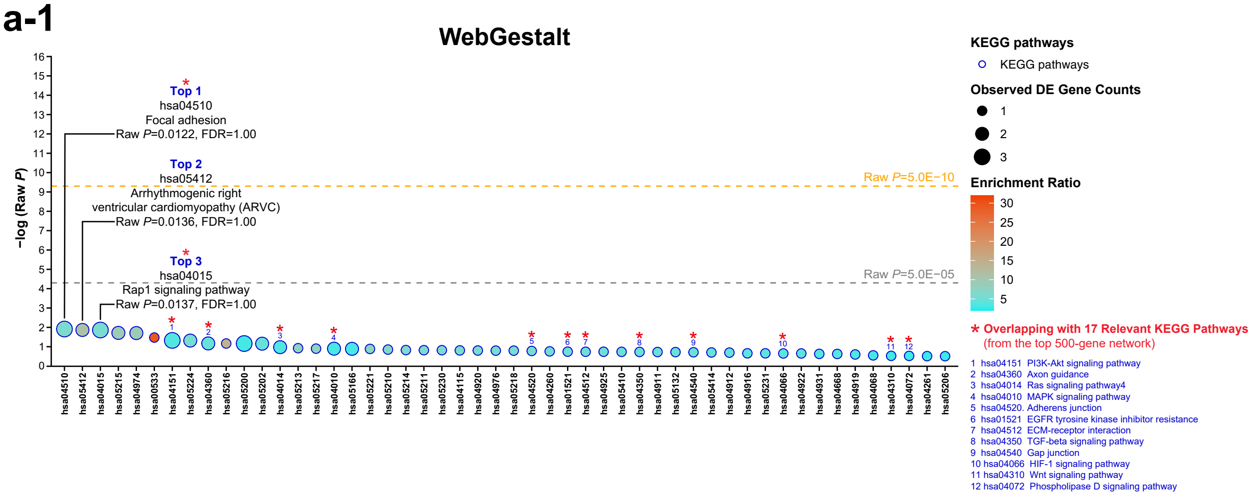

### a-2

| Source | KEGG ID | KEGG Name | Number of Ref Genes | Number of Query Genes | Obs Number of Genes | Exp Number of Genes | Enrichment Ratio | Raw *P*-value | B-H *FDR* |
|---|---|---|---|---|---|---|---|---|---|
| KEGG | hsa04510 | Focal adhesion | 203 | 41 | 3 | 0.49448345 | 6.06693712 | 0.01220106 | 1 |
| KEGG | hsa05412 | Arrhythmogenic right ventricular cardiomyopathy (ARVC) | 74 | 41 | 2 | 0.18025505 | 11.09538951 | 0.01360939 | 1 |
| KEGG | hsa04015 | Rap1 signaling pathway | 212 | 41 | 3 | 0.51640636 | 5.80937847 | 0.01371815 | 1 |
| KEGG | hsa05215 | Prostate cancer | 89 | 41 | 2 | 0.21679324 | 9.22538004 | 0.01931390 | 1 |
| KEGG | hsa04974 | Protein digestion and absorption | 90 | 41 | 2 | 0.21922912 | 9.12287582 | 0.01972473 | 1 |
| KEGG | hsa00533 | Glycosaminoglycan biosynthesis-keratan sulfate | 14 | 41 | 1 | 0.03410231 | 29.32352941 | 0.03359839 | 1 |
| KEGG | hsa04151 | PI3K-Akt signaling pathway | 341 | 41 | 3 | 0.83063476 | 3.61169571 | 0.04724952 | 1 |
| KEGG | hsa05224 | Breast cancer | 146 | 41 | 2 | 0.35563834 | 5.62369057 | 0.04814320 | 1 |
| KEGG | hsa04360 | Axon guidance | 177 | 41 | 2 | 0.43115060 | 4.63875042 | 0.06780170 | 1 |
| KEGG | hsa05216 | Thyroid cancer | 29 | 41 | 1 | 0.07064049 | 14.15618661 | 0.06841618 | 1 |
| KEGG | hsa05200 | Pathways in cancer | 397 | 41 | 3 | 0.96704399 | 3.10223737 | 0.06861158 | 1 |
| KEGG | hsa05202 | Transcriptional misregulation in cancer | 180 | 41 | 2 | 0.43845823 | 4.56143791 | 0.06982946 | 1 |
| KEGG | hsa04014 | Ras signaling pathway | 229 | 41 | 2 | 0.55781631 | 3.58540971 | 0.10560419 | 1 |
| KEGG | hsa05213 | Endometrial cancer | 52 | 41 | 1 | 0.12666571 | 7.89479638 | 0.11951860 | 1 |
| KEGG | hsa05217 | Basal cell carcinoma | 55 | 41 | 1 | 0.13397335 | 7.46417112 | 0.12598617 | 1 |
| KEGG | hsa04010 | MAPK signaling pathway | 255 | 41 | 2 | 0.62114916 | 3.21983852 | 0.12630639 | 1 |
| KEGG | hsa05166 | HTLV-I infection | 258 | 41 | 2 | 0.62845680 | 3.18239854 | 0.12875886 | 1 |
| KEGG | hsa05221 | Acute myeloid leukemia | 57 | 41 | 1 | 0.13884511 | 7.20227038 | 0.13027302 | 1 |
| KEGG | hsa05210 | Colorectal cancer | 62 | 41 | 1 | 0.15102450 | 6.62144213 | 0.14090372 | 1 |
| KEGG | hsa05214 | Glioma | 66 | 41 | 1 | 0.16076802 | 6.22014260 | 0.14932011 | 1 |
| KEGG | hsa05211 | Renal cell carcinoma | 67 | 41 | 1 | 0.16320390 | 6.12730465 | 0.15141204 | 1 |
| KEGG | hsa05230 | Central carbon metabolism in cancer | 67 | 41 | 1 | 0.16320390 | 6.12730465 | 0.15141204 | 1 |
| KEGG | hsa04115 | p53 signaling pathway | 69 | 41 | 1 | 0.16807566 | 5.94970162 | 0.15558140 | 1 |
| KEGG | hsa04920 | Adipocytokine signaling pathway | 70 | 41 | 1 | 0.17051154 | 5.86470588 | 0.15765884 | 1 |
| KEGG | hsa04976 | Bile secretion | 71 | 41 | 1 | 0.17294741 | 5.78210439 | 0.15973147 | 1 |
| KEGG | hsa05218 | Melanoma | 71 | 41 | 1 | 0.17294741 | 5.78210439 | 0.15973147 | 1 |
| KEGG | hsa04520 | Adherens junction | 74 | 41 | 1 | 0.18025505 | 5.54769475 | 0.16592060 | 1 |
| KEGG | hsa04260 | Cardiac muscle contraction | 78 | 41 | 1 | 0.18999857 | 5.26319759 | 0.17410605 | 1 |
| KEGG | hsa01521 | EGFR tyrosine kinase inhibitor resistance | 81 | 41 | 1 | 0.19730620 | 5.06826434 | 0.18019545 | 1 |
| KEGG | hsa04512 | ECM-receptor interaction | 82 | 41 | 1 | 0.19974208 | 5.00645624 | 0.18221585 | 1 |
| KEGG | hsa04925 | Aldosterone synthesis and secretion | 82 | 41 | 1 | 0.19974208 | 5.00645624 | 0.18221585 | 1 |
| KEGG | hsa05410 | Hypertrophic cardiomyopathy (HCM) | 83 | 41 | 1 | 0.20217796 | 4.94613749 | 0.18423155 | 1 |
| KEGG | hsa04350 | TGF-beta signaling pathway | 84 | 41 | 1 | 0.20461384 | 4.88725490 | 0.18624258 | 1 |
| KEGG | hsa04911 | Insulin secretion | 85 | 41 | 1 | 0.20704972 | 4.82975779 | 0.18824895 | 1 |
| KEGG | hsa05132 | Salmonella infection | 86 | 41 | 1 | 0.20948560 | 4.77359781 | 0.19025065 | 1 |
| KEGG | hsa04540 | Gap junction | 88 | 41 | 1 | 0.21435736 | 4.66510695 | 0.19424014 | 1 |
| KEGG | hsa05414 | Dilated cardiomyopathy | 90 | 41 | 1 | 0.21922912 | 4.56143791 | 0.19821112 | 1 |
| KEGG | hsa04912 | GnRH signaling pathway | 92 | 41 | 1 | 0.22410087 | 4.46227622 | 0.20216368 | 1 |
| KEGG | hsa04916 | Melanogenesis | 101 | 41 | 1 | 0.24602379 | 4.06464764 | 0.21972432 | 1 |
| KEGG | hsa05231 | Choline metabolism in cancer | 101 | 41 | 1 | 0.24602379 | 4.06464764 | 0.21972432 | 1 |
| KEGG | hsa04066 | HIF-1 signaling pathway | 103 | 41 | 1 | 0.25089554 | 3.98572244 | 0.22357697 | 1 |
| KEGG | hsa04922 | Glucagon signaling pathway | 103 | 41 | 1 | 0.25089554 | 3.98572244 | 0.22357697 | 1 |
| KEGG | hsa04931 | Insulin resistance | 109 | 41 | 1 | 0.26551082 | 3.76632488 | 0.23502777 | 1 |
| KEGG | hsa04668 | TNF signaling pathway | 110 | 41 | 1 | 0.26794670 | 3.73208556 | 0.23692072 | 1 |
| KEGG | hsa04919 | Thyroid hormone signaling pathway | 118 | 41 | 1 | 0.28743373 | 3.47906281 | 0.25190635 | 1 |
| KEGG | hsa04068 | FoxO signaling pathway | 134 | 41 | 1 | 0.32640780 | 3.06365233 | 0.28105051 | 1 |
| KEGG | hsa04310 | Wnt signaling pathway | 143 | 41 | 1 | 0.34833071 | 2.87083505 | 0.29697102 | 1 |
| KEGG | hsa04072 | Phospholipase D signaling pathway | 144 | 41 | 1 | 0.35076659 | 2.85089869 | 0.29871934 | 1 |
| KEGG | hsa04261 | Adrenergic signaling in cardiomyocytes | 149 | 41 | 1 | 0.36294598 | 2.75523095 | 0.30739971 | 1 |
| KEGG | hsa05206 | MicroRNAs in cancer | 150 | 41 | 1 | 0.36538186 | 2.73686275 | 0.30912361 | 1 |

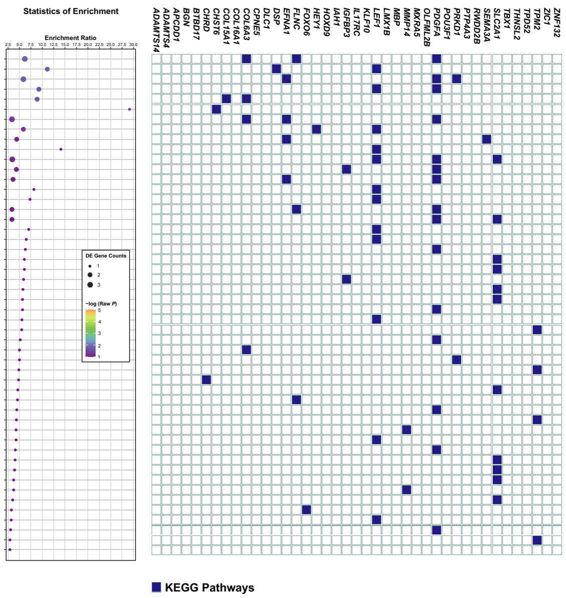

##### Summary and Parameters

**Enrich method:** Over-Representation Analysis (ORA)
**Organism:** Homo sapiens
**Enrichment categories:** KEGG pathway
**Interesting gene list:** 41 DE genes
**ID type:** Gene symbol
The interesting gene list contains **41** user IDs in which **41** user IDs are unambiguously mapped to the unique Entrez Gene IDs and 0 user IDs are mapped to multiple Entrez Gene IDs or could not be mapped to any Entrez Gene ID.
The GO Slim summary are based upon the **41** unique Entrez Gene IDs.
Among the **41** unique Entrez Gene IDs, **17** IDs are annotated to the selected functional categories and also in the reference gene list, which are used for the enrichment analysis.
**Reference gene list:** all mapped Entrez Gene IDs from the selected platform genome_protein-coding
The reference gene list contains **20691** IDs and **6979** IDs are annotated to the selected functional categories that are used as the reference for the enrichment analysis.

**Parameters for the enrichment analysis:**
- **Minimum number of Entrez Gene IDs in the category:** 5
- **Maximum number of Entrez Gene IDs in the category:** 2000
- **FDR method:** B-H
- **Significance level:** Top 50

## b-1

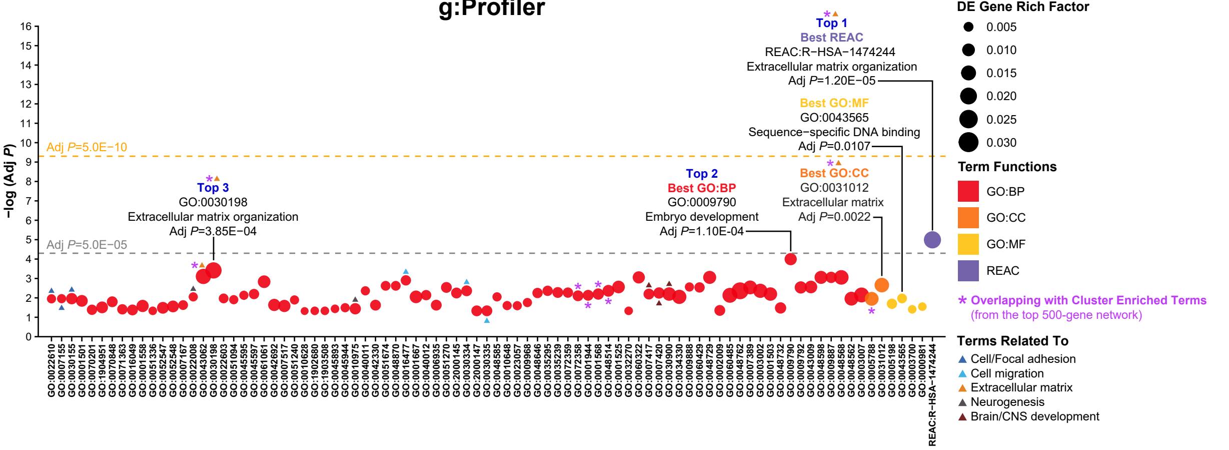

## b-2

| Source | Term ID | Term Name | No of Term Genes | No of Query Genes | No of Common Genes | Adjusted *P-value* (B-H *FDR*) |
|---|---|---|---|---|---|---|
| BP | GO:0022610 | Biological adhesion | 1351 | 41 | 10 | 1.12E-02 |
| BP | GO:0007155 | Cell adhesion | 1343 | 41 | 10 | 1.10E-02 |
| BP | GO:0030155 | Regulation of cell adhesion | 642 | 41 | 7 | 1.10E-02 |
| BP | GO:0001501 | Skeletal system development | 495 | 41 | 6 | 1.43E-02 |
| BP | GO:0070201 | Regulation of establishment of protein localization | 696 | 41 | 6 | 4.10E-02 |
| BP | GO:1904951 | Positive regulation of establishment of protein localization | 422 | 41 | 5 | 3.09E-02 |
| BP | GO:0070848 | Response to growth factor | 713 | 41 | 7 | 1.59E-02 |
| BP | GO:0071363 | Cellular response to growth factor stimulus | 685 | 41 | 6 | 3.89E-02 |
| BP | GO:0016049 | Cell growth | 482 | 41 | 5 | 4.19E-02 |
| BP | GO:0001558 | Regulation of cell growth | 400 | 41 | 5 | 2.61E-02 |
| BP | GO:0051336 | Regulation of hydrolase activity | 1256 | 41 | 8 | 4.66E-02 |
| BP | GO:0052547 | Regulation of peptidase activity | 431 | 41 | 5 | 3.25E-02 |
| BP | GO:0052548 | Regulation of endopeptidase activity | 406 | 41 | 5 | 2.77E-02 |
| BP | GO:0007167 | Enzyme linked receptor protein signaling pathway | 1031 | 41 | 8 | 2.35E-02 |
| BP | GO:0022008 | Neurogenesis | 1545 | 41 | 11 | 8.87E-03 |
| BP | GO:0043062 | Extracellular structure organization | 385 | 41 | 8 | 7.84E-04 |
| BP | GO:0030198 | Extracellular matrix organization | 332 | 41 | 8 | 3.85E-04 |
| BP | GO:0022603 | Regulation of anatomical structure morphogenesis | 1077 | 41 | 9 | 1.07E-02 |
| BP | GO:0051094 | Positive regulation of developmental process | 1378 | 41 | 10 | 1.24E-02 |
| BP | GO:0045595 | Regulation of cell differentiation | 1757 | 41 | 12 | 7.36E-03 |
| BP | GO:0045597 | Positive regulation of cell differentiation | 956 | 41 | 9 | 6.35E-03 |
| BP | GO:0061061 | Muscle structure development | 659 | 41 | 9 | 1.47E-03 |
| BP | GO:0042692 | Muscle cell differentiation | 381 | 41 | 5 | 2.31E-02 |
| BP | GO:0007517 | Muscle organ development | 400 | 41 | 5 | 2.61E-02 |
| BP | GO:0051240 | Positive regulation of multicellular organismal process | 1658 | 41 | 11 | 1.28E-02 |
| BP | GO:0010628 | Positive regulation of gene expression | 1863 | 41 | 10 | 4.79E-02 |
| BP | GO:1902680 | Positive regulation of RNA biosynthetic process | 1528 | 41 | 9 | 4.66E-02 |
| BP | GO:1903508 | Positive regulation of nucleic acid-templated transcription | 1527 | 41 | 9 | 4.66E-02 |
| BP | GO:0045893 | Positive regulation of transcription, DNA-templated | 1443 | 41 | 9 | 3.67E-02 |
| BP | GO:0045944 | Positive regulation of transcription by RNA polymerase II | 1112 | 41 | 8 | 3.21E-02 |
| BP | GO:0010975 | Regulation of neuron projection development | 454 | 41 | 5 | 3.64E-02 |
| BP | GO:0040011 | Locomotion | 1842 | 41 | 13 | 4.33E-03 |
| BP | GO:0042330 | Taxis | 581 | 41 | 6 | 2.37E-02 |
| BP | GO:0051674 | Localization of cell | 1609 | 41 | 13 | 2.39E-03 |
| BP | GO:0048870 | Cell motility | 1609 | 41 | 13 | 2.39E-03 |
| BP | GO:0016477 | Cell migration | 1466 | 41 | 13 | 1.25E-03 |
| BP | GO:0001667 | Ameboidal-type cell migration | 428 | 41 | 6 | 8.87E-03 |
| BP | GO:0040012 | Regulation of locomotion | 988 | 41 | 9 | 7.23E-03 |
| BP | GO:0006935 | Chemotaxis | 580 | 41 | 6 | 2.37E-02 |
| BP | GO:0051270 | Regulation of cellular component movement | 1000 | 41 | 10 | 2.90E-03 |
| BP | GO:2000145 | Regulation of cell motility | 915 | 41 | 9 | 5.61E-03 |
| BP | GO:0030334 | Regulation of cell migration | 861 | 41 | 9 | 4.33E-03 |
| BP | GO:2000147 | Positive regulation of cell motility | 513 | 41 | 5 | 4.67E-02 |
| BP | GO:0030335 | Positive regulation of cell migration | 496 | 41 | 5 | 4.64E-02 |
| BP | GO:0048585 | Negative regulation of response to stimulus | 1540 | 41 | 11 | 8.87E-03 |
| BP | GO:0010648 | Negative regulation of cell communication | 1311 | 41 | 9 | 2.50E-02 |
| BP | GO:0023057 | Negative regulation of signaling | 1315 | 41 | 9 | 2.53E-02 |
| BP | GO:0009968 | Negative regulation of signal transduction | 1206 | 41 | 9 | 1.74E-02 |
| BP | GO:0048646 | Anatomical structure formation involved in morphogenesis | 1144 | 41 | 10 | 5.61E-03 |
| BP | GO:0035295 | Tube development | 1079 | 41 | 10 | 4.33E-03 |
| BP | GO:0035239 | Tube morphogenesis | 899 | 41 | 9 | 5.29E-03 |
| BP | GO:0072359 | Circulatory system development | 1128 | 41 | 10 | 5.35E-03 |
| BP | GO:0072358 | Cardiovascular system development | 788 | 41 | 8 | 7.81E-03 |
| BP | GO:0001944 | Vasculature development | 779 | 41 | 8 | 7.36E-03 |
| BP | GO:0001568 | Blood vessel development | 743 | 41 | 8 | 6.35E-03 |
| BP | GO:0048514 | Blood vessel morphogenesis | 664 | 41 | 8 | 4.33E-03 |
| BP | GO:0001525 | Angiogenesis | 574 | 41 | 8 | 2.75E-03 |
| BP | GO:0032270 | Positive regulation of cellular protein metabolic process | 1526 | 41 | 9 | 4.66E-02 |
| BP | GO:0060322 | Head development | 754 | 41 | 10 | 8.80E-04 |
| BP | GO:0007417 | Central nervous system development | 942 | 41 | 9 | 6.35E-03 |
| BP | GO:0007420 | Brain development | 712 | 41 | 8 | 5.61E-03 |
| BP | GO:0030900 | Forebrain development | 379 | 41 | 6 | 6.35E-03 |
| BP | GO:0034330 | Cell junction organization | 272 | 41 | 5 | 8.87E-03 |
| BP | GO:0009888 | Tissue development | 1926 | 41 | 14 | 2.75E-03 |
| BP | GO:0060429 | Epithelium development | 1218 | 41 | 11 | 2.90E-03 |
| BP | GO:0048729 | Tissue morphogenesis | 602 | 41 | 9 | 8.80E-04 |
| BP | GO:0002009 | Morphogenesis of an epithelium | 490 | 41 | 5 | 4.44E-02 |
| BP | GO:0060485 | Mesenchyme development | 253 | 41 | 5 | 7.36E-03 |
| BP | GO:0048762 | Mesenchymal cell differentiation | 195 | 41 | 5 | 4.33E-03 |
| BP | GO:0007389 | Pattern specification process | 423 | 41 | 7 | 2.90E-03 |
| BP | GO:0003002 | Regionalization | 332 | 41 | 6 | 4.33E-03 |
| BP | GO:0001503 | Ossification | 380 | 41 | 6 | 6.35E-03 |
| BP | GO:0048732 | Gland development | 438 | 41 | 5 | 3.30E-02 |
| BP | GO:0009790 | Embryo development | 969 | 41 | 13 | 1.01E-04 |
| BP | GO:0009792 | Embryo development ending in birth or egg hatching | 593 | 41 | 8 | 2.90E-03 |
| BP | GO:0043009 | Chordate embryonic development | 574 | 41 | 8 | 2.75E-03 |
| BP | GO:0048598 | Embryonic morphogenesis | 590 | 41 | 9 | 8.80E-04 |
| BP | GO:0009887 | Animal organ morphogenesis | 969 | 41 | 11 | 8.80E-04 |
| BP | GO:0048568 | Embryonic organ development | 432 | 41 | 8 | 8.80E-04 |
| BP | GO:0048562 | Embryonic organ morphogenesis | 292 | 41 | 5 | 1.10E-02 |
| BP | GO:0003007 | Heart morphogenesis | 248 | 41 | 5 | 7.10E-03 |
| CC | GO:0005788 | Endoplasmic reticulum lumen | 298 | 41 | 5 | 1.12E-02 |
| CC | GO:0031012 | Extracellular matrix | 376 | 41 | 7 | 2.20E-03 |
| MF | GO:0005198 | Structural molecule activity | 763 | 41 | 7 | 2.02E-02 |
| MF | GO:0043565 | Sequence-specific DNA binding | 1076 | 41 | 9 | 1.07E-02 |
| MF | GO:0003700 | DNA-binding transcription factor activity | 1758 | 41 | 10 | 3.89E-02 |
| MF | GO:0000981 | DNA-binding transcription factor activity, RNA polymerase II-specific | 1626 | 41 | 10 | 2.83E-02 |
| rea | R-HSA-1474244 | Extracellular matrix organization | 296 | 22 | 8 | 1.02E-05 |

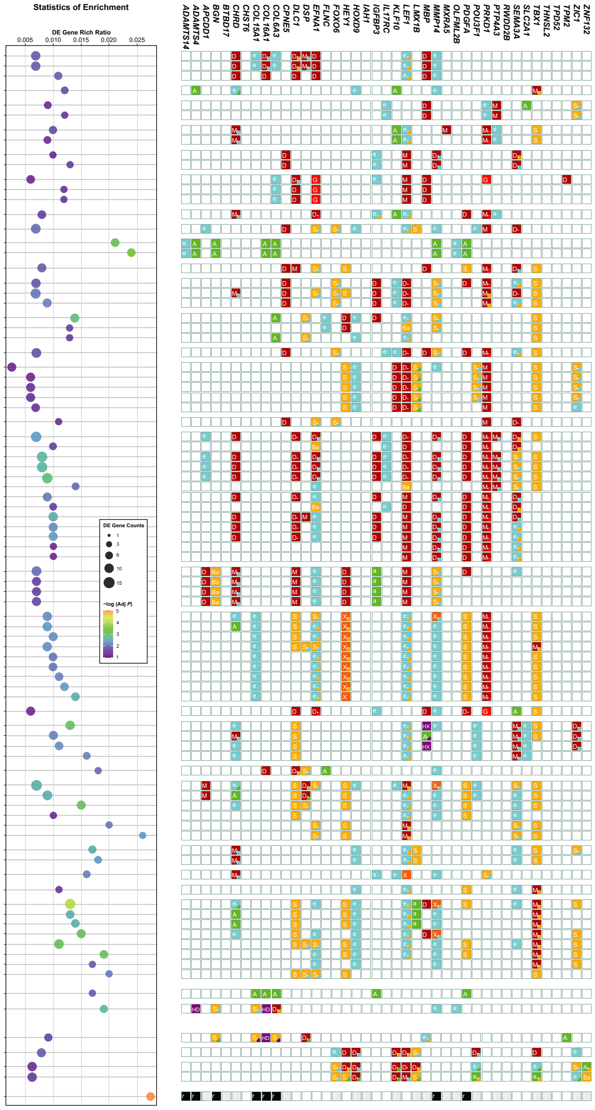

## b-3

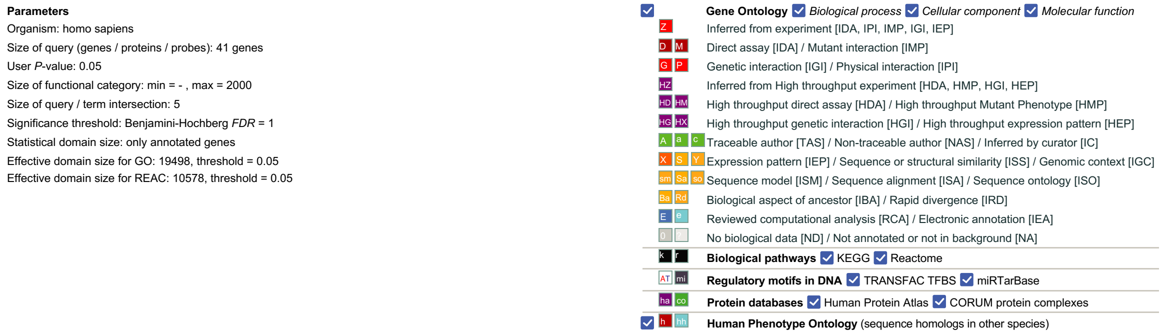

We used the WebGestalt[7] 2017 (http://www.webgestalt.org/2017/option.php#) and the g:Profiler[8] version r1760_e93_eg40 (https://biit.cs.ut.ee/gprofiler_archive2/r1760_e93_eg40/web/) web tools to analyze the preliminary functional enrichment of the 41 protein-coding DE genes. Additional details of these two web tools are described in **Supplemental Methods**. These 41 protein-coding DE genes were the RNA-seq genes with significantly different expression ($P$ <0.05, $\log_2$fold-change ≥|1|, and B-H $q$ ≤0.20; **Figures 1c**, **d**). The entire list and detailed differential expression of 41 DE genes in LR vs NR comparisons are shown in **Supplementary Table 8**.

**5a, Preliminary enrichment functions for 41 protein-coding DE genes from WebGestalt: The top 50 enriched pathways with Raw *P*-value <0.05.**
In brief, no KEGG pathways showed significant enrichment (B-H *FDR* <1.0E-05), since each pathway did not pass B-H *FDR* <1.00. The top pathway with the smallest Raw *P*-value was ‘hsa04510: focal adhesion’ (Raw $P$=0.0122; B-H $FDR$=1.0). Out of a total of 50 KEGG pathways, 14 (28.0%) pathways, including the top 1 (hsa04510: focal adhesion) and top 3 (hsa04015: Rap1 signaling pathway), overlapped with the KEGG pathway enrichment analysis of the top 500-proximal gene network in the current study. Note that ‘focal adhesion’ was shown to be the most significant pathway identified by the preliminary WebGestalt and our KEGG pathway enrichment analysis (see **Figure 4**; **Supplementary Figure 7**).

(**i**) **Bubble plot of WebGestalt enrichment analysis result (a-1).**
The bubble plot shows the top 50 significantly enriched KEGG pathways with Raw *P*-values <0.05. The x-axis indicates the corresponding KEGG IDs. The y-axis indicates the significance of enrichment represented as -log (Raw *P*). The size of bubbles indicates the number of observed DE genes in each corresponding pathway. Colors on bubbles indicate the enrichment ratio (low, blue; high, orange). The enrichment ratio is the ratio of observed number of genes divided by expected number of genes from the gene list of each KEGG pathway (see **a-2**, *output* panel). The larger enrichment ratio represents the greater number of observed genes in the interesting gene set. An asterisk (*) specifies the pathways that were also enriched in 17 KEGG pathways relevant to BD/neuronal system in our study. The orange dashed line on the y-axis

indicates the Raw *P*-value of 5.0E-10. The grey dashed line on the y-axis indicates the Raw *P*-value of 5.0E-05.

(**ii**) **Detailed enrichment output of WebGestalt enrichment analysis result (a-2).** Enrichment output is listed as follows: source, KEGG ID, KEGG name, number of genes (reference genes, query genes, observed number of genes, and expected number of genes), enrichment ratio, Raw *P*-value (*P*-value from hypergeometric test), B-H *FDR*, statistics of enrichment, query genes' involvements.

The bubble plot shows the statistics of enrichment for each of the enriched pathways. The size of bubbles represents the number of DE genes (gene counts) in each corresponding pathway. Location of bubbles on the x-axis represents the enrichment ratio (the ratio of observed number of genes divided by expected number of genes from the gene list of each pathway). The color scale indicates the degree of significance (Raw *P*-value ≤0.05) of enrichment for each corresponding term (low, purple; high, orange). The significance of enrichment is presented as the -log transformed Raw *P*-value.

Detailed parameters and legends of analysis result are described *below* the output panel.

**5b, Preliminary enrichment functions for 41 protein-coding DE genes from g:Profiler 2017: Total 88 enriched terms with *P*-value <0.05 and Adj *P* (B-H *FDR*) <0.05.**
Overall, no enriched terms showed strong significance, as a threshold was set at *P*-value <0.05 and Adj *P*-value (B-H *FDR*) <1.0E-05. The top 1 term was 'REAC:R-HSA-147244 Extracellular matrix organization' (Adj *P*=1.02E-05). Out of a total of 88, 15 (17.0%) terms were related to cell/focal adhesion (*n*=3), cell migration (*n*=3), ECM (*n*=4), and nervous system development (*n*=5); nine (10.2%, including four related to ECM) terms overlapped with the cluster enrichment of the top 500-proximal gene network in the current study. Remarkably, 'ECM' appeared to be the major findings identified by the preliminary g:Profiler and our cluster enrichment analysis (see **Figure 3**; **Supplementary Figure 6a**).

(**i**) **Plot of g:Profiler enrichment analysis result (b-1).**
The plot shows a total of 88 significantly enriched terms with *P*-value <0.05. The x-axis indicates the corresponding term IDs. The y-axis indicates the significance of enrichment represented as -log (Adj *P*). The size of points indicates the DE gene rich factor. The DE gene rich factor is the ratio of DE genes in each term to total genes in each term (see **b-2**). The larger gene rich factor represents the greater enrichment. Colors on points indicate the types of terms (GO term categories: BP, CC, MF; REAC). An asterisk (*) specifies the terms that were also enriched in our cluster enrichment analysis, all of which were enriched in cluster 0. A triangle (▲) indicates specific functions that terms are related to: cell/focal adhesion (dark blue), cell migration, (light blue), ECM (orange), neurogenesis (dark grey), and brain/central nervous system (CNS) development (brown). The orange dashed line on the y-axis indicates the Adj *P*-

value of 5.0E-10. The grey dashed line on the y-axis indicates the Adj *P*-value of 5.0E-05.

(ii) **Detailed enrichment output of g:Profiler enrichment analysis result (b-2).** Enrichment output is listed as follows: term source (term type), term ID, term name, number of genes (term genes, query genes, common genes (intersection between term genes and query genes), Adj *P*-value (B-H *FDR*), statistics of enrichment, and query genes' involvements.

The bubble plot shows the statistics of enrichment for each of the enriched terms. The size of bubbles represents the number of DE genes (DE gene counts) in each corresponding term. Location of bubbles on the x-axis represents the degree of enrichment for DE genes known as 'DE gene rich factor' (the ratio of DE genes in each term to total genes in each term). The color scale indicates the degree of significance (Adj *P*-value ≤0.05) in enrichment for each corresponding term (low, purple; high, orange). The significance of enrichment is presented as the -log transformed Adj *P*-value.

(iii) **Detailed parameters and legends of g:Profiler enrichment analysis result (b-3).**

B-H, Benjamini and Hochberg; BP, biological process; CC, cellular component; DE, differentially expressed; ECM, the extracellular matrix; GO, the Gene Ontology; FDR, false discovery rate; LR, BD Li responders; MF, molecular function; NR, BD Li non-responders; REAC, the REACTOME pathway database (https://reactome.org/).

#### Supplementary Figure 6: Functional enrichment analysis result for the three clusters.

**a**

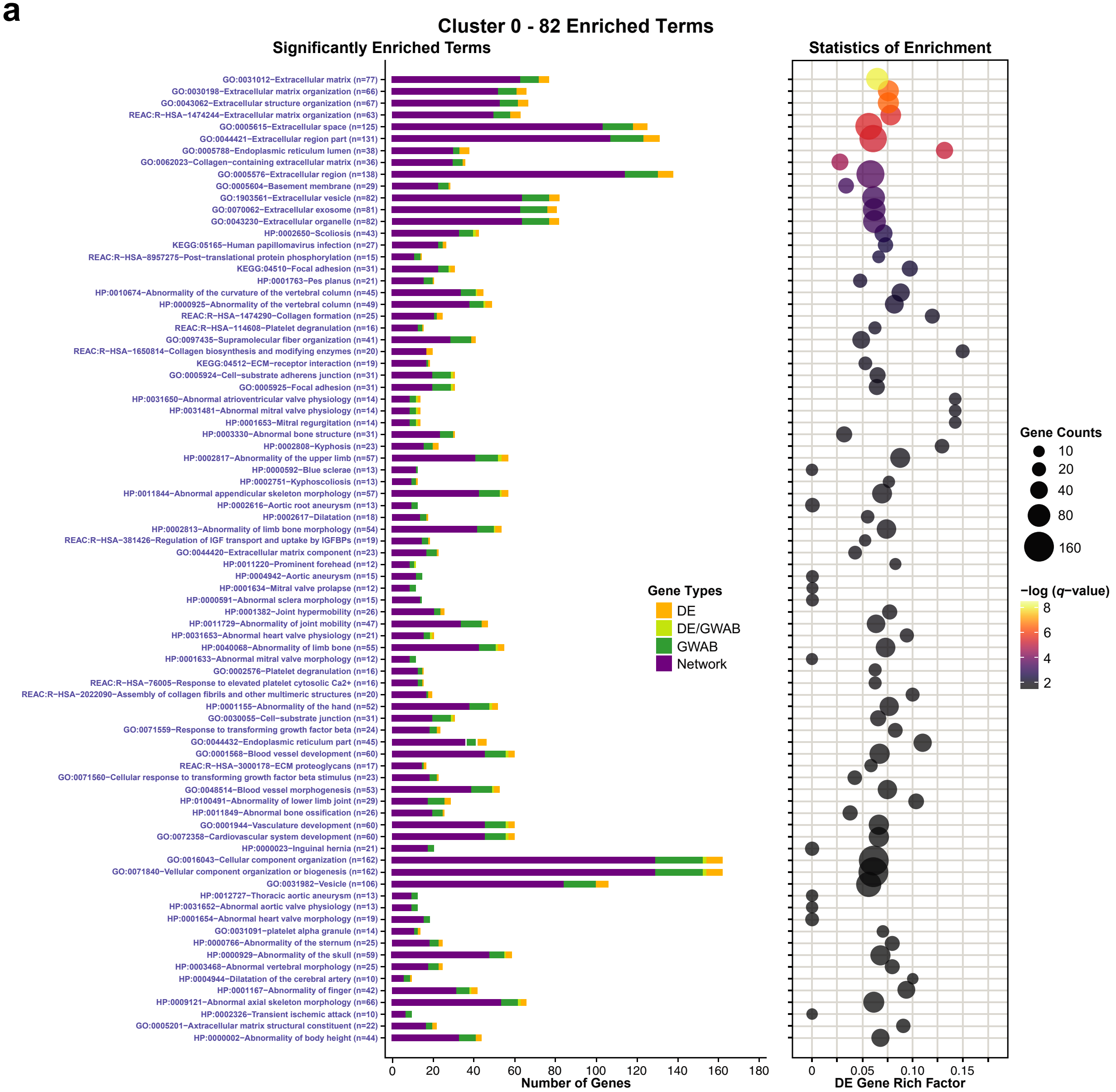

b

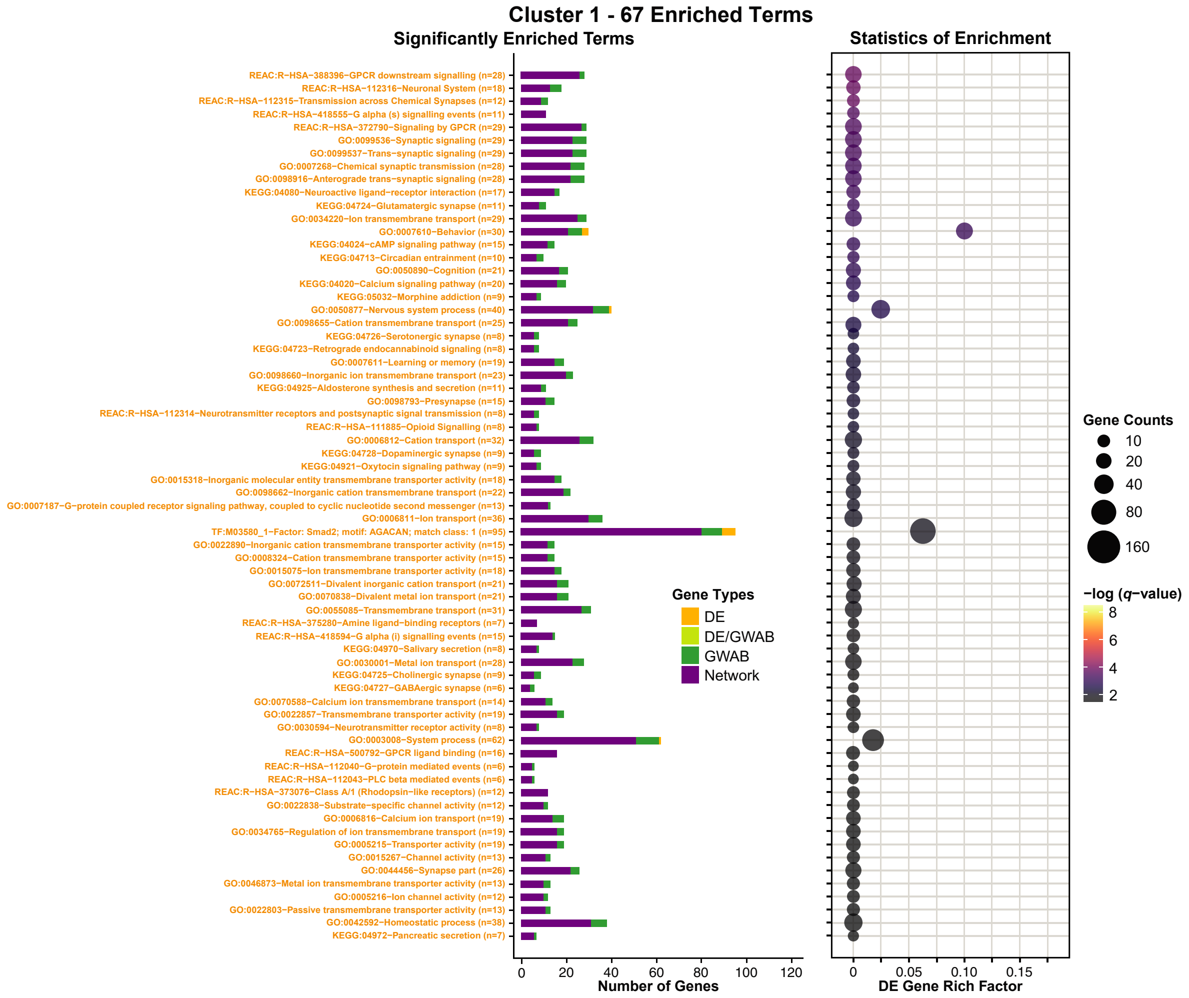

Cluster 1 - 67 Enriched Terms
Significantly Enriched Terms
Statistics of Enrichment
REAC:R−HSA−388396−GPCR downstream signalling (n=28)
REAC:R−HSA−112316−Neuronal System (n=18)
REAC:R−HSA−112315−Transmission across Chemical Synapses (n=12)
REAC:R−HSA−418555−G alpha (s) signalling events (n=11)
REAC:R−HSA−372790−Signaling by GPCR (n=29)
GO:0099536−Synaptic signaling (n=29)
GO:0099537−Trans−synaptic signaling (n=29)
GO:0007268−Chemical synaptic transmission (n=28)
GO:0098916−Anterograde trans−synaptic signaling (n=28)
KEGG:04080−Neuroactive ligand−receptor interaction (n=17)
KEGG:04724−Glutamatergic synapse (n=11)
GO:0034220−Ion transmembrane transport (n=29)
GO:0007610−Behavior (n=30)
KEGG:04024−cAMP signaling pathway (n=15)
KEGG:04713−Circadian entrainment (n=10)
GO:0050890−Cognition (n=21)
KEGG:04020−Calcium signaling pathway (n=20)
KEGG:05032−Morphine addiction (n=9)
GO:0050877−Nervous system process (n=40)
GO:0098655−Cation transmembrane transport (n=25)
KEGG:04726−Serotonergic synapse (n=8)
KEGG:04723−Retrograde endocannabinoid signaling (n=8)
GO:0007611−Learning or memory (n=19)
GO:0098660−Inorganic ion transmembrane transport (n=23)
KEGG:04925−Aldosterone synthesis and secretion (n=11)
GO:0098793−Presynapse (n=15)
REAC:R−HSA−112314−Neurotransmitter receptors and postsynaptic signal transmission (n=8)
REAC:R−HSA−111885−Opioid Signalling (n=8)
GO:0006812−Cation transport (n=32)
KEGG:04728−Dopaminergic synapse (n=9)
KEGG:04921−Oxytocin signaling pathway (n=9)
GO:0015318−Inorganic molecular entity transmembrane transporter activity (n=18)
GO:0098662−Inorganic cation transmembrane transport (n=22)
GO:0007187−G−protein coupled receptor signaling pathway, coupled to cyclic nucleotide second messenger (n=13)
GO:0006811−Ion transport (n=36)
TF:M03580_1−Factor: Smad2; motif: AGACAN; match class: 1 (n=95)
GO:0022890−Inorganic cation transmembrane transporter activity (n=15)
GO:0008324−Cation transmembrane transporter activity (n=15)
GO:0015075−Ion transmembrane transporter activity (n=18)
GO:0072511−Divalent inorganic cation transport (n=21)
GO:0070838−Divalent metal ion transport (n=21)
GO:0055085−Transmembrane transport (n=31)
REAC:R−HSA−375280−Amine ligand−binding receptors (n=7)
REAC:R−HSA−418594−G alpha (i) signalling events (n=15)
KEGG:04970−Salivary secretion (n=8)
GO:0030001−Metal ion transport (n=28)
KEGG:04725−Cholinergic synapse (n=9)
KEGG:04727−GABAergic synapse (n=6)
GO:0070588−Calcium ion transmembrane transport (n=14)
GO:0022857−Transmembrane transporter activity (n=19)
GO:0030594−Neurotransmitter receptor activity (n=8)
GO:0003008−System process (n=62)
REAC:R−HSA−500792−GPCR ligand binding (n=16)
REAC:R−HSA−112040−G−protein mediated events (n=6)
REAC:R−HSA−112043−PLC beta mediated events (n=6)
REAC:R−HSA−373076−Class A/1 (Rhodopsin−like receptors) (n=12)
GO:0022838−Substrate−specific channel activity (n=12)
GO:0006816−Calcium ion transport (n=19)
GO:0034765−Regulation of ion transmembrane transport (n=19)
GO:0005215−Transporter activity (n=19)
GO:0015267−Channel activity (n=13)
GO:0044456−Synapse part (n=26)
GO:0046873−Metal ion transmembrane transporter activity (n=13)
GO:0005216−Ion channel activity (n=12)
GO:0022803−Passive transmembrane transporter activity (n=13)
GO:0042592−Homeostatic process (n=38)
KEGG:04972−Pancreatic secretion (n=7)
0 20 40 60 80 100 120
Number of Genes
0 0.05 0.10 0.15
DE Gene Rich Factor
Gene Types
DE
DE/GWAB
GWAB
Network
Gene Counts
10
20
40
80
160
−log (q−value)
8
6
4
2

c

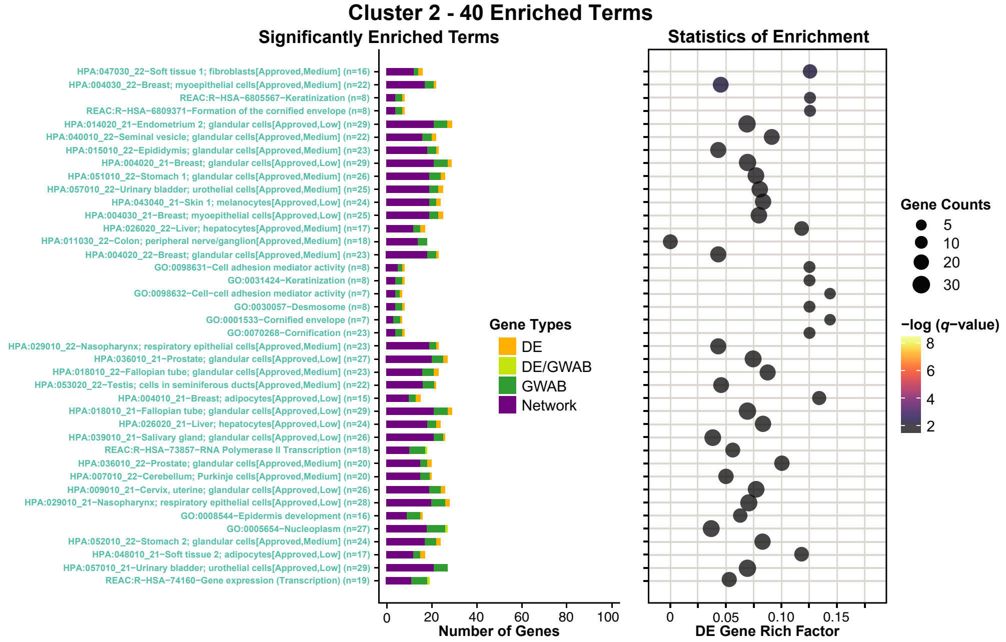

Cluster 2 - 40 Enriched Terms
Significantly Enriched Terms
Statistics of Enrichment
HPA:047030_22−Soft tissue 1; fibroblasts[Approved,Medium] (n=16)
HPA:004030_22−Breast; myoepithelial cells[Approved,Medium] (n=22)
REAC:R−HSA−6805567−Keratinization (n=8)
REAC:R−HSA−6809371−Formation of the cornified envelope (n=8)
HPA:014020_21−Endometrium 2; glandular cells[Approved,Low] (n=29)
HPA:040010_22−Seminal vesicle; glandular cells[Approved,Medium] (n=22)
HPA:015010_22−Epididymis; glandular cells[Approved,Medium] (n=23)
HPA:004020_21−Breast; glandular cells[Approved,Low] (n=29)
HPA:051010_22−Stomach 1; glandular cells[Approved,Medium] (n=26)
HPA:057010_22−Urinary bladder; urothelial cells[Approved,Medium] (n=25)
HPA:043040_21−Skin 1; melanocytes[Approved,Low] (n=24)
HPA:004030_21−Breast; myoepithelial cells[Approved,Low] (n=25)
HPA:026020_22−Liver; hepatocytes[Approved,Medium] (n=17)
HPA:011030_22−Colon; peripheral nerve/ganglion[Approved,Medium] (n=18)
HPA:004020_22−Breast; glandular cells[Approved,Medium] (n=23)
GO:0098631−Cell adhesion mediator activity (n=8)
GO:0031424−Keratinization (n=8)
GO:0098632−Cell−cell adhesion mediator activity (n=7)
GO:0030057−Desmosome (n=8)
GO:0001533−Cornified envelope (n=7)
GO:0070268−Cornification (n=23)
HPA:029010_22−Nasopharynx; respiratory epithelial cells[Approved,Medium] (n=23)
HPA:036010_21−Prostate; glandular cells[Approved,Low] (n=27)
HPA:018010_22−Fallopian tube; glandular cells[Approved,Medium] (n=23)
HPA:053020_22−Testis; cells in seminiferous ducts[Approved,Medium] (n=22)
HPA:004010_21−Breast; adipocytes[Approved,Low] (n=15)
HPA:018010_21−Fallopian tube; glandular cells[Approved,Low] (n=29)
HPA:026020_21−Liver; hepatocytes[Approved,Low] (n=24)
HPA:039010_21−Salivary gland; glandular cells[Approved,Low] (n=26)
REAC:R−HSA−73857−RNA Polymerase II Transcription (n=18)
HPA:036010_22−Prostate; glandular cells[Approved,Medium] (n=20)
HPA:007010_22−Cerebellum; Purkinje cells[Approved,Medium] (n=20)
HPA:009010_21−Cervix, uterine; glandular cells[Approved,Low] (n=26)
HPA:029010_21−Nasopharynx; respiratory epithelial cells[Approved,Low] (n=28)
GO:0008544−Epidermis development (n=16)
GO:0005654−Nucleoplasm (n=27)
HPA:052010_22−Stomach 2; glandular cells[Approved,Medium] (n=24)
HPA:048010_21−Soft tissue 2; adipocytes[Approved,Low] (n=17)
HPA:057010_21−Urinary bladder; urothelial cells[Approved,Low] (n=29)
REAC:R−HSA−74160−Gene expression (Transcription) (n=19)
0 20 40 60 80 100
Number of Genes
0 0.05 0.10 0.15
DE Gene Rich Factor
Gene Types
DE
DE/GWAB
GWAB
Network
Gene Counts
5
10
20
30
−log (q−value)
8
6
4
2

**d**

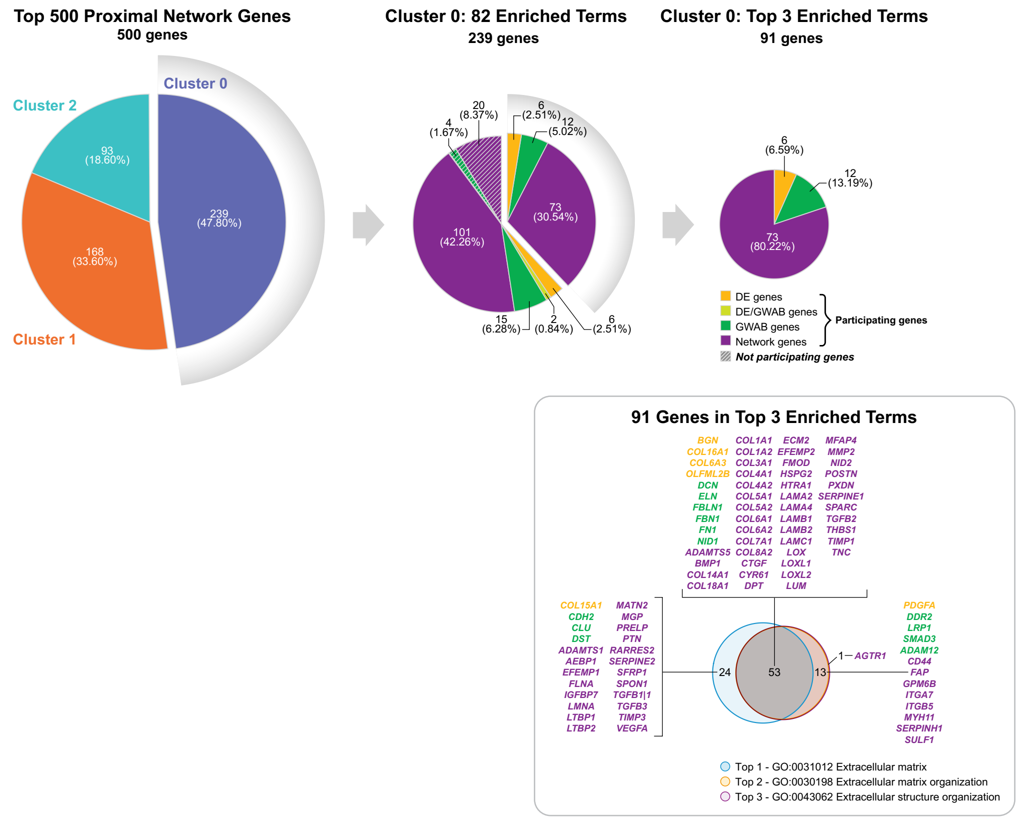

Functional enrichment analysis using cluster analysis[9] identified 82, 67, and 40 terms (B-H *q*-value ≤0.05) significantly enriched in clusters 0 (**a**), 1 (**b**), and 2 (**c**), respectively. A summary of the top 10 enriched terms for each cluster are shown in **Figure 3c**. Overall, the top 3 significant terms (**d**) were in cluster 0 as follows: top 1—'extracellular matrix' (GO:0031012; B-H *q*=1.01E-08), top 2—'extracellular matrix organization' (GO:0030198; B-H *q*=3.25E-07), and top 3—'extracellular structure organization' (GO:0043062; B-H *q*=3.25E-07). The most significant term in cluster 1 was GPCR downstream signaling (REAC:R-HAS-388396; B-H *q*=1.57E-04) and in cluster 2 were soft tissue 1; fibroblasts (HPA:047030_22) and breast; myoepithelial cells (HPA:004030_22) (B-H *q*=0.00541 for both). Cluster analysis is detailed in **Supplemental Methods**. The distribution of genes for each cluster are summarized in **Figures 2e** and **3a**. The entire lists and detailed significantly enriched terms for clusters 0, 1, and 2 are described in **Supplementary Tables 12**, **13**, and **14**, respectively.

**6a, Significantly enriched functional terms for Cluster 0.**
**6b, Significantly enriched functional terms for Cluster 1.**
**6c, Significantly enriched functional terms for Cluster 2.**

For **Figures 6a-c**, bar plot (*left* panel) shows the distribution of genes in each of the enriched terms for each cluster. The x-axis represents the number of genes (gene counts) and gene types in each corresponding pathway. The y-axis corresponds to enriched terms. Colors on bars indicate gene types.

The bubble plot (*right* panel) shows the statistics of enrichment for each of the enriched terms. The size of bubbles represents the number of genes (gene counts) in each corresponding term. The bubble coordinate on the x-axis represents the degree of enrichment for DE genes known as 'DE gene rich factor'. The DE gene rich factor is the ratio of DE genes in each term to total genes in each term. The larger the rich factor, the greater the enrichment. The color scale indicates the degree of significance (B-H *q*-value ≤0.05) in enrichment for each corresponding term (low, dark purple; high, yellow). The significance of enrichment is presented as the -log transformed B-H *q*-value.

**6d, Distribution and summarizing features of genes in cluster 0 including the top 3 significantly enriched terms.**
Pie charts (*top* panel) display the number of genes corresponding to a total of 500 proximal network genes (*left*), a set of 239 genes of cluster 0 (*middle*), and a set of 91 genes of the top 3 significant terms in cluster 0 (*right*). Each pie represents the proportion of gene types in numbers and percentages. Colors represent gene types. Solid and stripe patterns indicate function (participating) and non-function (non-participating) of genes, respectively, in each gene set of sub-charts.

The Venn diagram (*bottom right* panel) shows the 91 genes in the top 3 enriched terms in cluster analysis, containing 6 DE, 12 GWAB, and 73 network genes. In brief, there are 77, 66, and 67 genes involved in the top 1 (GO:0031012 'extracellular matrix'), top 2 (GO:0030198 'extracellular matrix organization'), and top 3 (GO:0043062 'extracellular structure organization') significant terms, respectively. Strikingly, all the top 3 terms were involving in the ECM and shared more than half (*n*=53) of the 91 genes among them. This indicates that the ECM was the main function in cluster 0. A total of 91 gene names are listed and categorized into gene types. Venn diagram colors indicate the top 3 significant terms.

Gene types are indicated by colors: DE, orange; DE/GWAB, light green; GWAB, green; network, purple.

B-H, Benjamini and Hochberg; DE, differentially expressed; ECM, the extracellular matrix; FDR, false discovery rate; GPCR, G-protein-coupled receptors; GO, the Gene Ontology; GWAB, genome-wide association boosting; HPA, Human Proteome Atlas; REAC, the REACTOME pathway database (https://reactome.org/).

#### Supplementary Figure 7: Functional enrichment analysis result for KEGG pathways.

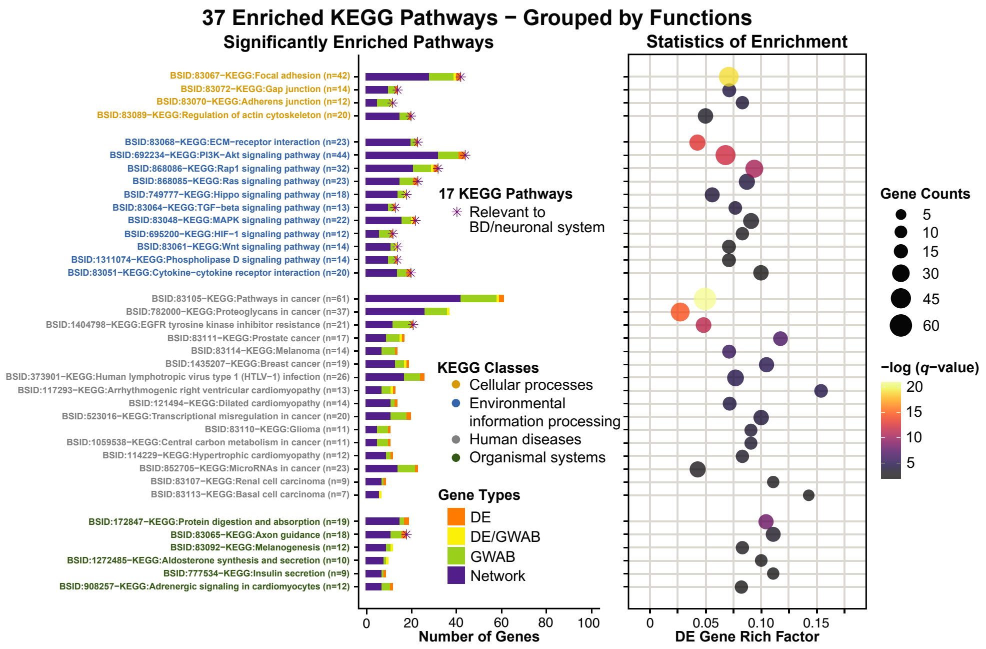

Functional enrichment analysis that was filtered to focus on KEGG (Kyoto Encyclopedia of Genes and Genomes) pathways (https://www.genome.jp/kegg/pathway.html) (containing at least one seed gene and one GWAB gene) identified 37 KEGG pathways (B-H *q*-value ≤0.05) significantly enriched in the top 500-proximal gene network, of which 17 were relevant to BD/neuronal system (marked with an asterisk). Overall, the most significant pathway among 37 enriched KEGG pathways was 'pathways in cancer' (hsa05200; B-H *q*=1.05E-21), followed by 'focal adhesion' (hsa04510; B-H *q*=8.04-E20), which also was the top pathway among a set of 17 relevant enriched KEGG pathways. Detailed KEGG pathway analysis can be seen in **Supplemental Methods**. The distribution of genes for 37 and 17 enriched KEGG pathways are summarized in **Figures 4a** and **b**. The details for each pathway are described in **Supplementary Table 15**.

Bar plot (*left* panel) shows the distribution of genes in each enriched pathway. The x-axis represents the number of genes (gene counts) and gene types in each corresponding pathway. The y-axis corresponds to enriched pathways, which were classified into four functional classes: cellular processes (yellow), environmental information processing (blue), human diseases (grey), and organismal systems (green). Colors on bars indicate gene types (DE, orange; DE/GWAB, yellow; GWAB, green; network, purple). An asterisk (*) on bars specifies the KEGG pathways that are relevant to BD/neuronal system.

Bubble plot (*right* panel) shows the statistics of enrichment of each of the enriched pathways. The size of bubbles represents the number of genes (gene counts) in each corresponding pathway. The bubble coordinate on the x-axis represents the degree of enrichment for DE genes known as 'DE gene rich factor'. The DE gene rich factor is the ratio of DE genes in each pathway to total genes in each pathway. The larger the rich factor, the greater the enrichment. The color scale indicates the degree of significance (B-H *q*-value ≤0.05) in enrichment for each corresponding pathway (low, dark purple; high, yellow). The significance of enrichment is presented as the -log transformed B-H *q*-value.

B-H, Benjamini and Hochberg; DE, differentially expressed; GWAB, genome-wide association boosting.
