## Supplementary Tables for "Network-based integrative analysis of lithium response in bipolar disorder using transcriptomic and GWAS data": Suppl_Table1_Characteristics_GWAS_Dec2021.pdf

**Supplementary Table 1.** Characteristics of the PGBD/VA patients used for the genome-wide association analysis.

| | Patients From The PGBD/VA Study Entered Maintenance | | | |
|---|---|---|---|---|
| | ***N*** | **BD Li Responders** | **BD Li Non-Responders** | ***P**** |
| **Total, *n* (%)** | 256 | 157 (62%) | 99 (38%) | - |
| **Ethnicity, %EUR** | 256 | 100% | 100% | NS |
| **Gender, M : F (%M)** | 256 | 71 : 86 (45%) | 48 : 51 (48%) | NS |
| **Time in the study†, Mean days ± SD** | 256 | 463.6 ± 18.0 | 57.9 ± 22.7 | <0.0001 |
| **Age at onset, Mean yrs ± SD (*n* completed)** | 256 | 20.1 ± 13 (156) | 17.9 ± 11 (98) | NS |
| **Family history of BD‡** | 222 | 63/136 (46%) | 41/86 (48%) | NS |
| **Psychotic features‡** | 205 | 87/125 (70%) | 47/80 (59%) | NS |
| **Rapid cycling‡** | 174 | 21/109 (19%) | 31/65 (48%) | 0.0001 |
| **Suicide attempts‡** | 125 | 37/125 (29%) | 29/76 (38%) | NS |

See additional details of the PGBD/VA study in **Supplemental Methods**.
*Categorical variables were tested using contingency table $\chi^2$ analysis, continuous variables were tested by *t*-test. Note that data was not available on all subjects for some phenotypes.
†Time in study is calculated as the time as either the time to failure to remit or the time to relapse in those who achieve maintenance. Failure to remit is decided by the study physician who concludes an adequate trial resulted in no response and the patient should be removed from the study and treated clinically.
‡Units are presented as a proportion and percentage of those whose positive history and a total number of the data completeness.
NS, not significant.
PGBD/VA study, a study from the Pharmacogenomics of Bipolar Disorder study and the Veterans Affairs San Diego Healthcare System.
SD, the standard deviation.
