## Supplementary Tables for "Network-based integrative analysis of lithium response in bipolar disorder using transcriptomic and GWAS data": Supplemental_Table_02_Characteristics of iPSC_Dec11_2021_medRXiv_09Jan2022.pdf

**Supplementary Table 2.** Characteristics of patients used for the iPSC studies.

| Original Study | Cell ID | Obtained Tissues | Ethnicity | Gender | Age at Sampling (range, yrs) | Age of Onset (range, yrs) | Main Diagnosis | Time to Treatment Failure/Relapse – *Prospective Study* (mos) | Termination Reasons – *Prospective Study** | Alda Score (total) | Episodes, Depression+ Mania (n) | Lifetime Psychotic Features (Yes/No) |
|---|---|---|---|---|---|---|---|---|---|---|---|---|
| | | | | | | **Controls (*n*=6)** | | | | | | |
| PGBD/VA | 149-59 | Fibroblast | Caucasian | Male | 56-60 | - | None | - | - | - | - | - |
| PGBD/VA | 149-61 | Fibroblast | Caucasian | Male | 36-40 | - | None | - | - | - | - | - |
| HALIFAX | J008 | Lymphoblast | Caucasian | Male | 20-25 | - | - | - | - | - | - | - |
| HALIFAX | J009 | Lymphoblast | Caucasian | Male | 50-55 | - | - | - | - | - | - | - |
| HALIFAX | J011 | Lymphoblast | Caucasian | Male | 60-65 | - | - | - | - | - | - | - |
| HALIFAX | J012 | Lymphoblast | Caucasian | Male | 50-55 | - | - | - | - | - | - | - |
| | | | | | | **BD Responders (*n*=6)** | | | | | | |
| PGBD/VA | 144-74 | Fibroblast | Caucasian | Male | 60-65 | 36-40 | BD I | 24 | Completed | - | 9 | N |
| PGBD/VA | 118-22 | Fibroblast | Caucasian | Male | 56-60 | 6-10 | BD I | 22 | Relapse | - | 30 | N |
| PGBD/VA | 116-92 | Fibroblast | Caucasian | Male | 56-60 | 6-10 | BD I | 23 | Relapse | - | 24 | N |
| HALIFAX | J005 | Lymphoblast | Caucasian | Male | 40-45 | 30-35 | BD I | - | - | 10/10 | 5 | Y |
| HALIFAX | J007 | Lymphoblast | Caucasian | Male | 30-35 | 10-15 | BD I | - | - | 9/10 | 1 | N |
| HALIFAX | J010 | Lymphoblast | Caucasian | Male | 50-55 | 30-35 | BD I | - | - | 9/10 | | |
| | | | | | | **BD Non-Responders (*n*=5)** | | | | | | |
| PGBD/VA | 96-91 | Fibroblast | Caucasian | Male | 50-55 | 10-15 | BD I | 3 | Treatment failure | - | 41 | Y |
| PGBD/VA | 149-34 | Fibroblast | Caucasian | Male | 66-70 | 10-15 | BD I | 4 | Treatment failure | - | 11 | N |
| HALIFAX | J001 | Lymphoblast | Caucasian | Male | 50-55 | 30-35 | BD I | - | - | 3/10 | 3 | Y |
| HALIFAX | J002 | Lymphoblast | Caucasian | Male | 56-60 | 20-25 | BD I | - | - | 1/10 | 10 | N |
| HALIFAX | J004 | Lymphoblast | Caucasian | Male | 36-40 | 20-25 | BD I | - | - | 0/10 | 31 | N |

See additional details of the PGBD/VA and HALIFAX studies in **Supplemental Methods**.
***Completed** refers to those subjects who completed the 24-month maintenance follow-up phase without relapse; **Relapse** refers to those who completed the initial 4-month stabilization phase but relapsed in the maintenance phase follow-up; **Treatment failure** refers to those who failed to achieve remission during the initial 4-month stabilization phase.
HALIFAX, a study from Dalhouise University; PGBD/VA, a study from the Pharmacogenomics of Bipolar Disorder study and the Veterans Affairs San Diego Healthcare System.
